# Quantifying phenotype and genotype distributions in single muscle fibres from patients carrying the pathogenic mtDNA variant m.3243A>G

**DOI:** 10.1101/2022.04.04.22272484

**Authors:** Syeda T Ahmed, Robert W Taylor, Doug M Turnbull, Conor Lawless, Sarah J Pickett

**Affiliations:** Wellcome Centre for Mitochondrial Research, Institute of Translational and Clinical Research, Faculty of Medical Sciences, Newcastle University, Newcastle upon Tyne, NE2 4HH, UK. 2085685

## Abstract

**Background:** Pathogenic mitochondrial (mt)DNA variants cause neuromuscular disease with highly variable severity and phenotypic presentation, the reason for which is poorly understood. Cells are thought to tolerate the presence of pathogenic mtDNA variants up to a threshold proportion with little or no functional consequence, developing significant respiratory complex defects above this threshold. We developed a robust method to identify deficient muscle fibres, applied it to biopsies from 17 patients carrying the common m.3243A>G variant and examined the relationship between respiratory deficiency and m.3243A>G level in hundreds of single skeletal muscle fibres. We hypothesised that single-cell between-patient differences may explain the vast clinical heterogeneity of mtDNA disease.

**Results:** Immunohistochemical measurements of respiratory complexes I and IV and unsupervised machine learning identified muscle fibres with respiratory deficiency; the pattern of deficiency and proportion of deficient fibres (range 0-64%) varies between patients. Tissue homogenate m.3243A>G level is a poor surrogate for the broad and complex distributions of m.3243A>G level in single cells from individual patients. Estimated thresholds do not differ between patients, but sections with narrow m.3243A>G distributions have a lower proportion of deficient fibres.

**Conclusions:** Inter-individual differences in respiratory complex deficiency in muscle fibres from patients with m.3243A>G are more complex than previously thought and may be driven by differential segregation and expansion of mtDNA molecules. Our quantitative observations could constrain the range of feasible mechanisms responsible for phenotypic diversity in mitochondrial disease.

## BACKGROUND

The most common pathogenic mitochondrial (mt)DNA variant is an A to G transition at position 3243 (m.3243A>G) within the *MT-TL1* gene which encodes tRNA^Leu(UUR)^ and causes a mitochondrial translation defect (Goto et al. 1990; Manwaring et al. 2007; Elliott et al. 2008). Initially identified due to its association with a syndrome defined by mitochondrial encephalomyopathy and stroke-like episodes (MELAS), it is associated with highly variable disease severity, rate of progression and phenotypic presentation (Kaufmann et al. 2011; de Laat et al. 2012; Nesbitt et al. 2013; Chin et al. 2014; Mancuso et al. 2014; Grady et al. 2018). Individuals who carry m.3243A>G can be clinically asymptomatic or can suffer from a number of disorders including diabetes, deafness, myopathy, cardiac disease, stroke-like episodes and gastro-intestinal disturbances. The proportion of m.3243A>G mutant mtDNA varies between individuals, between tissues and between cells (Ozawa et al. 1998; Sue et al. 1998; Rahman et al. 2001; de Laat et al. 2012) and, along with other factors such as mtDNA copy number, age and nuclear background, contributes to this variability (reviewed in (Boggan et al. 2019; Richter et al. 2021)). Nevertheless, a large proportion of variability in both phenotypic presentation and disease severity remains to be explained (Chinnery et al. 1997; Fayssoil et al. 2017; Grady et al. 2018; Pickett et al. 2018).

Unlike nuclear DNA, mtDNA replicates continuously in post-mitotic tissue by a process called ‘relaxed replication’, which is independent of the cell cycle (Gross et al. 1969; Bogenhagen and Clayton 1977; Chinnery and Samuels 1999). mtDNA turnover is likely a driving factor behind the stochastic expansion of mtDNA variants, which are believed to contribute to age-related pathogenicity (Elson et al. 2001; Müller-Höcker et al. 1992; Bua et al. 2006; Greaves et al. 2014). mtDNA also undergoes ‘strict replication’, which, although not tied directly with the cell cycle as in the replication of nuclear DNA, nevertheless occurs to facilitate cell division, for example during embryogenesis, and which includes random segregation of mitochondrial genomes into daughter cells. The relative contribution of these relaxed and strict replication processes to mtDNA population dynamics, via the total number of replications experienced by an individual mtDNA molecule, is unknown (reviewed in (Lawless et al. 2020)). The accumulation of pathogenic variants to a high level in some cells results in a mosaic pattern of respiratory complex deficiency; assuming no significant changes in mtDNA copy number, normal respiratory chain function is maintained up to a certain threshold proportion or level of pathogenic variant, above which substantial respiratory complex deficiency occurs (Rossignol et al. 2003). Studies in *trans*mitochondrial cybrid cells lines containing different proportions of mutant mtDNA demonstrated the existence of this threshold, which differs between variants (Wallace 1986; Hayashi et al. 1991; Chomyn et al. 1992), however, variability also occurs between cell lines indicating that nuclear background may modify the cellular response to variant level (Dunbar et al. 1995). Furthermore, the genetically unstable and mitotic nature of these cells means they may not be representative of the post-mitotic patient tissues which are most frequently affected by mitochondrial disease (Sasarman et al. 2008a; Gorman et al. 2016).

Early investigations into threshold effects of the pathogenic m.3243A>G variant in patient biopsies used sequential cytochrome *c* oxidase (COX)-succinate dehydrogenase (SDH) histochemistry to identify focal fibres demonstrating biochemical defects in COX; consistent with stochastic segregation and expansion of mtDNA variants, they showed a mosaic pattern of deficiency, with higher m.3243A>G levels in COX-deficient and ragged red fibres (RRFs) compared to COX-positive fibres (Tokunaga et al. 1994; Petruzzella et al. 1994; Ozawa et al. 1998; Kärppä et al. 2005). However, only a small proportion of fibres from m.3243A>G patients tend to be RRF and/or COX-deficient (Moraes et al. 1992; Petruzzella et al. 1994; Kärppä et al. 2005; Jeppesen et al. 2006), and in some patients, no obvious COX histochemical deficiency is observed in skeletal muscle (Moraes et al. 1992). Biochemical studies have shown that complex I activity is preferentially affected in muscle from patients with m.3243A>G (Goto et al. 1992; Moraes et al. 1992; Hammans et al. 1992; Morgan-Hughes et al. 1995; James et al. 1996; Fornuskova et al. 2008), affirmed by a recently-described quadruple immunofluorescence assay which has shown that complex I deficiency appears to precede complex IV deficiency in muscle fibres from m.3243A>G carriers (Rocha et al. 2015; Ahmed et al. 2017).

Considering the effect on complex I and the phenotypic heterogeneity associated with m.3243A>G, we wanted to compare mutation load distributions in normal and deficient fibres. An obvious step is to determine whether threshold varies between patients. We explored patterns of both complex I and complex IV deficiency in patient muscle to further characterise the complexity associated with this variant and used our single-fibre observations to estimate the threshold levels of m.3243A>G above which respiratory deficiency occurs (**Figure 1**). As these thresholds cannot be directly observed in patient tissue; mechanistic models (Henderson et al. 2009) or statistical models (Rocha et al. 2018) can be used to estimate them. Here we progress the statistical approach (Rocha et al. 2018), using a richer dataset and a more robust statistical analysis to identify deficient fibres: we use unsupervised machine learning to characterise the respiratory complex status of muscle fibres, allowing us to investigate the distribution of m.3243A>G levels within hundreds of “normal” and “deficient” single fibres. We model these m.3243A>G level distributions using kernel density estimates, generate bootstrap estimates of the m.3243A>G threshold at which fibres develop a respiratory complex defect and compare estimated threshold distributions between patients. We expected that any significant between-individual variation that we might observe could explain some of the heterogeneity in individual outcomes for m.3243A>G-related disease and provide clues for individualised approaches to treatment.

**Figure 1:**
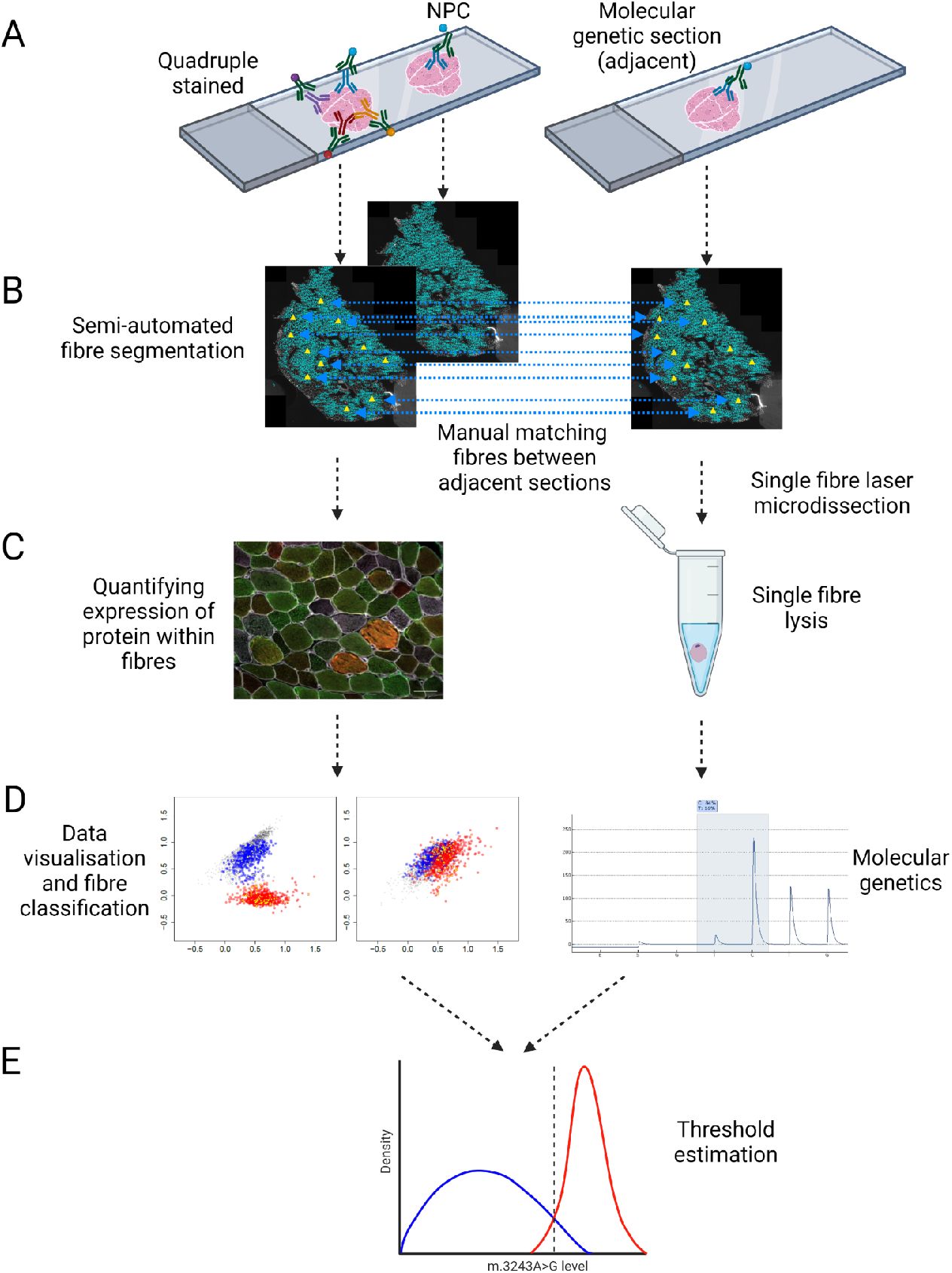
Schematic diagram of experimental method linking single cell protein expression and genetics in skeletal muscle sections. A: Quantitative quadruple immunohistochemistry was performed on serial 10µm transverse skeletal muscle sections labelled with antibodies detecting subunits of mitochondrial Complex I (NDUFB8), Complex IV (COX-1), a mitochondrial mass marker (mitochondrial porin; VDAC1) and laminin (membrane protein). A no-primary control section (NPC), labelled only for laminin, was processed in parallel; a serial 20µm section was also cut to enable molecular genetics to be performed. B: Fibre boundaries were defined using semi-automated fibre segmentation and manually matched across serial sections to enable downstream integration of protein quantification and molecular genetic assays. C: Protein expression was quantified using fluorescent imaging and selected fibres were isolated using laser microdissection of the serial 20µm skeletal muscle section. D: The OXPHOS status of muscle fibres was characterised by unsupervised machine learning; m.3243A>G levels in isolated single fibres were quantified by pyrosequencing. E: Distributions of the m.3243A>G level within ‘normal’ and ‘OXPHOS deficient’ fibres were modelled using kernel density estimates generating estimates of the m.3243A>G threshold at which fibres develop an OXPHOS defect.

## RESULTS

### Complex I deficiency predominates over Complex IV deficiency

We visualised protein expression patterns for key structural subunits of mitochondrial complex I (NDUFB8; CI) and complex IV (COX-1; CIV) in individual skeletal muscle fibres using 2Dmito plots (Warren et al. 2020), **Figure 2** and **Supplementary figures**) and used unsupervised machine learning (GMM clustering) to identify clusters of fibres within individual muscle sections. This approach allowed us to define muscle fibres from a single patient as either belonging to a respiratory complex “deficient” or “normal” cluster, based on relative distance of protein expression profiles from those in control subjects. We thereby avoided the difficult problem of accounting for batch effects or inter-individual variability in protein expression in the control population when classifying respiratory complex status of individual fibres. We find that there is considerable variability in protein expression profiles between healthy controls. In previous analysis (Rocha et al. 2015; Warren et al. 2020), the only way we could account for such effects was by increasing the number of control subjects, which involves considerable extra cost and difficulty recruiting subjects.

**Figure 2:**
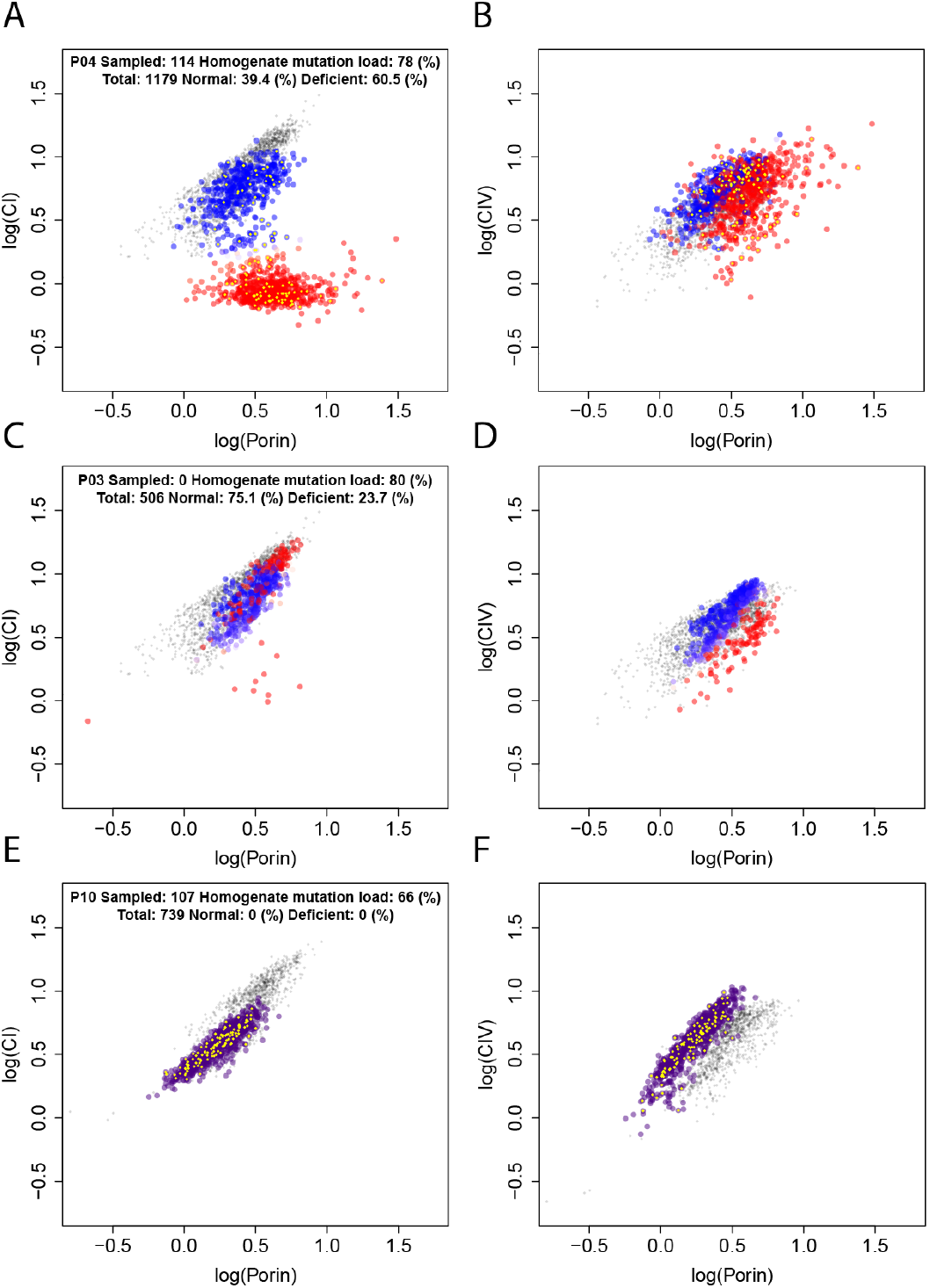
2Dmito plots show diverse Complex I, Complex IV and porin protein expression profiles. Each fibre is represented by one point in both Complex I (CI; left) and Complex IV (CIV; right) panels and is coloured according to its Gaussian Mixture Model classification (grey – unclassified fibres from five non-disease, control individuals, blue - respiratory chain normal fibres from a patient, red - respiratory chain deficient fibres from a patient, purple – unclassified fibres from a patient: only one class identified). Examples from three m.3243A>G positive individuals showing common patterns are shown: **A & B** – P04: CI expression increases with porin in OXPHOS normal fibres but not in OXPHOS deficient fibres. CIV expression is similar in both groups; OXPHOS deficient fibres have higher porin levels without a corresponding increase in CIV levels. **C & D** - P03: Only a very small number of fibres have low CI levels; some fibres with low CIV levels have normal CI levels. **E & F** - P10: All fibres in the same class and appear to show slightly reduced CI and porin expression and slightly increased CIV expression relative to controls.

In fibres from 15 of the 17 patients examined, we identified two distinct clusters. We classified the cluster that was most similar to controls as displaying normal CI and CIV expression relative to porin (blue points; **Figure 2 a-d**) and those that were least similar to controls as defective for either CI or CIV or both (red points; **Figure 2 a-d**). For the majority of patients, CI deficiency predominated over CIV deficiency; 2Dmito plots show a v-shaped pattern of CI expression with two distinct clusters, whereas CIV deficiency is not as clearly defined and appears to affect fewer fibres. In two of the muscle sections examined, GMM clustering identified only one cluster of fibres (P05 and P10, **Figure 2 e-f** and **Supplementary figures**).

### Patterns of single fibre respiratory complex deficiency vary greatly between individuals

Although fibres from the majority of muscle sections displayed a similar pattern of CI deficiency, the relative patterns of CI, CIV and porin expression varied greatly between patients (**Supplementary figures, Table 2**). We identified some broad patterns in the 2Dmito plots we generated, some of which appear in the same individuals.

For the majority of patients (exemplified by P04, **Figure 2 a-b**), the 2Dmito plots for CI show two distinct clusters of fibres (of varying proportions). In the normal cluster, CI expression increases as porin increases. In the deficient cluster CI expression does not increase as porin increases. However, there is no such strong split evident in CIV data. We also see other patterns in much smaller numbers of patients. In some we observe a shift to the right for the deficient fibres, indicating evidence of mitochondrial mass (porin) upregulation in the presence of respiratory complex deficiency (exemplified by P04, **Figure 2 a-b**). Some muscle sections show evidence of CIV deficiency in the absence of CI deficiency; most deficient fibres overlay the normal fibres in the CI 2Dmitoplot and CIV expression increases with porin at the same rate in deficient fibres as in normal fibres (exemplified by P3, **Figure 2 c-d)**. A small number of fibres from four patients (exemplified by P02, **Supplementary figures**) display reduced expression of CI, CIV and porin, forming a ‘tail’ in the bottom left corner of the 2Dmito plots. In addition, we also noted fibres that we have defined as normal for P02 have CI, CIV and porin comparable to the lowest values from controls, though this could be due to incomplete sampling of the natural variation in the healthy control population. Fibres from P06 also display a unique pattern (**Supplementary figures)**; there are two distinct clusters but there is also a general down-shift in the levels of CI, CIV and porin and in this patient, the two clusters segregate in both CI and CIV 2Dmito plots.

In fibres from two patients (P05, P10), we could only identify one cluster. Because our methodology relies on defining fibres as either normal or deficient based on the presence of two clusters, we chose to define the status of these single clusters as unknown, nevertheless protein expression patterns were very different. For P05, CI expression was substantially lower than controls with all fibres clustering below the control fibres, whilst porin expression was higher and CIV expression similar compared to controls. In contrast, for P10, expression of both CI and porin was lower and CIV higher than controls. It is possible that these apparent shifts are due to incomplete sampling of the natural variation in the healthy control population.

### The proportion of deficient fibres varies between individuals

We identified a sub-population of deficient fibres in the muscle sections of 15 out of 17 patients. The proportion of deficient fibres in patients with a deficient sub-population ranged from 3.6 to 64.3% (**Table 2**). We found no significant association between the proportion of deficient fibres and homogenate m.3243A>G level, the number of fibres analysed, total scaled NMDAS score (linear regression; *p* = 0.090, 0.130 and 0.548 respectively) or myopathy (Wilcoxon rank sum test; p = 0.155). However, we detected a significant negative correlation with age at biopsy (R^2^ = 0.37, p = 0.009, slope = −1.20, SE = 0.40). This is likely to reflect a selection bias; only a small proportion of m.3243A>G carriers who come to clinic undergo a skeletal muscle biopsy and these are likely to be younger, more clinically affected patients.

### Homogenate m.3243A>G level corresponds to the mean of single cell estimates, but is a poor representation of underlying distribution in single fibres

As m.3243A>G level is known to vary considerably within tissues, next we explored the distribution of m.3243A>G levels in individual muscle fibres. We selected ten individual skeletal muscle sections; eight with varying proportions of deficient fibres (range = 3.6% − 60.5%, median = 26.8%, IQR = 39.6%; P01, P02, P04, P07, P10, P12, P14, P15, P17) and two sections containing only one cluster (P5, P10) and quantified m.3243A>G variant levels in a sample including normal and deficient fibres (range of number of fibres sampled = 89 - 118, median = 106.5, IQR = 9.5).

Single-fibre m.3243A>G level showed considerable within-section variability in all muscle sections (**Supplementary file S1**). In four cases, (P02, P05, P14 and exemplified by P10, **Figure 3c and d**), the m.3243A>G level distribution is relatively narrow and the median sits close to the tissue homogenate m.3243A>G variant level. In contrast, m.3243A>G variant level distributions in fibres from six other cases (P01, P07, P12, P15, P17 and exemplified by P04, **Figure 3a and b**) are much wider and are bimodal, with higher, narrower distributions in the deficient fibres. The complexity of these distributions and the observation that they often span almost the full range of possible values (0-100%) confirms that tissue homogenate m.3243A>G level is a poor summary of the broad and complex distributions of m.3243A>G level found in cells from individual fibres.

**Figure 3:**
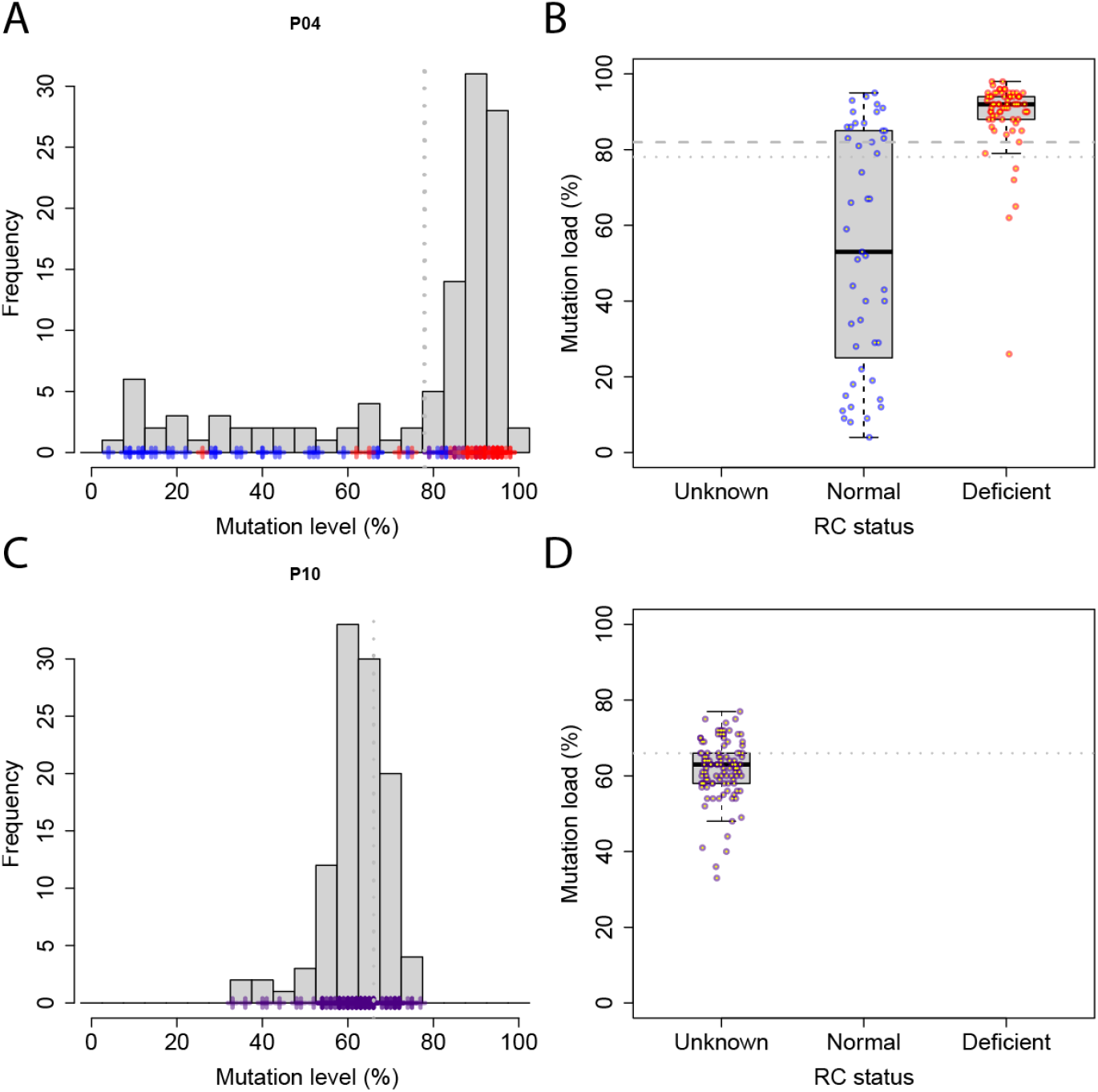
m.3243A>G variant level distribution in single skeletal muscle fibres from P04 (a & b) and P10 (c & d). **A & C**: frequency distribution of m.3243A>G levels; single-fibre observations are marked with crosses, coloured according to OXPHOS status. Vertical grey dotted line = tissue homogenate m.3243A>G level. **B & D**: m.3243A>G level distribution where fibres are split by OXPHOS class: blue = OXPHOS normal fibres, red = OXPHOS deficient fibres, purple = OXPHOS status unknown, grey dotted line = tissue homogenate m.3243A>G level, grey dashed line = mean threshold estimate.

### Tissue segregation and expansion of m.3243A>G determines muscle histochemistry profile

We observed that the six patients whose muscle sections show wider, bimodal distributions of single-fibre m.3243A>G level display a different pattern of deficient fibres compared to those from four patients whose muscle sections have narrower, unimodal distributions (**Table 2**). Wider distributions, more evident in normal fibres (**Figure 4**), are associated with a higher proportion of deficient fibres (range = 8.2% - 60.5%, median = 36.1%, IQR = 29.9%). Furthermore, m.3243A>G levels in deficient fibres are higher and more tightly distributed than levels in normal fibres. In two of the sections displaying narrower distributions, the proportions of deficient fibres are much lower (P02: 3.6% and P14:6.9%) and only one cluster of fibres was identified in the other two (P05 and P10). These observations suggest that cellular mechanisms responsible for the differential segregation and expansion of the pathogenic m.3243A>G variant differ between individuals and that these processes are a major driver of OXPHOS deficiency in single muscle fibres.

**Figure 4:**
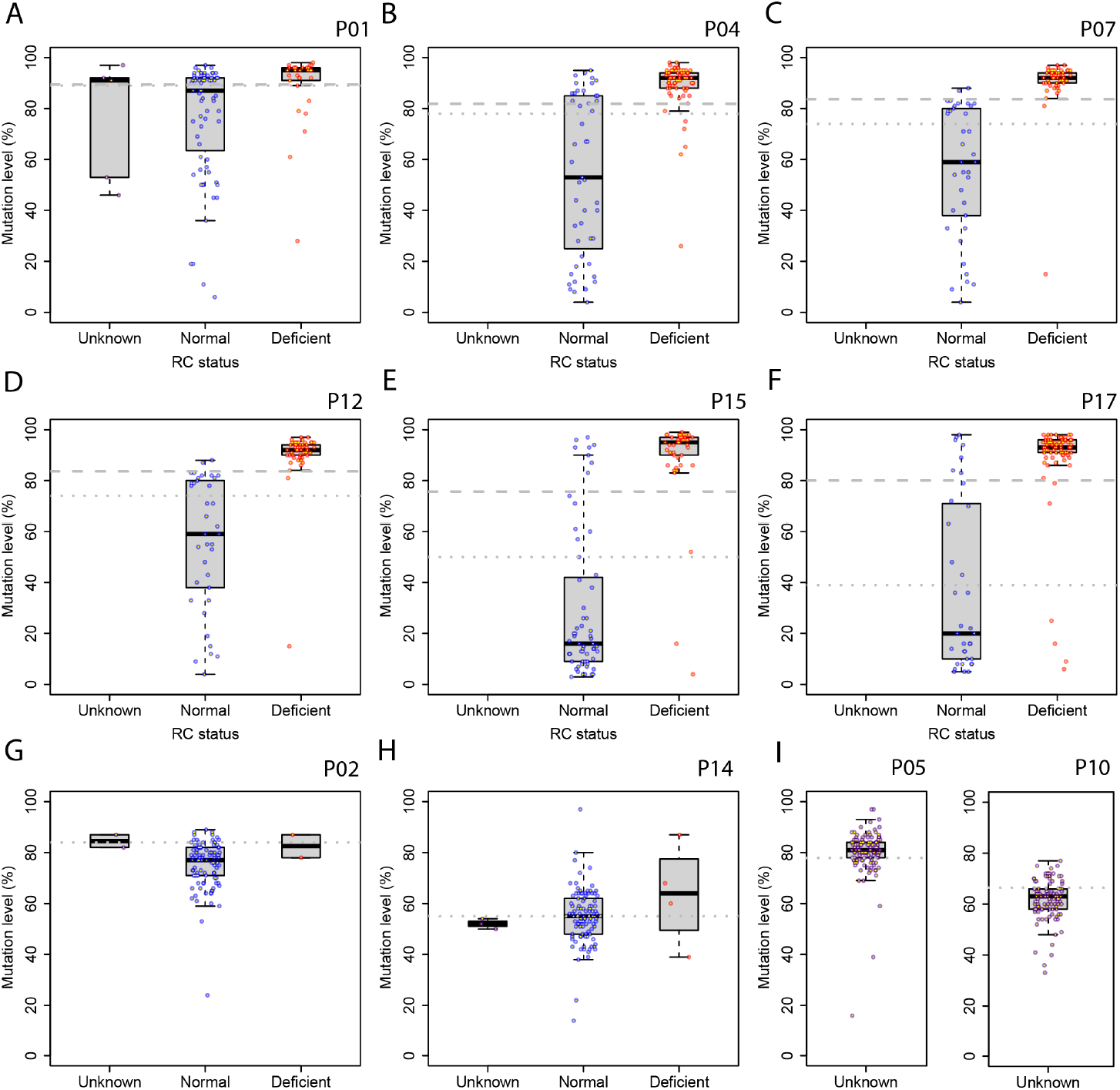
m.3243A>G level distribution in muscle fibres split by OXPHOS status. Blue = OXPHOS normal fibres, red = OXPHOS deficient fibres, purple = OXPHOS status unknown, grey dotted line = tissue homogenate m.3243A>G level, grey dashed line = mean threshold estimate. Box plots show the median and IQR, whiskers represent minimum and maximum values within 1.5 x IQR. A-F example patients where m.3243A>G level distribution is very broad. G & H example patients where m.3243A>G level distribution is relatively narrow. I example patients where m.3243A>G level distribution is narrow and only one cluster is detected from biochemical data.

### We find no difference in the threshold of m.3243A>G required for deficiency between patients

Having observed that the level of m.3243A>G variant is higher in deficient fibres, and given the hypothesis that cells can tolerate m.3243A>G levels up to a threshold with little or no functional consequence, next we aimed to determine the critical level of m.3243A>G; the level at which respiratory complex deficiency becomes detectable. For the six muscle sections with at least 10 deficient fibres, we generated bootstrap estimates of this threshold (**Figure 5, Table S2**);.

**Figure 5:**
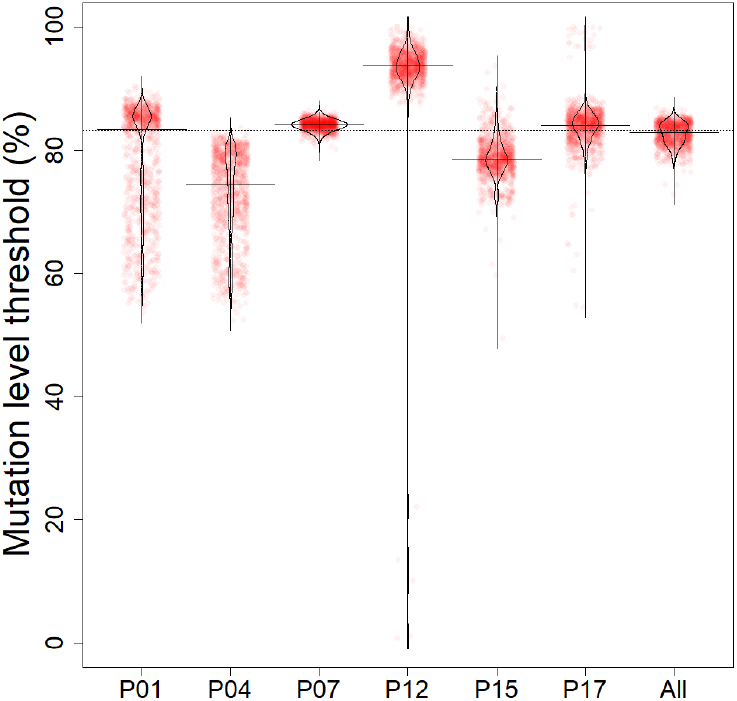
Estimated m.3243A>G level thresholds for patients carrying m.3243A>G variant. 10,000 bootstrapped samples from each patient representing uncertainty about the m.3243A>G level threshold above which biochemical deficiency occurs (red points) along with kernel density estimates of bootstrapped samples (black curves). Horizontal line segments represent median threshold estimates. Dotted line represents the median threshold across all patients. The All column represents uncertainty about the mean level across all six patients for which estimates are possible.

Pairwise comparisons of threshold estimates using the permutation test allowed us to determine the magnitude and significance of these differences. We find that none of the 15 possible pairwise comparisons have a difference in threshold estimates that is statistically significant (p>0.05).

We find no significant association between higher thresholds and a lower disease burden (scaled NMDAS), however, the statistical power of our study to detect such an association is low (slope = −1.28, SE = 1.42, p = 0.4188). We also had insufficient power to detect any significant association with age, m.3243A>G level in tissue homogenate or the proportion of deficient fibres.

We combine data from all six patients for which we can estimate a threshold to give an overall estimate of the threshold at which respiratory chain defect occurs. We do this by calculating, for each of the 10,000 bootstrap estimates, the median across all six patients. We find that the overall median threshold estimate is 82.9% with IQR between 81.5% and 84.1%.

## DISCUSSION

The underlying cellular mechanisms that contribute to the vast phenotypic heterogeneity observed in individuals with the pathogenic m.3243A>G variant are poorly understood. In an effort to characterise this variation within skeletal muscle and to estimate threshold mutation levels, we quantified mitochondrial respiratory complex I and IV expression in single skeletal muscle fibres from 17 patients, revealing mosaic patterns of biochemical deficiency that primarily affected complex I protein expression. This is something that previous studies, which only evaluated COX (CIV) activity, were unable to detect. Using unsupervised clustering to classify fibres, we saw striking variation in both the proportion of respiratory complex “deficient” myofibres and the pattern of deficiency between patients. This between-individual variability confirms the utility of skeletal muscle, which is relatively accessible, in understanding the different mechanisms underlying the variable phenotypic expression of m.3243A>G in post-mitotic tissue.

In order to study the effect of the proportion of pathogenic mtDNA variants on respiratory complex defects, we also quantified m.3243A>G levels in individual muscle fibres from ten patients, revealing substantial intra- and inter-individual diversity not captured by the established approach of measuring the level of m.3243A>G in tissue homogenate. We found that wider, bimodal distributions of m.3243A>G level are associated with a higher proportion of respiratory-deficient fibres, whereas samples containing lower proportions of deficiency show a tighter distribution. Differential segregation of pathogenic variant and wild-type mtDNA molecules into myofibres during the mitotic development of skeletal muscle may contribute to these differences, but variability in cellular response to the presence of the m.3243A>G pathogenic variant and subsequent clonal expansion could also play a role (Battersby et al. 2003; Lawless et al. 2020).

Consistent with previous studies, we found higher and more tightly distributed m.3243A>G levels in respiratory complex “deficient” compared to “normal” fibres, and by modelling these distributions have been able to generate bootstrap estimates of the m.3243A>G level threshold at which respiratory complex deficiency begins for six patients. These thresholds are relatively high, but not significantly different from each other. Our median estimate for the threshold is 82.9% with IQR between 81.5% and 84.1%, confirming the functionally recessive nature of this mtDNA variant. Although previous studies have shown higher mutation levels in COX-deficient fibres (Moraes et al. 1992; Tokunaga et al. 1994; Petruzzella et al. 1994; Ozawa et al. 1998; Kärppä et al. 2005; Jeppesen et al. 2006), this is the first time that critical thresholds of respiratory complex deficiency have been estimated using m.3243A>G level distributions. Extending this observation to a larger number of patients may give us some insight into the differences we see in disease severity and progression between patients with similar homogenate m.3243A>G levels (Grady et al. 2018). It is likely to be influenced by factors such as nuclear background, which may modulate the cellular response to the presence of m.3243A>G (Pickett et al. 2018). It would be fascinating to determine whether there is also between-individual variability in tissue-specific critical thresholds and whether this contributes to the diversity of m.3243A>G-related phenotypes that is also largely unexplained (Kaufmann et al. 2011; de Laat et al. 2012; Nesbitt et al. 2013; Mancuso et al. 2014; Fornuskova et al. 2008).

Currently it is thought that much of the heterogeneity that we see in the mutation level in single cells sampled from post-mitotic tissues from patients arises due to clonal expansion during relaxed mtDNA replication throughout the patient’s lifespan. However, heterogeneity arising as a result of variability in the “initial condition” of this process: heterogeneity generated by clonal expansion during very rapid, strict mtDNA replication during development, is largely ignored, but might contribute (Lawless et al. 2020). Determining the relative contribution of these two processes in human muscle tissue remains a challenge.

Studies of patient skeletal muscle biopsies are cross-sectional in nature, showing only a single time point for each fibre, and with no single-cell m.3243A>G level data available for early stages of foetal development, it is difficult to assess the relative contributions of differential segregation and clonal expansion to the heteroplasmic mosaicism and between-individual variability in m.3243A>G level distributions. Previous work has shown that homogenate m.3243A>G levels in the early developing foetus are similar across tissues, which suggests that the segregation of mutant and wild-type mitochondrial genomes into daughter cells during tissue development may be random and unrelated to respiratory function (Monnot et al. 2011). In contrast, tissue-specific nuclear genetic control of mtDNA segregation has been demonstrated in mice carrying two very different mitochondrial genomes (Battersby et al. 2003); whether a mechanism that could differentiate human mitochondrial genomes based on a single point mutation is yet to be demonstrated, although we do know that the bottleneck in oogenesis is influenced by both nuclear and mitochondrial genetics (Wilson et al. 2016; Pickett et al. 2019), suggesting that differential segregation could be shaped by an active mechanism that shows variability between individuals.

Random drift by relaxed replication is thought to be sufficient to explain the clonal expansion of mitochondrial point mutations in post-mitotic tissues (Chinnery and Samuels 1999; Elson et al. 2001; Coller et al. 2001), however, the striking difference in the distribution of single-fibre m.3243A>G levels in muscle sections displaying a higher degree of respiratory complex deficiency compared to those with relatively low levels of deficiency suggests that some selective pressure may take place. Recent work focusing on mtDNA deletions suggests that the initiation of clonal expansion occurs in the perinuclear niche of skeletal myofibres, providing more evidence that clonal expansion is likely to be under the control of the nucleus and thus may be influenced by nuclear genetic background (Vincent et al. 2018). If this is also the case for point mutations such as m.3243A>G, it could give us clues as to the cellular control mechanisms that contribute to clonal expansion in muscle, age-related pathology and the vast phenotypic variation associated with this variant, which is likely to be influenced by nuclear factors (Maeda et al. 2016; Pickett et al. 2018).

The predominance of complex I deficiency in m.3243A>G muscle has been well-documented (Goto et al. 1992; Moraes et al. 1992; Hammans et al. 1992; Morgan-Hughes et al. 1995; James et al. 1996; Fornuskova et al. 2008), and hypothesised to be due to the impact of m.3243A>G on the synthesis of ND6 and ND5 subunits as a result of their high UUG codon content (Chomyn et al. 2000; Dunbar et al. 1996; Kirino et al. 2004; Sasarman et al. 2008b). Importantly, we show that the patterns of complex I and complex IV deficiency are very different between individuals, suggesting that the m.3243A>G pathomechanism is not necessarily universal. Theories proposed to explain the pathogenicity of m.3243A>G include the mis-incorporation of amino acids into mtDNA-encoded proteins (Yasukawa et al. 2000; Flierl et al. 1997; Sasarman et al. 2008b), impairment of the termination of mt-rRNA transcription (Hess et al. 1991; King et al. 1992) and a decrease in steady-state levels of aminoacylated mt-tRNA(Leu[UUR]) (Janssen et al. 1999; Yasukawa et al. 2000; Chomyn et al. 2000). Given the between-individual variability in focal respiratory deficiency observed, it would seem likely that the lack of consensus could, in part, be related to the impact of other (nuclear) genetic factors.

Ultimately, it would be fascinating to be able to replace quadruple immunofluorescence with Imaging Mass Cytometry (IMC) - examining many more respiratory complex proteins and signalling molecules, but the real strength of the approach that we present here is the combined quantification of protein expression and variant level in single cells. Other important extensions would be to observe the variation in respiratory complex defect (as in (Elson et al. 2002)) and m.3243A>G level along the length of muscle fibres, and the variation in both between sections. Both would give different insights into the spatial variation of the processes of clonal expansion and the loss of respiratory complex function, which is currently largely unknown.

The statistical analyses presented here suggest that there could be a great deal of heterogeneity in the timing and strength of clonal expansion and the effect this has on mitochondrial function. These factors are likely to contribute to the diverse phenotypic presentation of m.3243A>G, highlighting the importance of understanding these complex processes. As a single muscle fibre cannot be studied over multiple time points, moving from naive statistical models and clustering towards predictive, mechanistic, mathematical models of respiratory complex function and dysfunction as well as the processes leading to dysfunction might usefully bridge this gap. If we could predict the evolution of mutant and wild type genotype distribution during disease progression, it could explain some of the observed clinical heterogeneity. Adding even a simple, threshold model of the onset of focal biochemical deficiency in response to mtDNA mutant load could help us explore the link between focal respiratory deficiency and clonal expansion, shedding light on the phenotypic diversity observed between patients.

## CONCLUSIONS

Quantifying the diverse phenotypic consequences of the m.3243A>G variant in patient cells is an important first step towards understanding and ultimately treating patients carrying this common cause of mitochondrial disease. We have demonstrated a robust new method for classifying patient myofibres into respiratory normal and respiratory deficient sub-populations and describe distinct distributions of m.3243A>G allele frequency in the two sub-populations. The diversity of fibre phenotypes and genotype distributions which we quantified was surprising and will provide important constraints on possible mechanisms to describe the progression of disease in patients carrying the pathogenic m.3243A>G variant.

## METHODS

### Patient Cohort

We obtained skeletal muscle biopsies (Tibialis Anterior muscle or Quadriceps) from 17 patients (10 females) who were registered in the Mitochondrial Disease Patient Cohort: A Natural History Study and Patient Registry and Newcastle Mitochondrial Research Biobank (REC references 13/NE/0326 and 16/NE/0267) and who harbour the pathogenic m.3243A>G variant (**Table 1**). Patients were between 18 and 60 years at the time of skeletal muscle biopsy (mean = 43.6 years, SD = 12.6), their scaled NMDAS score (calculated as described previously (Grady et al. 2018)) ranged from 4.1 to 49.7 (mean = 24.5, SD = 11.8) and mean muscle homogenate m.3243A>G levels ranged from 39% to 89% (mean = 66.8, SD = 14.0). Muscle biopsy referrals, clinical evaluation and molecular genetics had previously been undertaken as part of the diagnostic pathway at the NHS Highly Specialised Service for Rare Mitochondrial Disorders of Adults and Children.

**Table 1.** Clinical and molecular genetic characteristics of patient cohort. Clinical phenotypes are derived from the NMDAS assessment closest to the time of muscle biopsy using NMDAS scores and previously defined thresholds (Pickett 2018); scaled total NMDAS are calculated as previously described (Grady et al 2018); differences in age between biopsy and NMDAS assessment ranged from 0 - 2.4 years (median = 0.40, IQR = 0.1). Homogenate m.3243A>G level refers to tissue homogenate m.3243A>G level measured in the skeletal muscle sample studied. CPEO - chronic progressive external ophthalmoplegia.

| Patient | Sex | Homogenate m.3243A>G Level (%) | Age at biopsy | Clinical Presentation | Total Scaled NMDAS |
| --- | --- | --- | --- | --- | --- |
| P01 | M | 89 | 21-25 | Seizures | 12.4 |
| P02 | F | 84 | 51-55 | Hearing impairment, diabetes | 20.7 |
| P03 | M | 80 | 46-50 | Migraine, myopathy | 20.4 |
| P04 | F | 78 | 31-35 | Hearing impairment, myopathy, cerebellar ataxia | 23.2 |
| P05 | F | 78 | 41-45 | Hearing impairment, diabetes, psychiatric involvement, myopathy | 27.9 |
| P06 | F | 77 | 21-25 | Seizures, stroke-like episodes, encephalopathy, myopathy, dysphonia/dysarthria | 33.5 |
| P07 | F | 74 | 46-50 | Migraine, ptosis, myopathy | 21.8 |
| P08 | F | 70 | 56-60 | Hearing impairment, diabetes | 26.9 |
| P09 | M | 67 | 46-50 | Hearing impairment, diabetes, cerebellar ataxia, myopathy | 45.1 |
| P10 | F | 66 | 16-20 | Migraine, myopathy | 21.5 |
| P11 | M | 63 | 31-35 | Mild phenotypes only (under threshold) | 4.1 |
| P12 | F | 62 | 56-60 | Mild phenotypes only (under threshold) | 9.7 |
| P13 | M | 57 | 36-40 | Cardiovascular involvement, ptosis, CPEO | 26.9 |
| P14 | F | 55 | 56-60 | Migraine, myopathy | 22.8 |
| P15 | M | 50 | 51-55 | Diabetes, cardiovascular involvement, ptosis, CPEO, myopathy, cerebellar ataxia, neuropathy | 35.2 |
| P16 | M | 46 | 41-45 | Hearing impairment, migraine | 14.0 |
| P17 | F | 39 | 55-60 | Hearing impairment, seizures, encephalopathy, CPEO, cerebellar ataxia | 49.7 |

**Table 2.** Summary of clustering patterns from 2Dmito plots. Complex I and Complex IV protein expression is compared with mitochondrial mass (porin expression). OXPHOS = oxidative phosphorylation, CI = Complex I, CIV = Complex IV, nd = not determined, Y = some evidence, N = no evidence.

| Patient | Number of fibres | % OXPHOS deficient fibres | No. clusters | Description | Separation between groups visible in plots for: |  | Higher porin expression in respiratory complex deficient fibres? | Fibres with lower CI, CIV and porin? | Number of fibres sampled for molecular genetics | Distribution of m.3243A>G level in OXPHOS normal fibres |
| --- | --- | --- | --- | --- | --- | --- | --- | --- | --- | --- |
|  |  |  |  |  | CI | CIV |  |  |  |  |
| P01 | 871 | 57.4 | 2 | Fibres low in CI present<br>Higher porin level in some OXPHOS deficient fibres | Y | 1 or 2 fibres only | Y | N | 106 | Wide |
| P02 | 101 | 3.6 | 2 | Small group of fibres with low expression of CI, CIV and porin | Y | 1 or 2 fibres only | N | Y | 101 | Narrow |
| P03 | 506 | 23.7 | 2 | Fibres low in CI and CIV present<br>Fibres low in CIV but not CI present | Y | Y | Slightly | N | 0 | nd |
| P04 | 1179 | 60.5 | 2 | Fibres low in CI and CIV present<br>Higher porin level in some OXPHOS deficient fibres | Y | Y | Y | N | 114 | Wide |
| P05 | 854 | nd | 1 | CI expression lower than controls<br>Porin expression higher than controls | N | N | N | N | 118 | Narrow |
| P06 | 857 | 64.3 | 2 | Fibres low in CI and CIV present<br>CIV expression lower than controls<br>Porin is not higher in OXPHOS deficient fibres<br>Porin expression lower than controls | Y | Y | N | Y | 0 | nd |
| P07 | 625 | 44.2 | 2 | Fibres low in CI and CIV present<br>Some deficient fibres have higher porin expression | Y | A few fibres only | Y | Y | 89 | Wide |
| P08 | 318 | 10.1 | 2 | Fibres low in CI and CIV present<br>Small group of fibres with low expression of CI, CIV and porin | Y | A few fibres only | N | Y | 0 | nd |
| P09 | 104 | 15.4 | 2 | Fibres low in CI present | Y | N | N | N | 0 | nd |
| P10 | 739 | nd | 1 | Downward-shift in CI and porin and upward-shift in CIV compared to controls | N | N | N | N | 107 | Narrow |
| P11 | 431 | 5.3 | 2 | Some CIV deficient fibres are normal for CI | 2 fibres only | Y | N | N | 0 | nd |
| P12 | 1261 | 8.2 | 2 | Fibres low in CI and CIV present | Y | Y | N | N | 89 | Wide |
| P13 | 422 | 12.3 | 2 | Fibres low in CI and CIV present | Y | A few fibres only | N | N | 0 | nd |
| P14 | 991 | 6.9 | 2 | Fibres low in CIV but not CI present | Y | Y | N | N | 108 | Narrow |
| P15 | 601 | 25.6 | 2 | Fibres low in CI and CIV present<br>Small group of fibres with low expression of CI, CIV and porin | Y | Y | N | Y | 106 | Wide |
| P16 | 290 | 6.9 | 2 | Fibres low in CI and CIV present<br>Fibres low in CIV but not CI present | Y | Y | N | N | 0 | nd |
| P17 | 1690 | 27.9 | 2 | Fibres low in CI and CIV present<br>Small group of fibres with low expression of CI, CIV and porin | Y | Y | Y | Y | 113 | Wide |

We obtained non-disease control skeletal muscle tissue (distal region of hamstring) from five healthy individuals (from the Newcastle Mitochondrial Research Biobank), following anterior cruciate ligament surgery (ages 16-20, 21-25, 21-25, 26-30 and 31-35 years; two females).

### Quantitative mitochondrial immunohistochemistry

We undertook quantitative quadruple immunohistochemistry on 10µm transverse skeletal muscle sections as previously described (3). Briefly, we labelled sections with antibodies (**Table S1**) detecting subunits of mitochondrial complex I (NDUFB8) and complex IV (COX-1), a mitochondrial mass marker (mitochondrial porin; VDAC1) and a cell membrane marker (laminin), followed by incubation with secondary antibodies (Alexa Fluor 488, 546, biotinylated IgG1 and 750) and subsequently with streptavidin 647. Alongside each sample, we processed a no-primary control section (labelled only for laminin). We captured fluorescent images of the muscle sections using automated scanning at 20× magnification using Zen 2011 (blue edition) software and Zeiss Axio imager MI microscope and maintained exposure times across all sections for each experimental batch.

We analysed the fluorescent images using an in-house analysis software Quadruple Immuno Analyser written in Matlab 2015a (*https://github.com/CnrLwlss/quad_immuno*), as previously described (Ahmed et al. 2017). This software automatically identifies single transverse myofibres in the images by creating surfaces over fibres using the laminin immunofluorescence (750 channel); segmentation was subsequently checked manually and adjusted where necessary. The mean pixel intensities of 488 (COX-1), 546 (Porin) and 647 (NDUFB8) in each individual fibre were measured for all fluorescent labelled muscle sections and for each of the no-primary control sections to determine the levels of non-specific binding. Following this, .csv files containing mean intensities for each muscle fibre were generated and used in subsequent statistical analyses.

### Single cell molecular genetics

We selected and isolated fibres for molecular genetic analysis using laser microdissection of serial 20µm skeletal muscle sections, as previously described (4) and quantified m.3243A>G levels in single muscle fibres using pyrosequencing on the Pyromark Q24 platform, as previously described (5). Primer sequences according to GenBank Accession number NC_012920.1 are: 5’biotinylated forward: m.3143-3163; reverse: m.3331-3353; reverse pyrosequencing primer: m.3244-3258 (IDT, Coralville, USA). We used the allele quantification application from Pyromark proprietary Q24 software to calculate m.3243A>G levels (test sensitivity > 3% mutant mtDNA, accuracy ± 3%).Three controls with known homogenate m.3243A>G levels were used to validate each experimental run.

### Statistical analysis

#### Classification of skeletal muscle fibres

To classify skeletal muscle fibres as either respiratory complex “normal” or “deficient”, we carry out two-component Gaussian Mixture Model (GMM) clustering of the multidimensional, mean protein expression levels estimated within each fibre by quadruple immunohistochemistry. We use GMM to classify patient fibres into exactly one or two clusters, using the `mclust` package (6) in R. For patients with two clusters, we randomly sample one fibre from each cluster and one fibre from controls from the same experimental batch and calculate the Euclidean distance in protein expression space between the two individual fibres and the control fibre. By repeating this random sampling procedure 15,000 times, we can calculate the proportion of times that a fibre from cluster #1 is closer to the control fibres than a fibre from cluster #2. If that proportion is > 0.5 then cluster #1 is classified as “normal” and cluster #2 is classified as “deficient”. Otherwise the opposite classification is used. For patients where one cluster is found, fibres are classified as “unknown”, however it is interesting to consider whether this cluster is distant from control fibres.

### Estimation of thresholds of biochemical deficiency

For patients with two clusters, in order to estimate the threshold m.3243A>G level at which a biochemical defect occurs, we identify those fibres in each cluster which have been sampled for genetic testing. If there are at least 10 sampled fibres in each of the two groups (normal and deficient), then we estimate the threshold m.3243A>G level (*T*) above which cells become respiratory complex deficient. To estimate *T*, we generate non-parametric kernel density estimates for the distribution of m.3243A>G levels in each of the two clusters, defining the threshold estimate as the proportion with the highest probability density where the density for the two clusters are equal. Bootstrap sampling the original data 10,000 times captures our uncertainty about the estimate.

### Demonstration of differences between estimated individual thresholds using bootstrapping

To examine whether differences that we observed between the threshold m.3243A>G level above which a respiratory complex defect occurs for each patient were reproducible, we test for significance using a non-parametric resampling method: the permutation test. We calculate, for each pair A & B taken from the set of patients where we can estimate a respiratory complex defect threshold, the absolute difference in their thresholds: *Δ*_*AB*_, as a test statistic. We then take the single cell data from patients A & B and shuffle the patient labels attached to each fibre and recalculate *Δ*_*AB*_. By repeating the last step 10,000 times, we build up the distribution of our two-tailed test statistic under the null hypothesis that there is no difference between patient A and B. We then calculate a p-value: the proportion of times that *Δ*_*AB*_ under H_0_ is greater than *Δ*_*AB*_ when the data are labelled correctly. We correct for multiple testing using Hochberg correction (`p.adjust` function in R).

## Supporting information

Supplemental Figures (all subjects)

## Data Availability

All data produced are available online at:
https://github.com/lwlss/mito_muscle_sections and https://dx.doi.org/10.25405/data.ncl.19416005

## DECLARATIONS

### Ethics approval and consent to participate

Mitochondrial disease patient tissue and ethical approval was provided by the Newcastle Mitochondrial Research Biobank (REC reference 16/NE/0267-Application Ref: MRBOC ID 016). Data for the study were obtained from the Wellcome Centre for Mitochondrial Research Patient Cohort (MitoCohort) (REC Ref: 13/NE/0326-Application Ref: MDOC ID 049).

### Consent for publication

Only anonymised samples and data were used in this research.

### Availability of data and materials

R code and data can be found in the GitHub repository for this article: *https://github.com/CnrLwlss/Ahmed_2022*. Quadruple Immuno Analyser is an open source Matlab 2015a program available in the GitHub repository (*https://github.com/CnrLwlss/quad_immuno*). Imaging files and fibre-matching images are available at: *https://dx.doi.org/10.25405/data.ncl.19416005*

Link to tables and figure legends: https://drive.google.com/file/d/18uG3oNOf8iJRFtu3-6uu-2l6VGDb1p19/view?usp=sharing Supplementary tables are included in Figure and Tables document.

Supplementary file S1 is a multi-page .pdf report showing 2Dmito plots and m.3243A>G level distributions (where available) for all patients in cohort:

https://drive.google.com/file/d/1pQMxsPziyHTS-KrKHIyWm7AnfISdUuru/view?usp=sharing

## Authors’ contributions

STA, RWT, DMT & SJP contributed to study design; STA performed laboratory experiments and collected the data; CL classified respiratory status of fibres, visualised fibre classifications, estimated and visualised thresholds; SJP carried out regression modelling against clinical data; SJP and CL wrote the initial draft of the manuscript; all authors commented on and approved the final version of the manuscript.

## Acknowledgements

We would like to thank the patients within our cohort for participating in this study, the clinical, laboratory and research administration and support teams within the Rare Mitochondrial Disorders of Adults and Children Service in Newcastle upon Tyne. We are grateful to Gavin Falkous and Amy Vincent for technical advice, Alasdair Blain for statistical advice and Robert McFarland for clinical advice.

---

**Table S1:** Primary and secondary antibodies used in the quadruple Immunofluorescence assay

| Antibody | Target | Supplier | Catalogue number | Dilution |
| --- | --- | --- | --- | --- |
| **Primary antibody** |  |  |  |  |
| Mouse IgG1 NDUFB8 | Subunit of Complex I | Abcam | Ab110242 | 1:100 |
| Mouse IgG2a MTCO1 | Subunit of Complex IIV | Abcam | Ab14705 | 1:100 |
| Mouse IgG2b VDAC (Porin) | Voltage gated anion on outer membrane of mitochondria | Abcam | Ab14734 | 1:100 |
| Polyclonal Rabbit IgG Laminin α-1 | Protein of the extracellular matrix | Sigma Aldrich | L9393 | 1:50 |
| **Secondary antibody** |  |  |  |  |
| Goat Anti- mouse IgG1 (488nm) | N/A | Life Technologies | A21121 | 1:200 |
| Goat Anti- mouse IgG2b (546nm) | N/A | Life Technologies | A21143 | 1:200 |
| Goat Anti-rabbit IgG (750nm) | N/A | Life Technologies | A21039 | 1:100 |
| Streptavidin (647nm) | N/A | Life Technologies | S32357 | 1:100 |
| Goat Anti-mouse IgG1 Biotin | N/A | Life Technologies | A10519 | 1:200 |

**Table S2:** Estimates of critical m.3243A>G threshold estimates from 10,000 bootstrapped samples.

| Patient | Median | IQR |
| --- | --- | --- |
| P01 | 82.68 | 16.05 |
| P04 | 74.60 | 10.55 |
| P07 | 84.14 | 1.11 |
| P12 | 93.53 | 2.82 |
| P15 | 78.78 | 3.67 |
| P17 | 84.10 | 2.98 |

File S1: **Multi-page .pdf report showing 2Dmito plots and m.3243A>G level distributions (where available) for all patients in cohort**.

https://drive.google.com/file/d/1pQMxsPziyHTS-KrKHIyWm7AnfISdUuru/view?usp=sharing

