## Supplemental Figures (all subjects) for "Quantifying phenotype and genotype distributions in single muscle fibres from patients carrying the pathogenic mtDNA variant m.3243A>G"

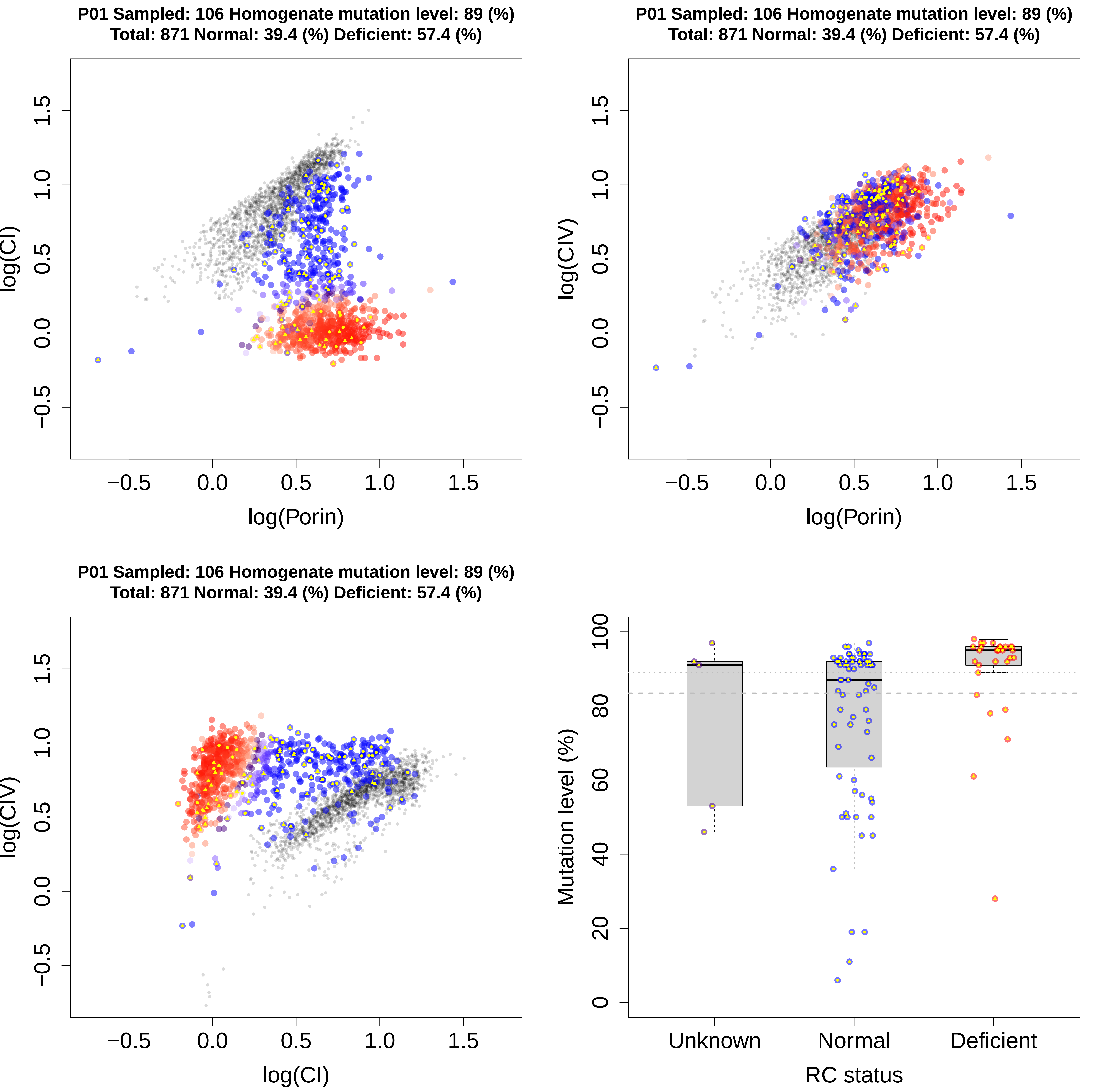
P01 Sampled: 106 Homogenate mutation level: 89 (%)
Total: 871 Normal: 39.4 (%) Deficient: 57.4 (%)
log(CI)
log(Porin)
P01 Sampled: 106 Homogenate mutation level: 89 (%)
Total: 871 Normal: 39.4 (%) Deficient: 57.4 (%)
log(CIV)
log(Porin)
P01 Sampled: 106 Homogenate mutation level: 89 (%)
Total: 871 Normal: 39.4 (%) Deficient: 57.4 (%)
log(CIV)
log(CI)
Mutation level (%)
Unknown
Normal
Deficient
RC status

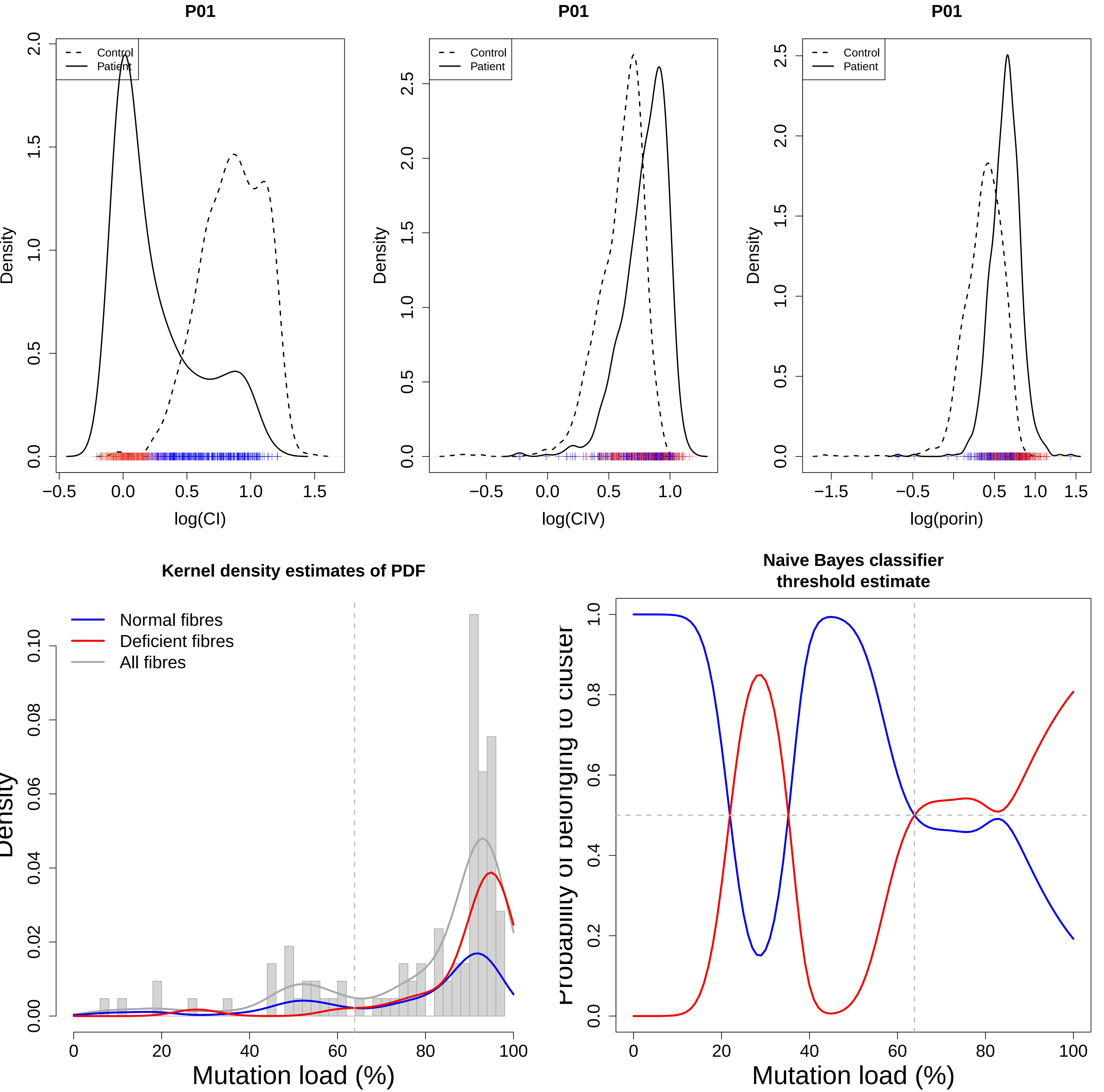
P01
Control
Patient
Density
log(CI)
P01
Control
Patient
Density
log(CIV)
P01
Control
Patient
Density
log(porin)
Kernel density estimates of PDF
Normal fibres
Deficient fibres
All fibres
Density
Mutation load (%)
Naive Bayes classifier
threshold estimate
Probability of belonging to cluster
Mutation load (%)

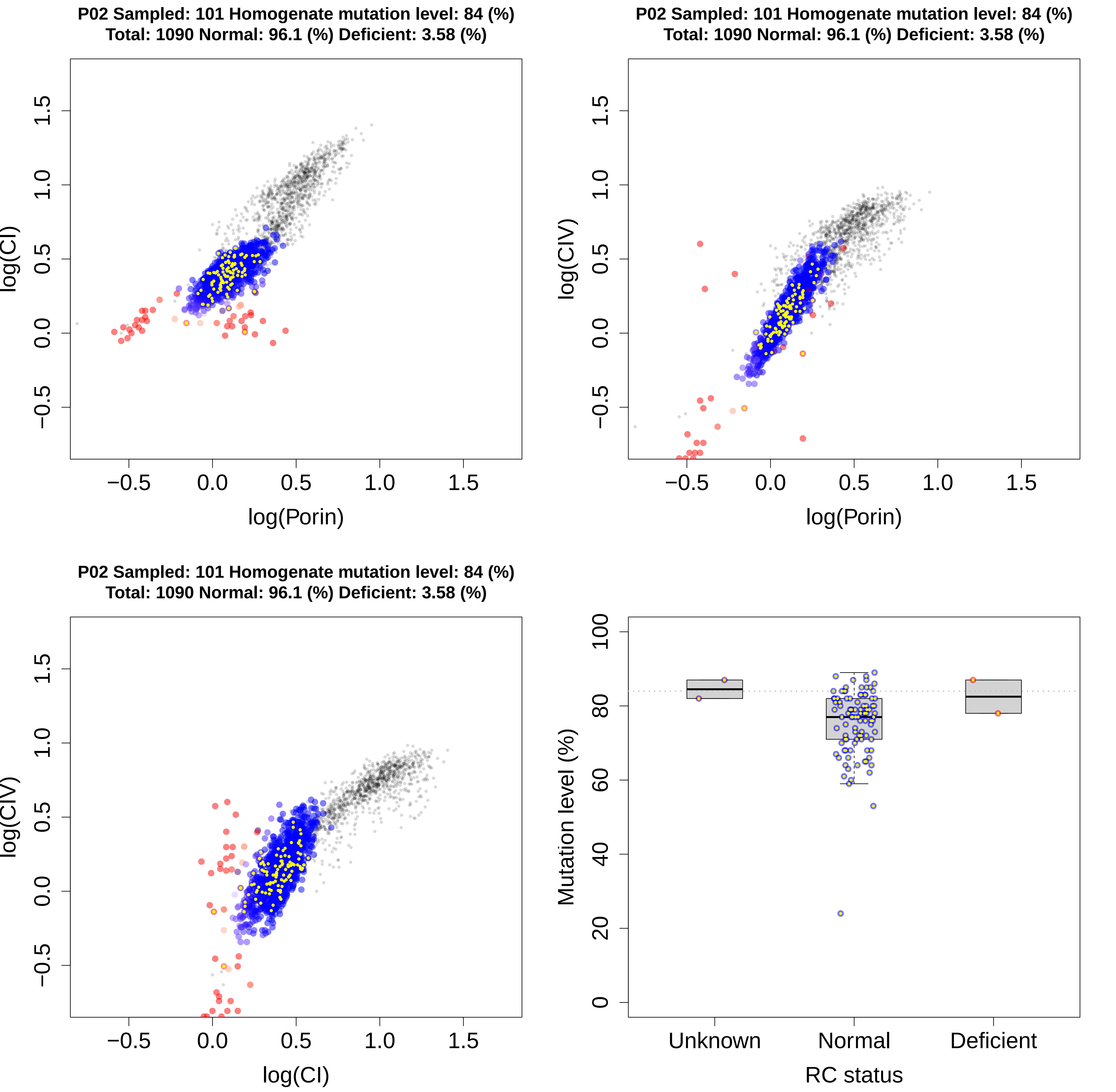

P02 Sampled: 101 Homogenate mutation level: 84 (%)
Total: 1090 Normal: 96.1 (%) Deficient: 3.58 (%)
log(CI)
log(Porin)
−0.5
0.0
0.5
1.0
1.5
P02 Sampled: 101 Homogenate mutation level: 84 (%)
Total: 1090 Normal: 96.1 (%) Deficient: 3.58 (%)
log(CIV)
log(Porin)
P02 Sampled: 101 Homogenate mutation level: 84 (%)
Total: 1090 Normal: 96.1 (%) Deficient: 3.58 (%)
log(CIV)
log(CI)
Mutation level (%)
0
20
40
60
80
100
Unknown
Normal
Deficient
RC status

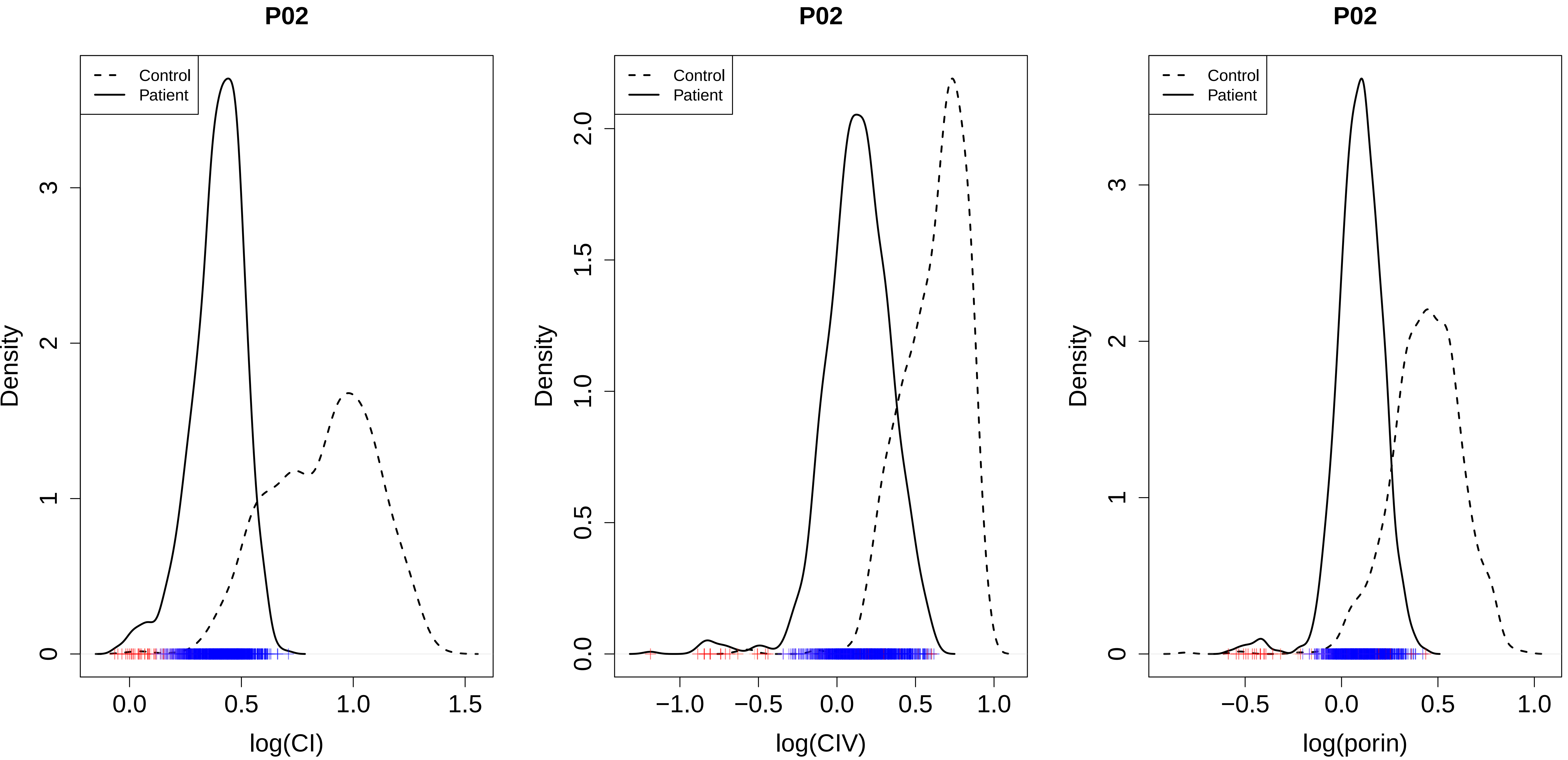

P02
Control
Patient
Density
log(CI)
P02
Control
Patient
Density
log(CIV)
P02
Control
Patient
Density
log(porin)

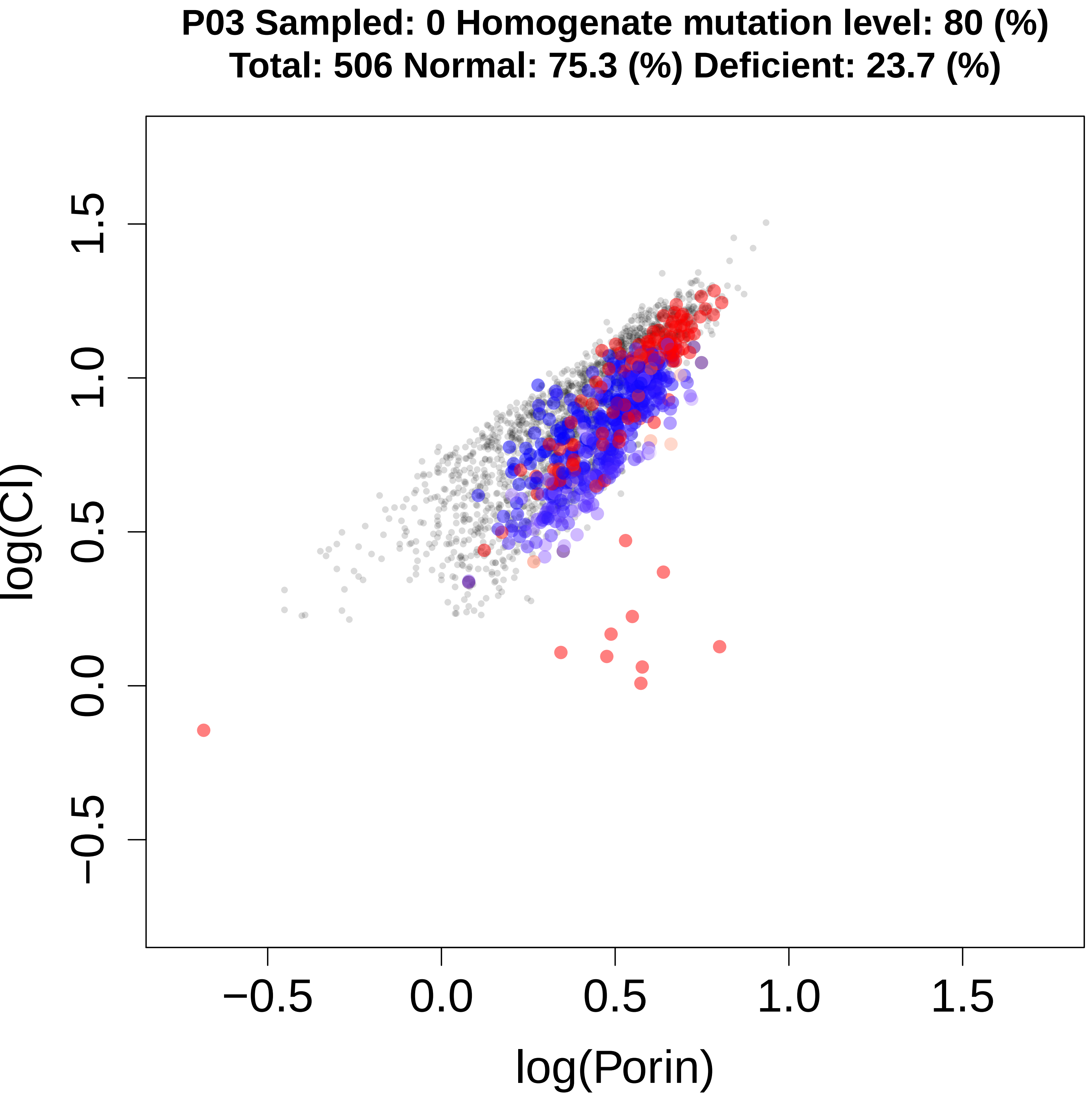
P03 Sampled: 0 Homogenate mutation level: 80 (%)
Total: 506 Normal: 75.3 (%) Deficient: 23.7 (%)
log(CI)
log(Porin)
−0.5
0.0
0.5
1.0
1.5

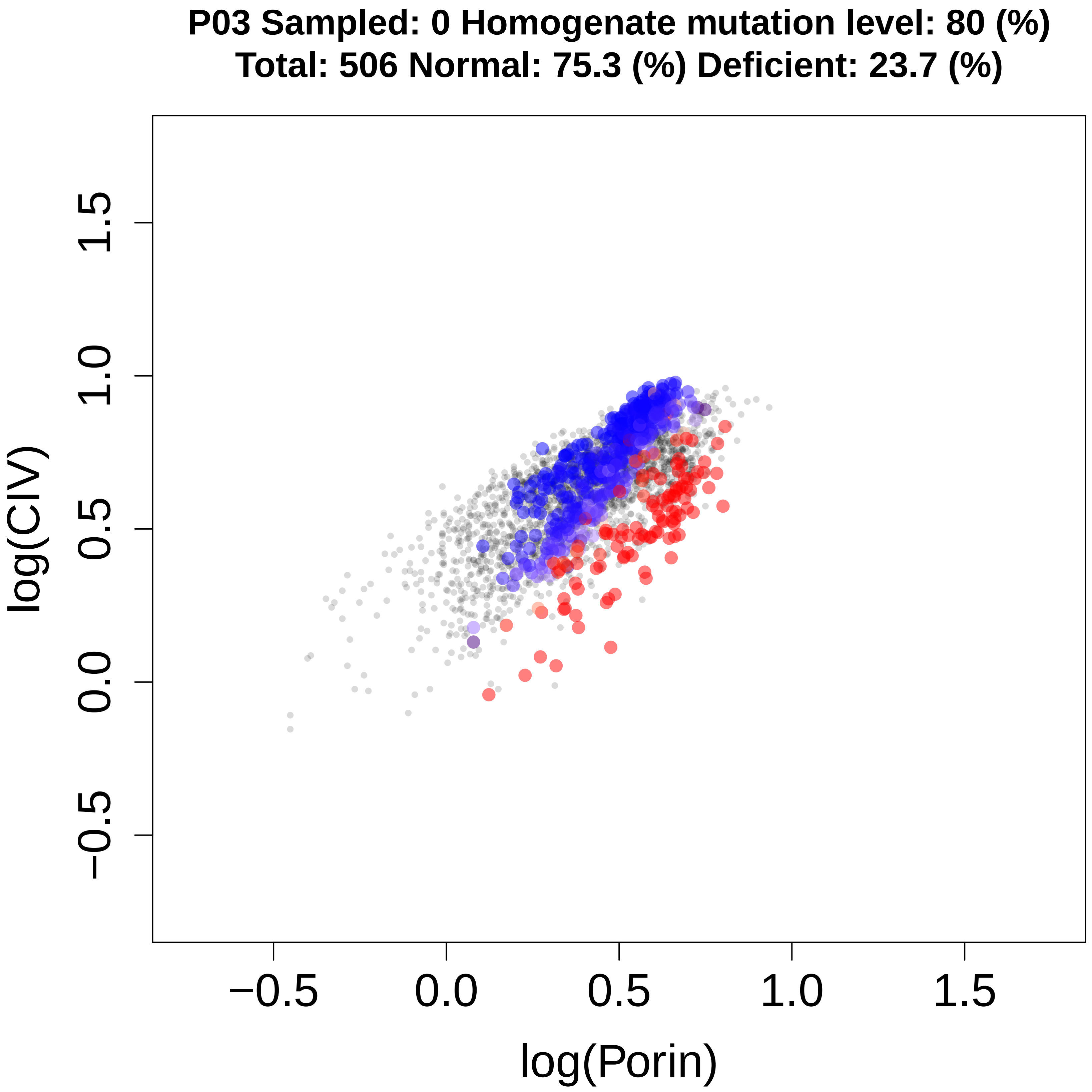
P03 Sampled: 0 Homogenate mutation level: 80 (%)
Total: 506 Normal: 75.3 (%) Deficient: 23.7 (%)
log(CIV)
log(Porin)
−0.5
0.0
0.5
1.0
1.5

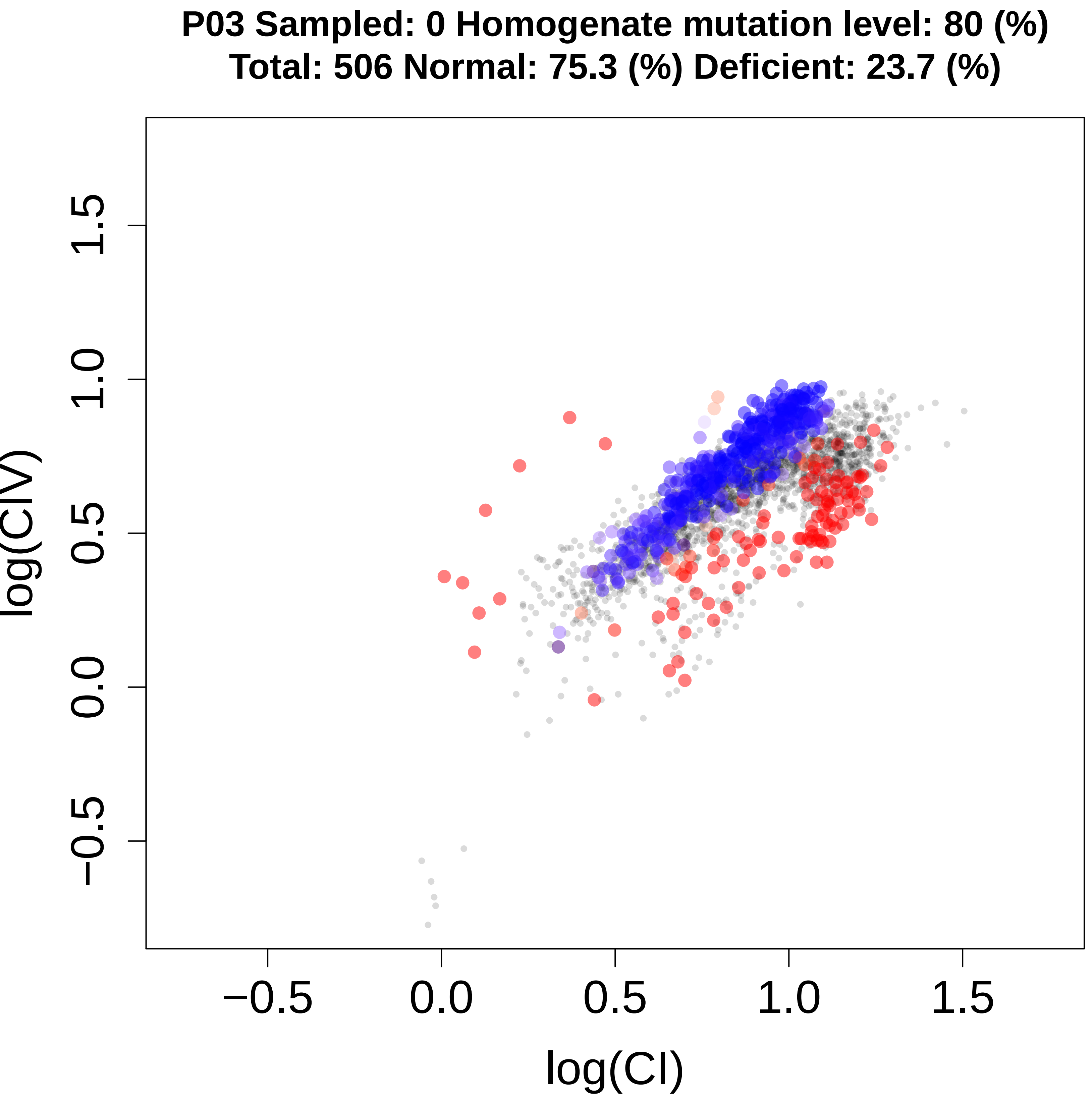
P03 Sampled: 0 Homogenate mutation level: 80 (%)
Total: 506 Normal: 75.3 (%) Deficient: 23.7 (%)
log(CIV)
log(CI)
−0.5
0.0
0.5
1.0
1.5

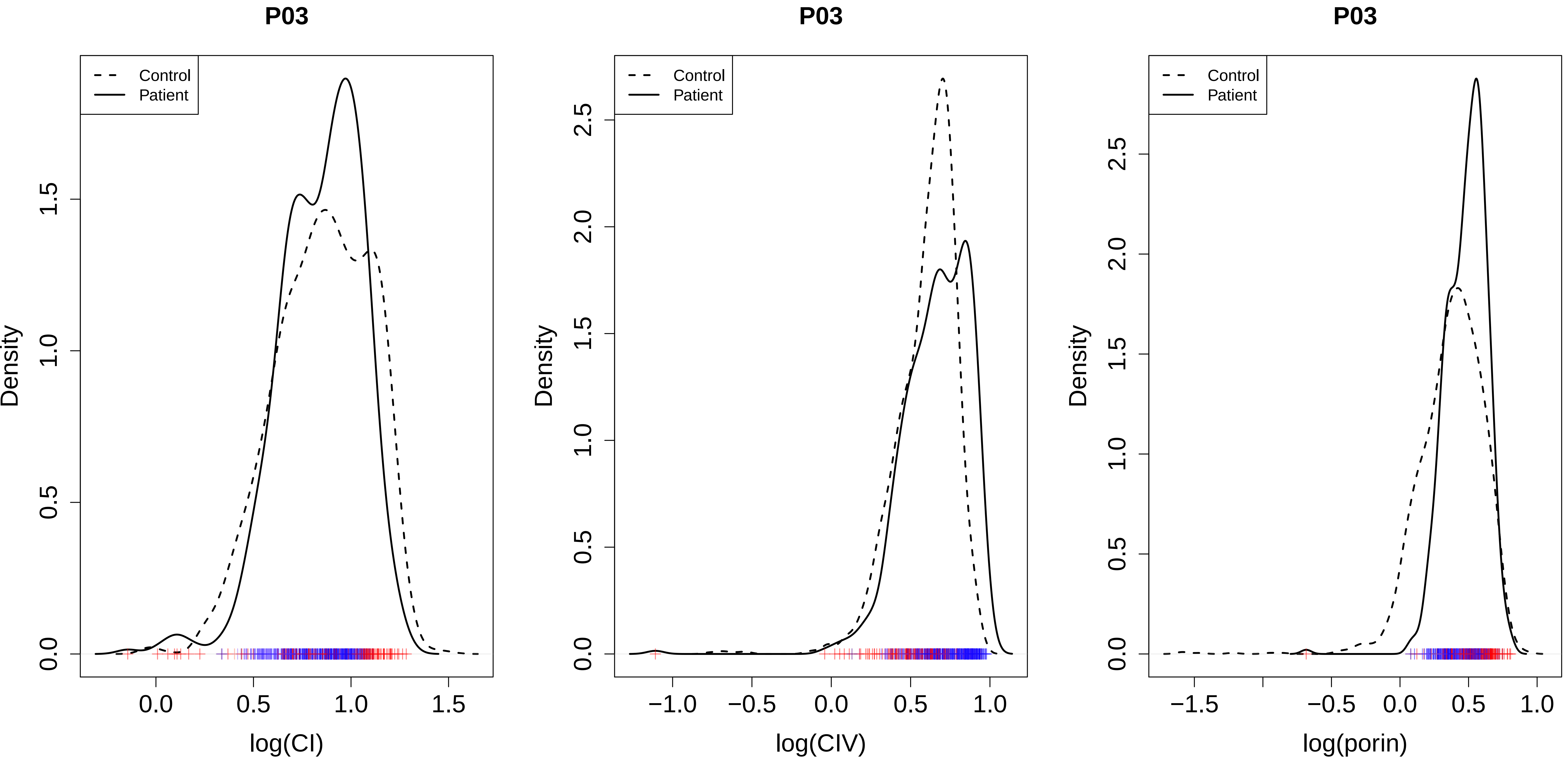
P03
Control
Patient
Density
log(CI)
P03
Control
Patient
Density
log(CIV)
P03
Control
Patient
Density
log(porin)

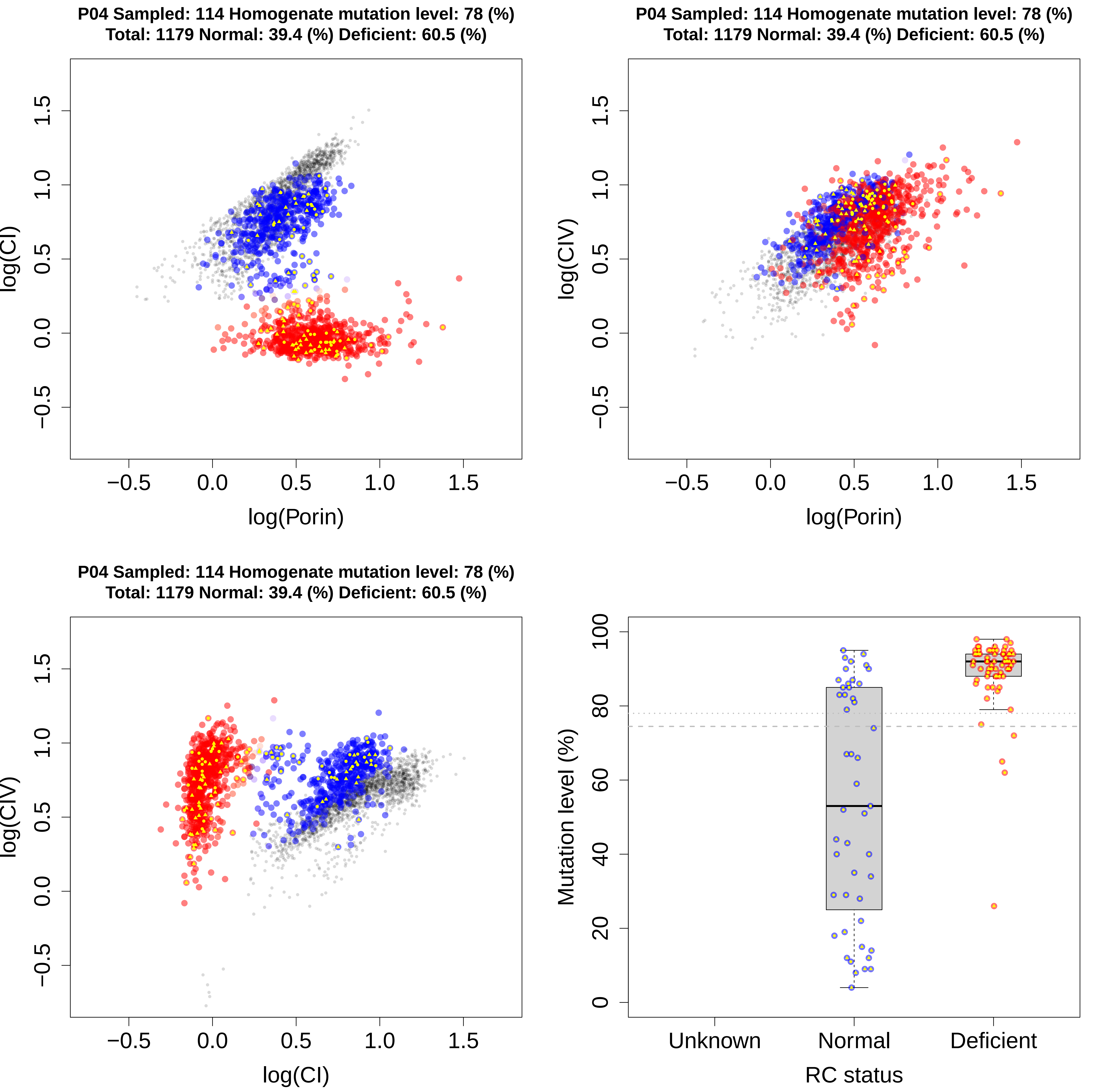

P04 Sampled: 114 Homogenate mutation level: 78 (%)
Total: 1179 Normal: 39.4 (%) Deficient: 60.5 (%)
log(CI)
log(Porin)
log(CIV)
log(Porin)
log(CIV)
log(CI)
Mutation level (%)
Unknown
Normal
Deficient
RC status

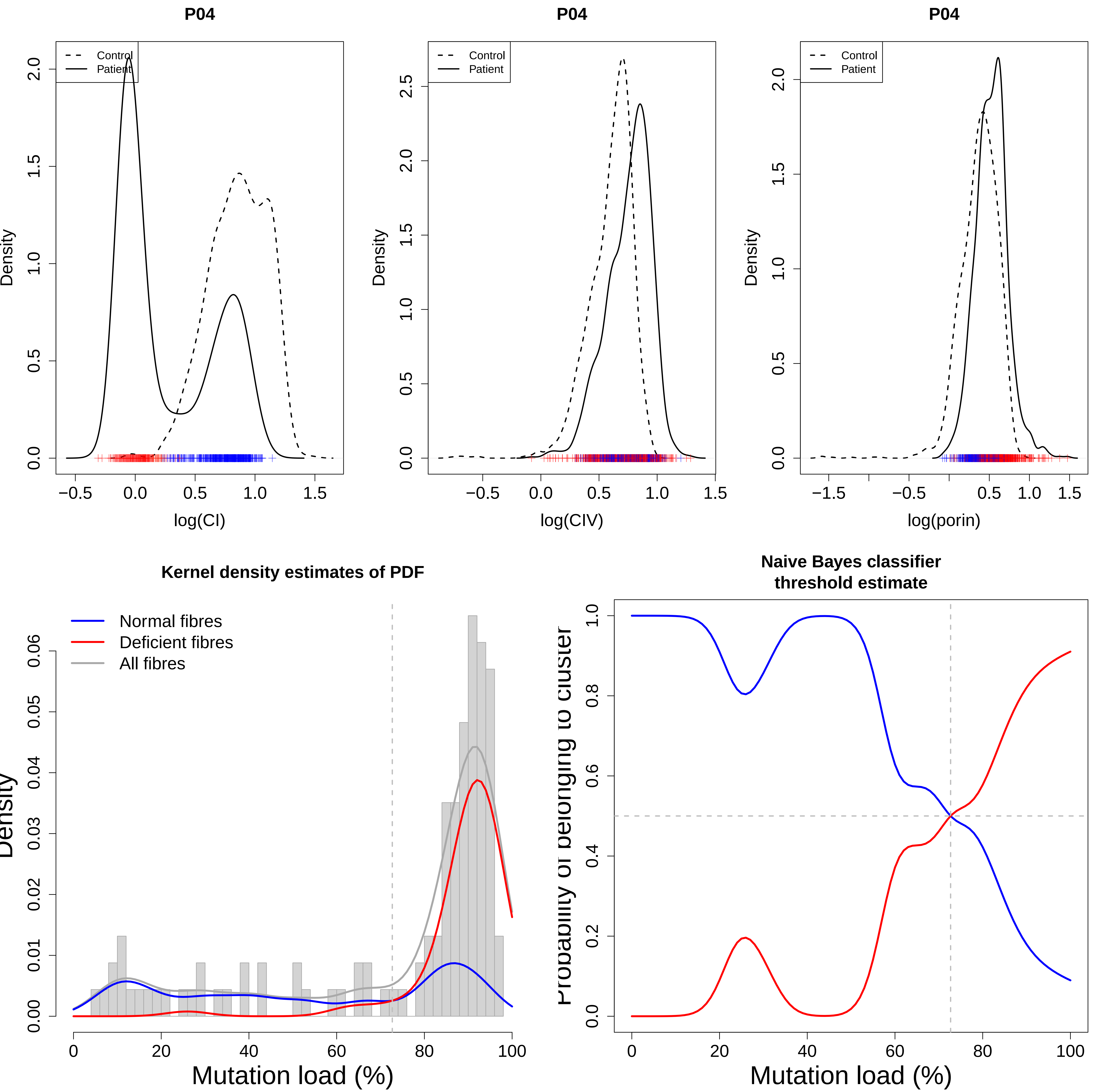

P04
Control
Patient
Density
log(CI)
P04
Control
Patient
Density
log(CIV)
P04
Control
Patient
Density
log(porin)
Kernel density estimates of PDF
Normal fibres
Deficient fibres
All fibres
Mutation load (%)
Naive Bayes classifier
threshold estimate
Mutation load (%)

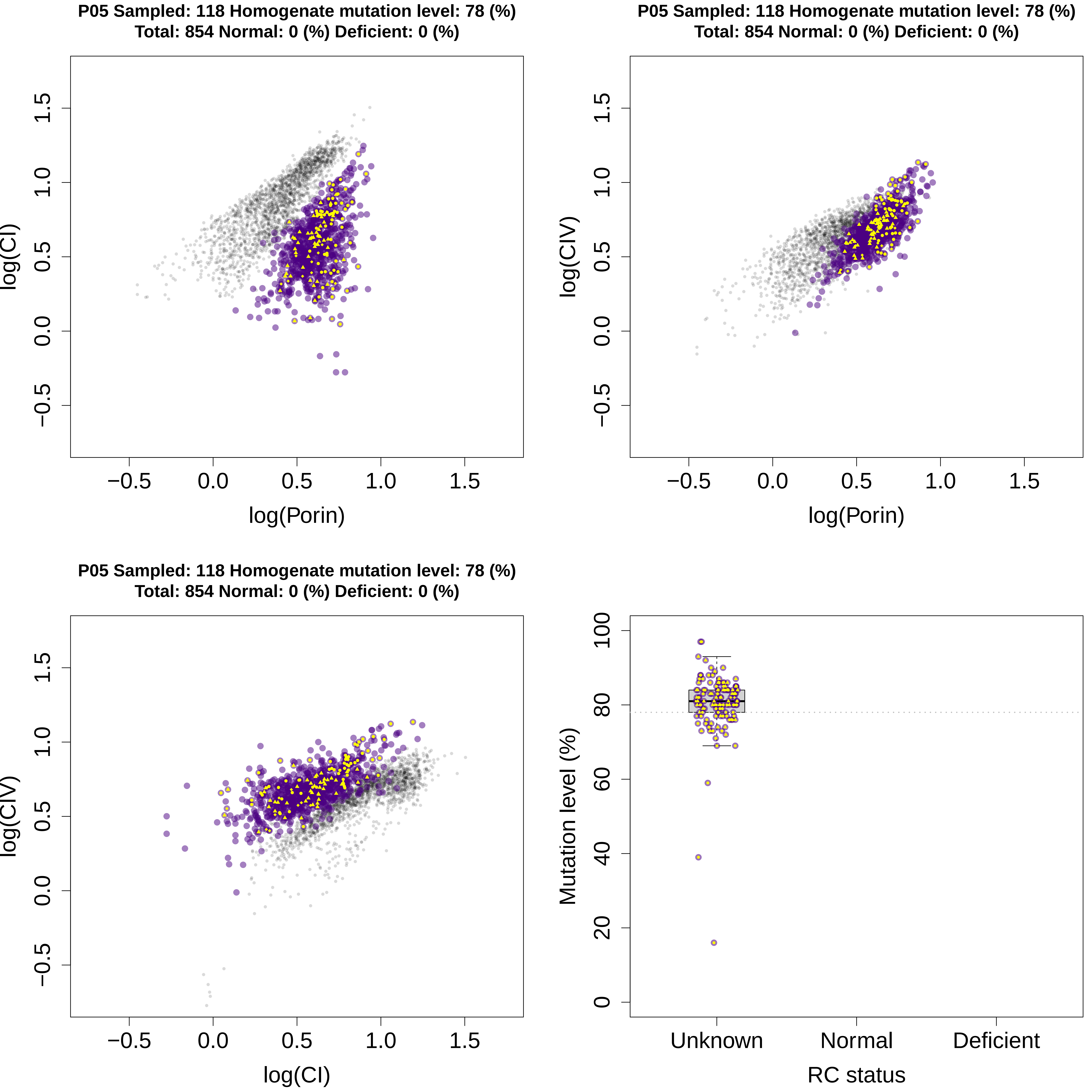
P05 Sampled: 118 Homogenate mutation level: 78 (%)
Total: 854 Normal: 0 (%) Deficient: 0 (%)
log(CI)
log(Porin)
P05 Sampled: 118 Homogenate mutation level: 78 (%)
Total: 854 Normal: 0 (%) Deficient: 0 (%)
log(CIV)
log(Porin)
P05 Sampled: 118 Homogenate mutation level: 78 (%)
Total: 854 Normal: 0 (%) Deficient: 0 (%)
log(CIV)
log(CI)
−0.5
0.0
0.5
1.0
1.5
Mutation level (%)
0
20
40
60
80
100
Unknown
Normal
Deficient
RC status

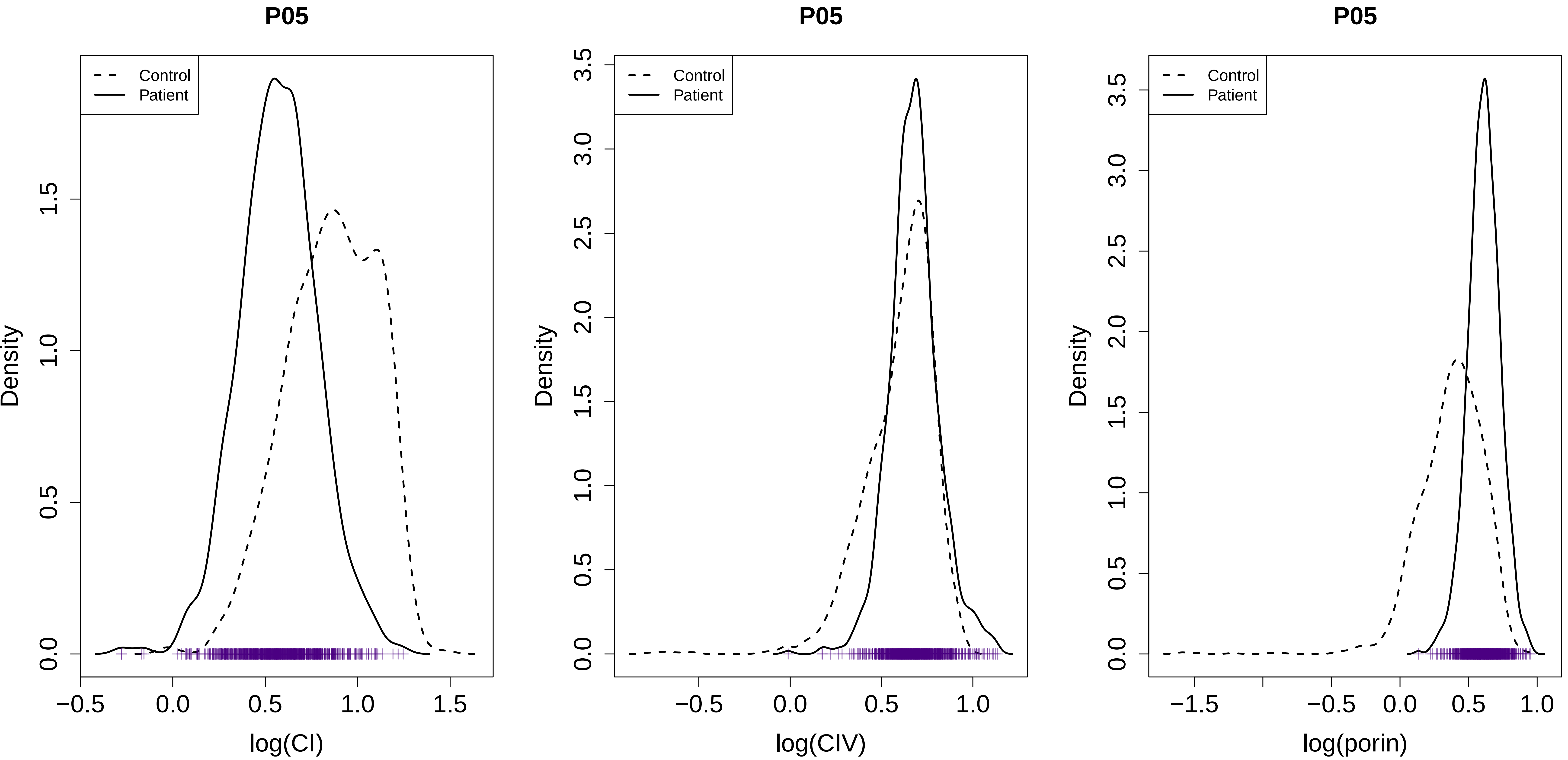
P05
Control
Patient
Density
log(CI)
P05
Control
Patient
Density
log(CIV)
P05
Control
Patient
Density
log(porin)

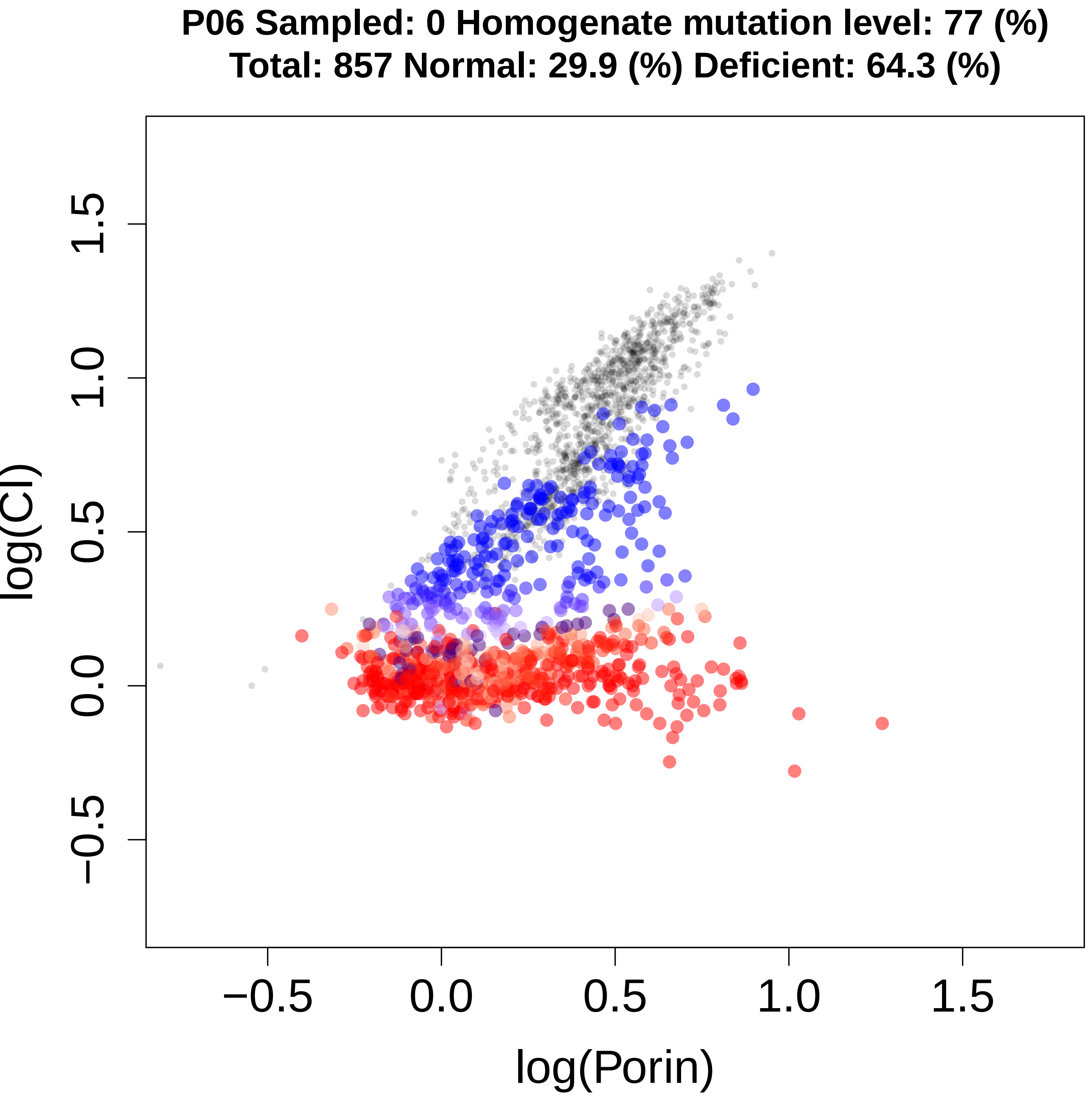
P06 Sampled: 0 Homogenate mutation level: 77 (%)
Total: 857 Normal: 29.9 (%) Deficient: 64.3 (%)
log(CI)
log(Porin)
−0.5
0.0
0.5
1.0
1.5

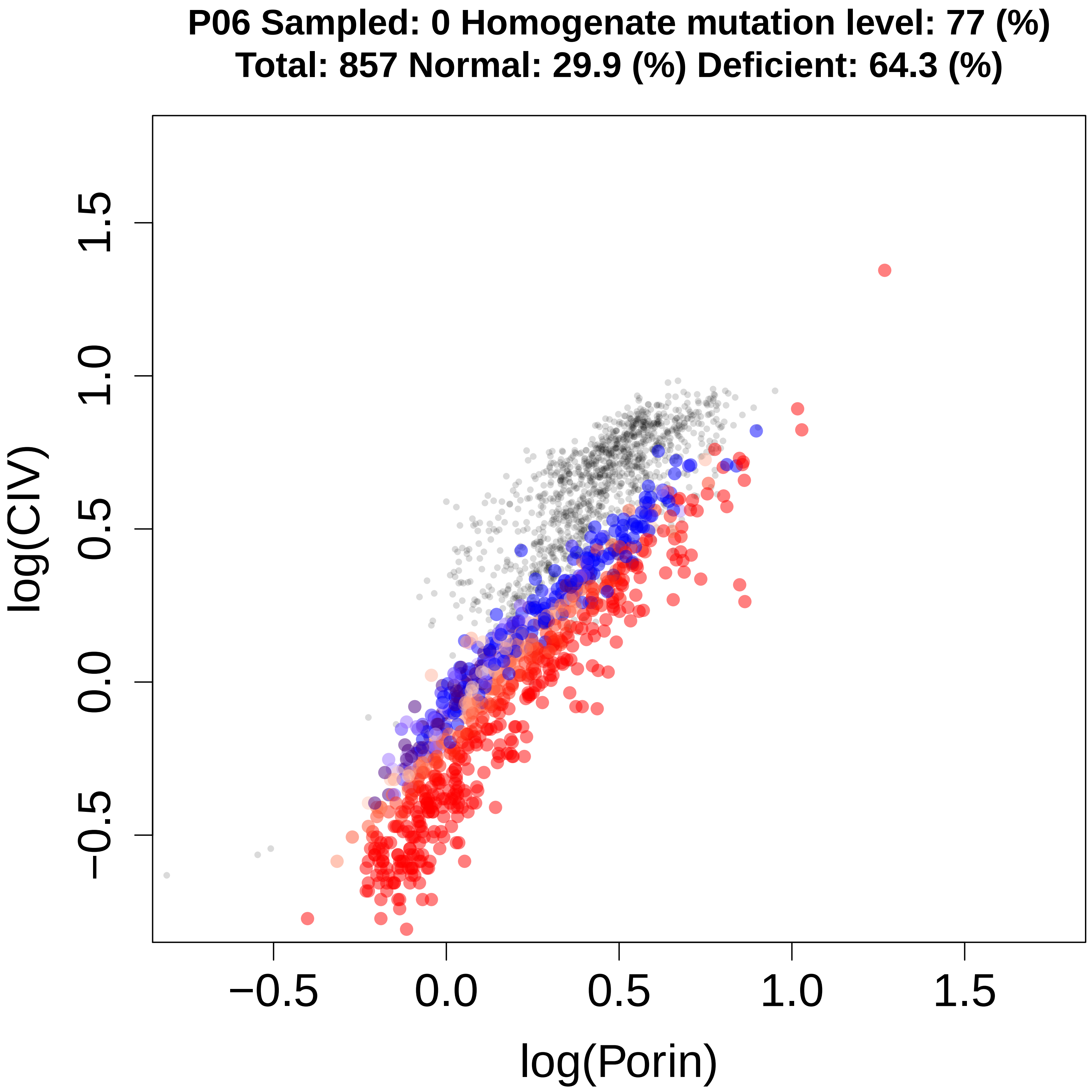
P06 Sampled: 0 Homogenate mutation level: 77 (%)
Total: 857 Normal: 29.9 (%) Deficient: 64.3 (%)
log(CIV)
log(Porin)
−0.5
0.0
0.5
1.0
1.5

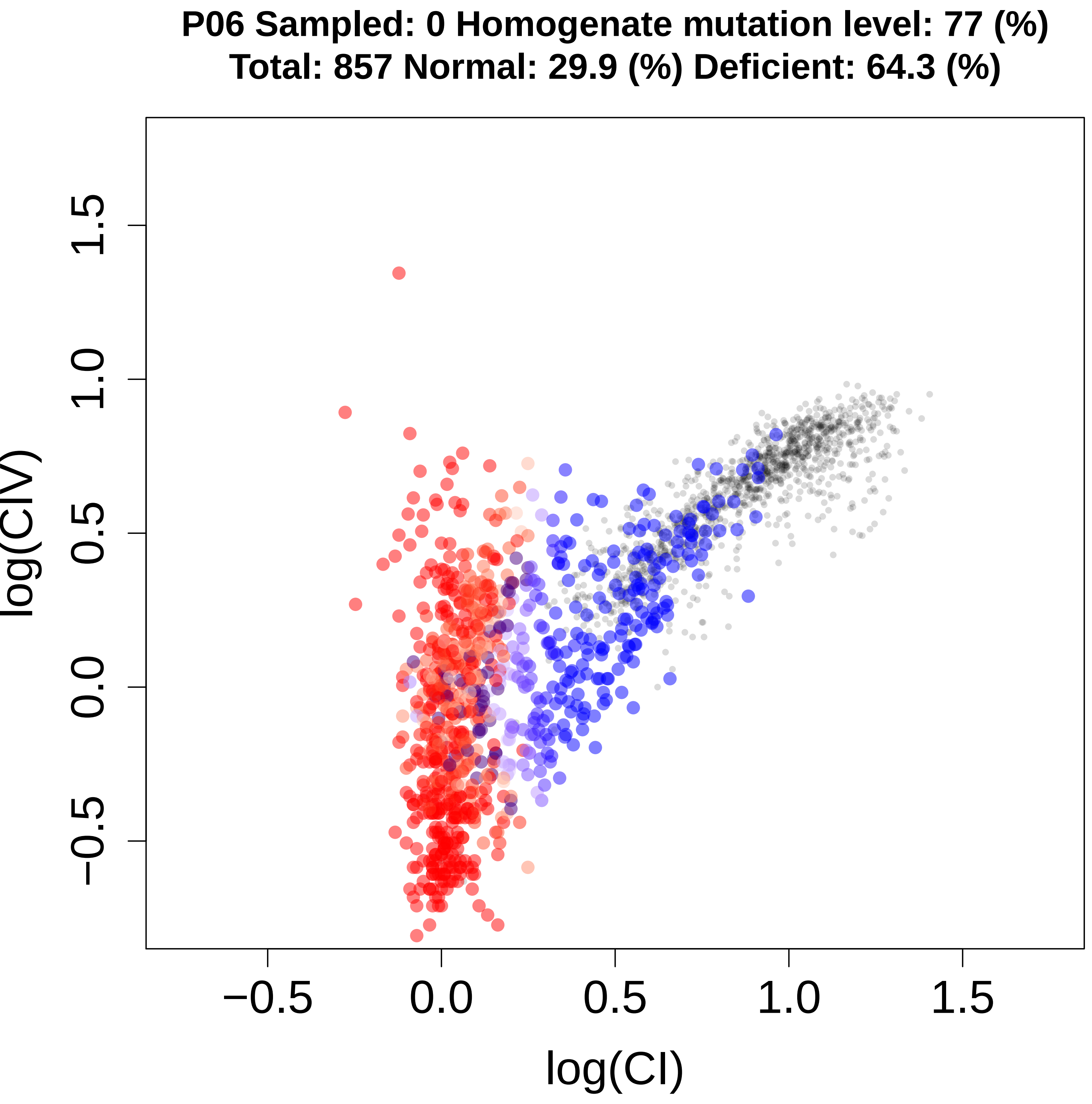
P06 Sampled: 0 Homogenate mutation level: 77 (%)
Total: 857 Normal: 29.9 (%) Deficient: 64.3 (%)
log(CIV)
log(CI)
−0.5
0.0
0.5
1.0
1.5

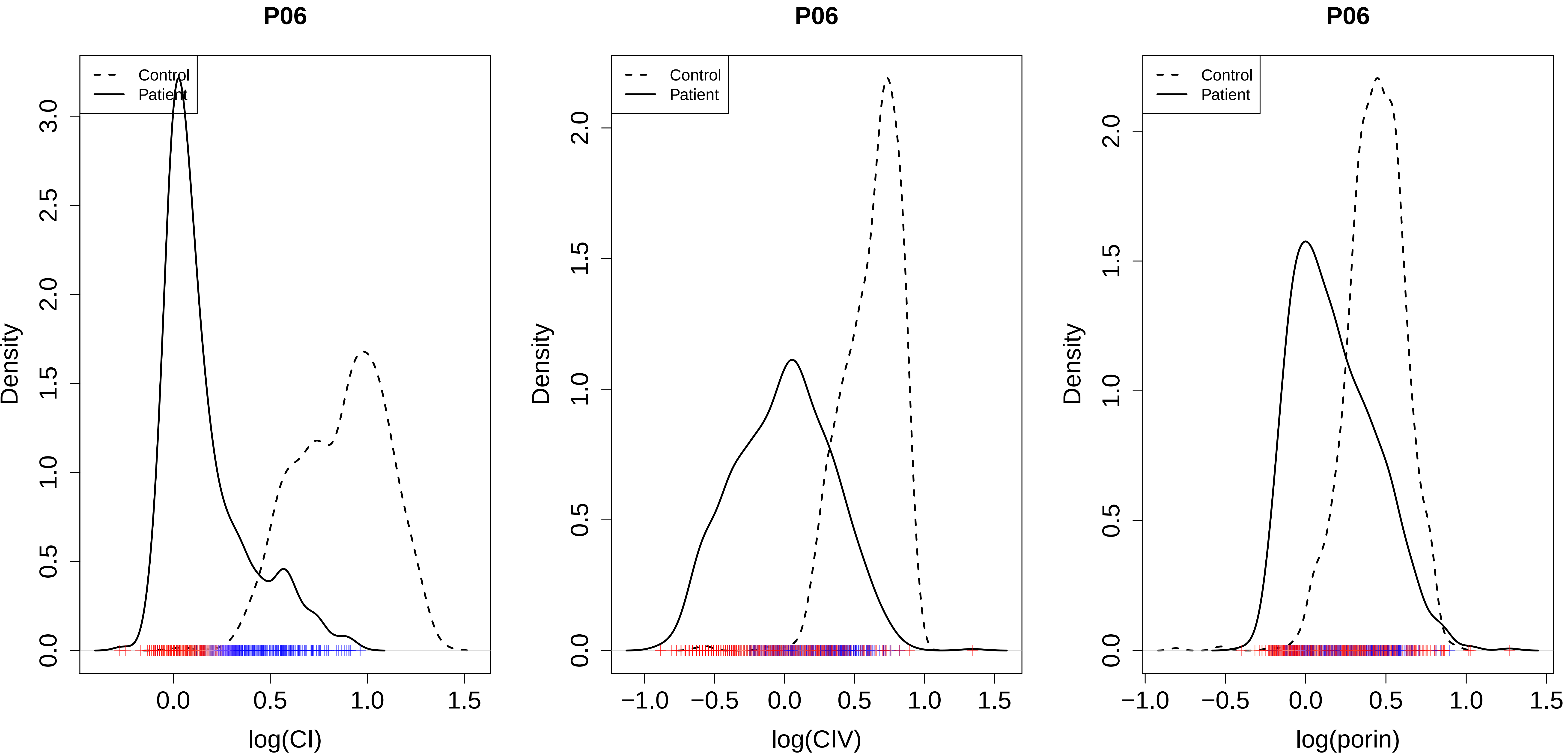

P06
Control
Patient
Density
log(CI)
P06
Control
Patient
Density
log(CIV)
P06
Control
Patient
Density
log(porin)

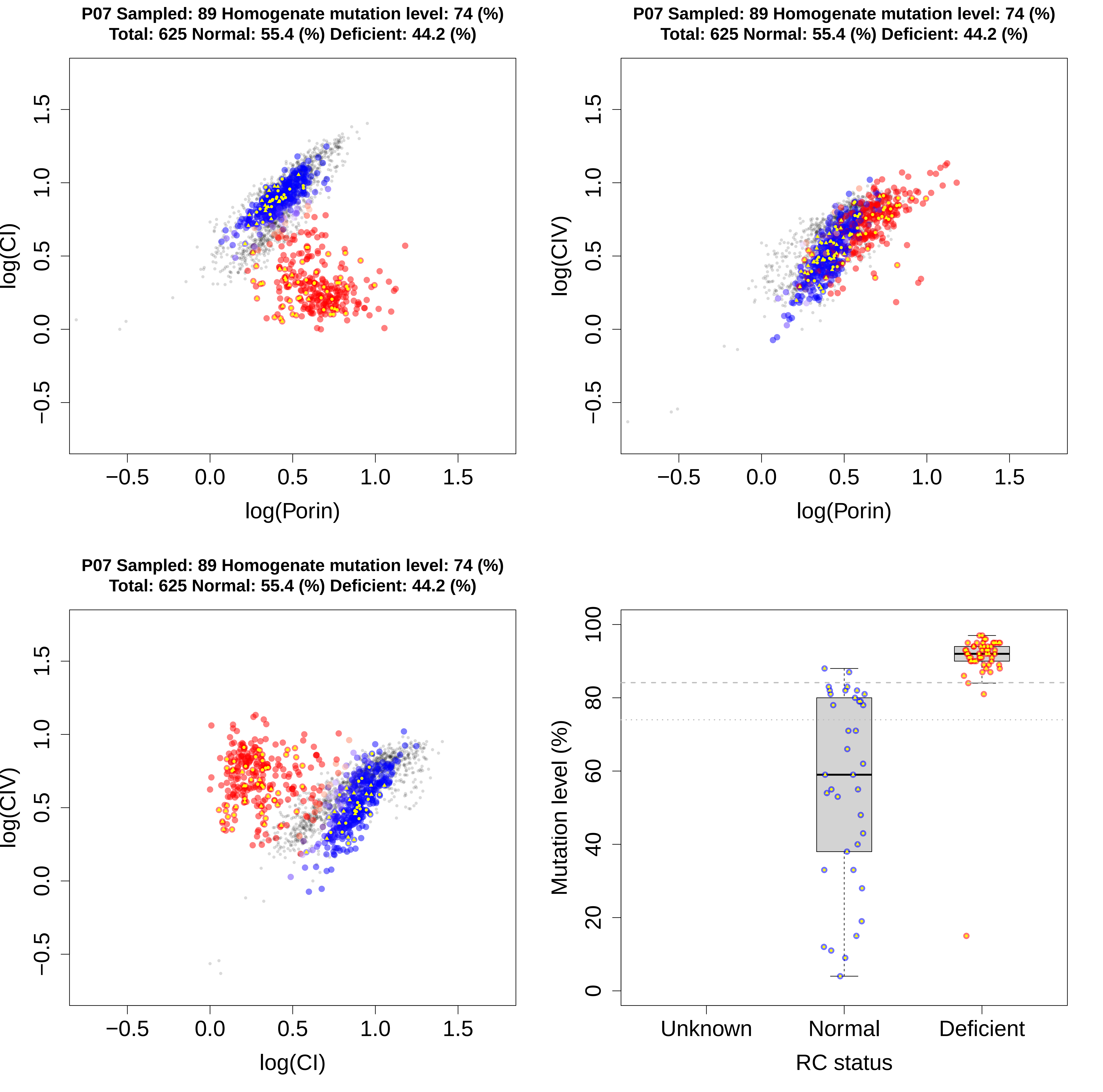

P07 Sampled: 89 Homogenate mutation level: 74 (%)
Total: 625 Normal: 55.4 (%) Deficient: 44.2 (%)
log(CI)
log(Porin)
P07 Sampled: 89 Homogenate mutation level: 74 (%)
Total: 625 Normal: 55.4 (%) Deficient: 44.2 (%)
log(CIV)
log(Porin)
P07 Sampled: 89 Homogenate mutation level: 74 (%)
Total: 625 Normal: 55.4 (%) Deficient: 44.2 (%)
log(CIV)
log(CI)
−0.5
0.0
0.5
1.0
1.5
Mutation level (%)
0
20
40
60
80
100
Unknown
Normal
Deficient
RC status

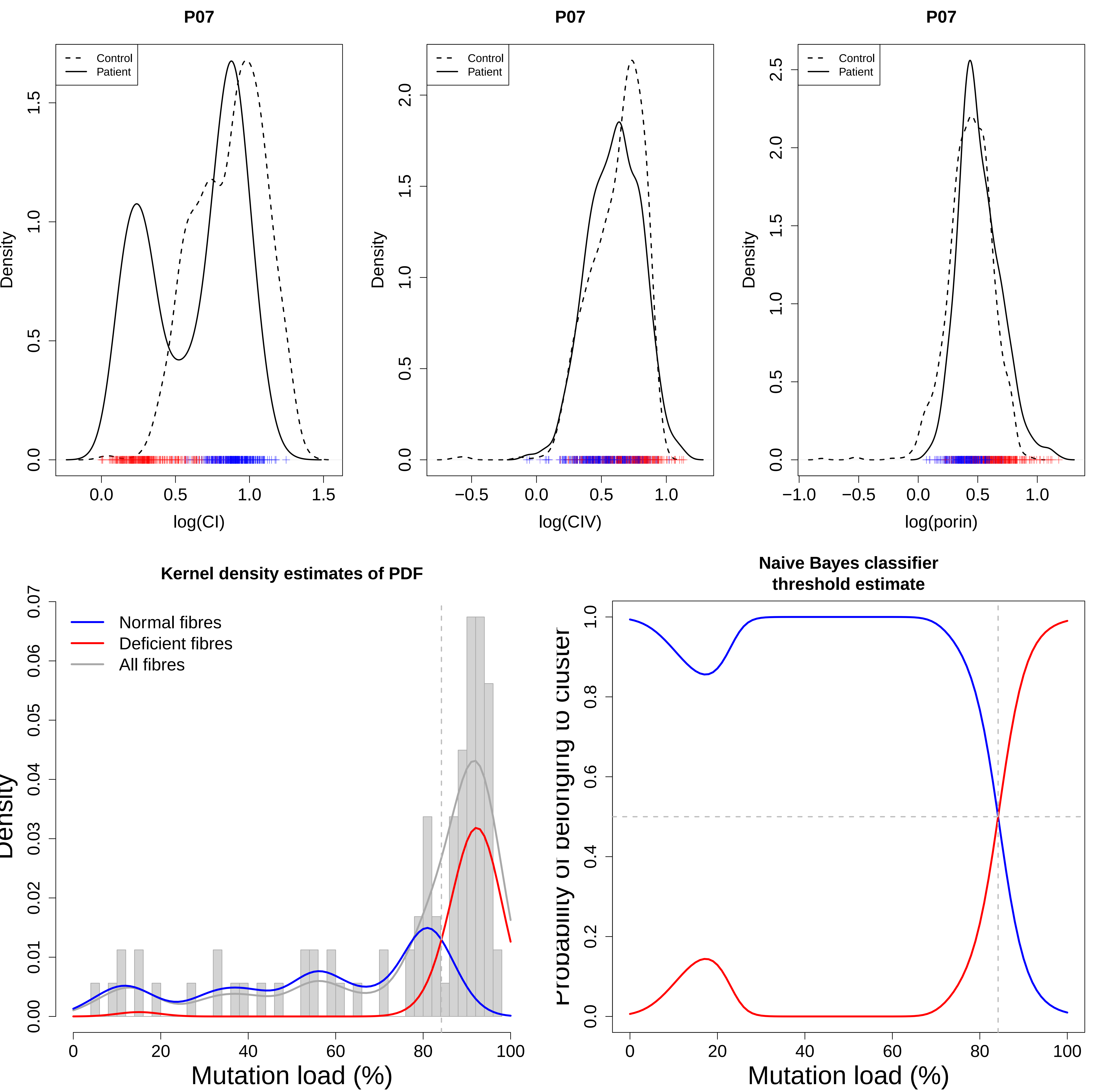
P07
Control
Patient
Density
log(CI)
P07
Control
Patient
Density
log(CIV)
P07
Control
Patient
Density
log(porin)
Kernel density estimates of PDF
Normal fibres
Deficient fibres
All fibres
Mutation load (%)
Naive Bayes classifier
threshold estimate
Mutation load (%)

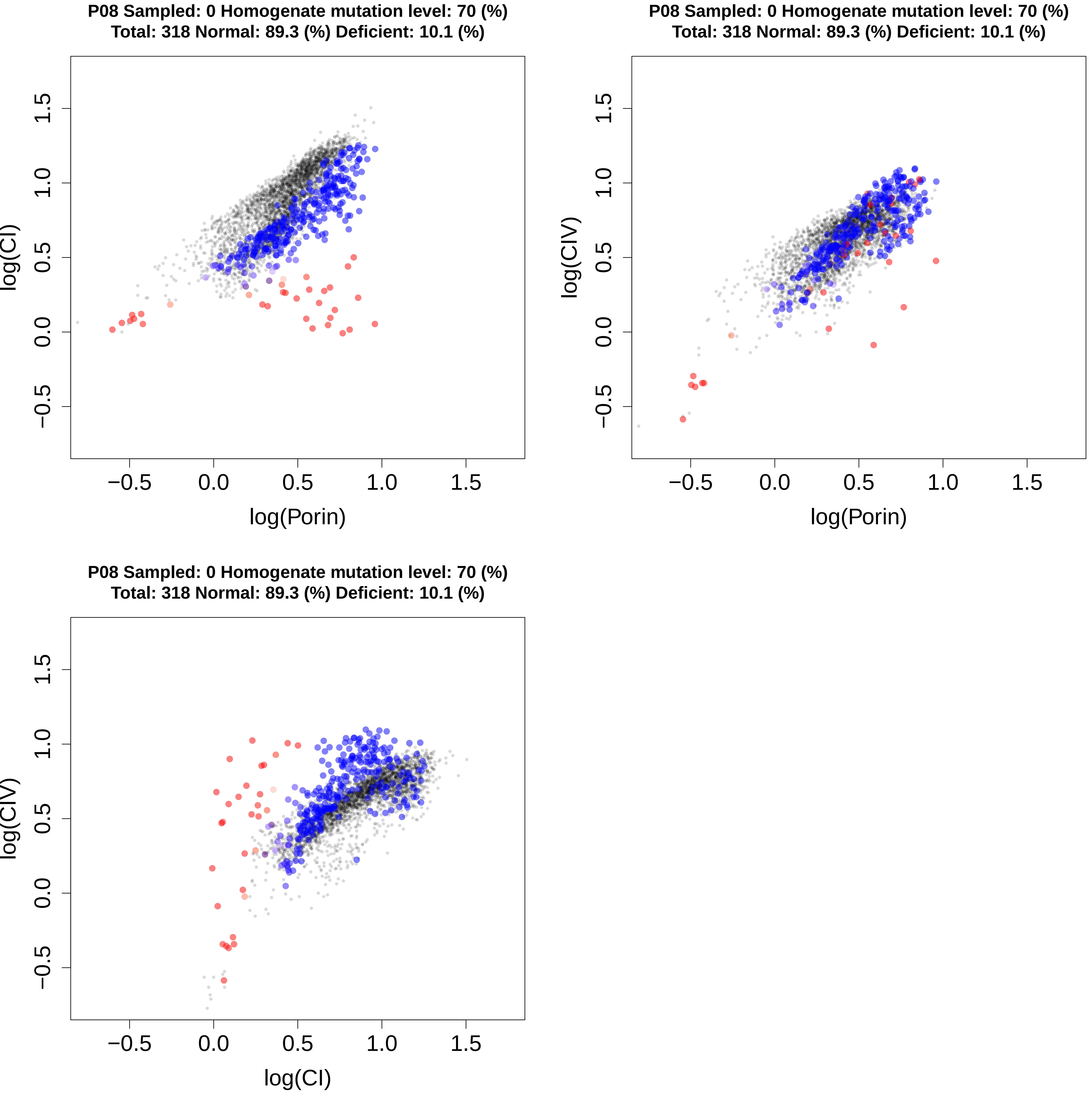
P08 Sampled: 0 Homogenate mutation level: 70 (%)
Total: 318 Normal: 89.3 (%) Deficient: 10.1 (%)
log(CI)
log(Porin)
P08 Sampled: 0 Homogenate mutation level: 70 (%)
Total: 318 Normal: 89.3 (%) Deficient: 10.1 (%)
log(CIV)
log(Porin)
P08 Sampled: 0 Homogenate mutation level: 70 (%)
Total: 318 Normal: 89.3 (%) Deficient: 10.1 (%)
log(CIV)
log(CI)

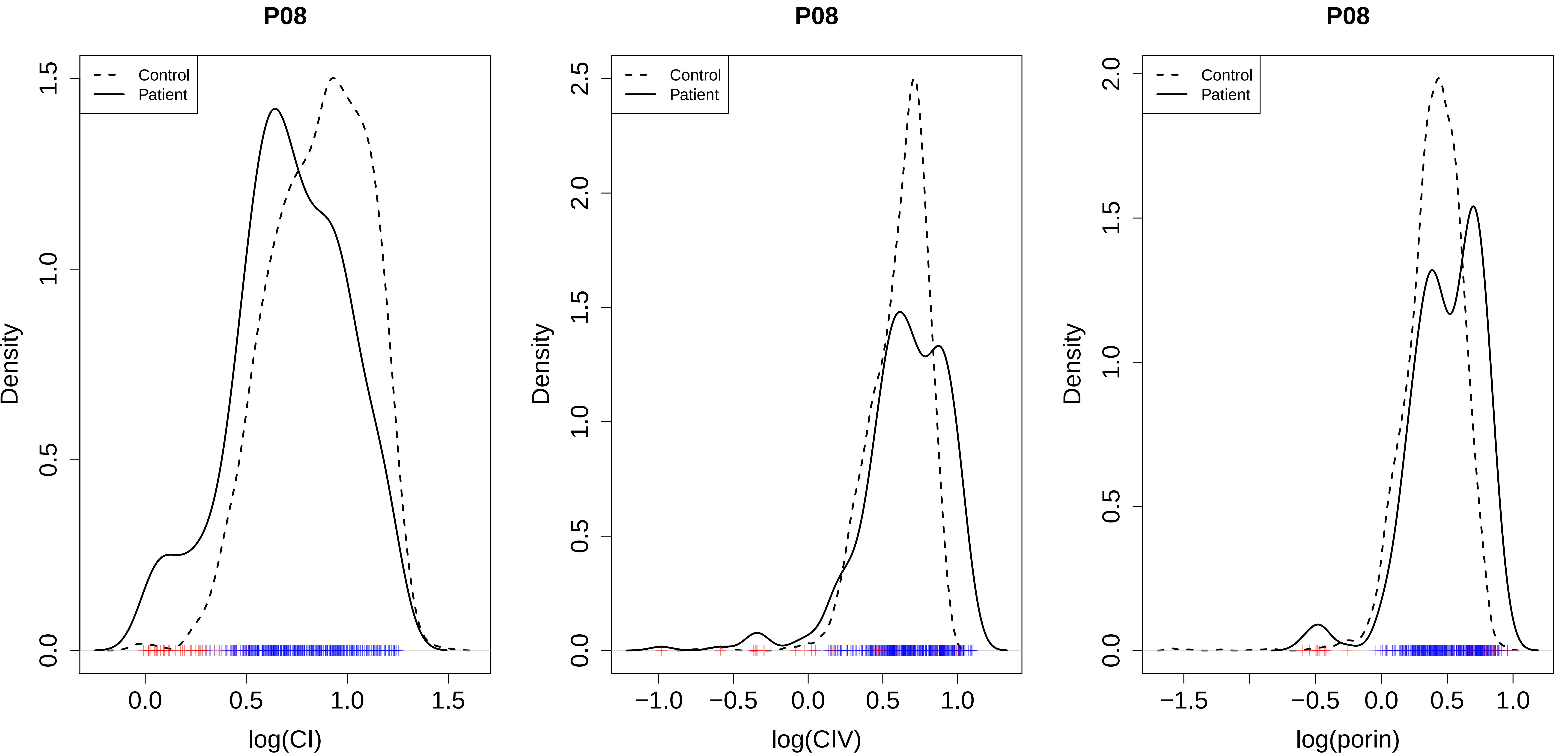
P08
Control
Patient
Density
log(CI)
P08
Control
Patient
Density
log(CIV)
P08
Control
Patient
Density
log(porin)

P09 Sampled: 0 Homogenate mutation level: 67 (%)
Total: 104 Normal: 84.6 (%) Deficient: 15.4 (%)

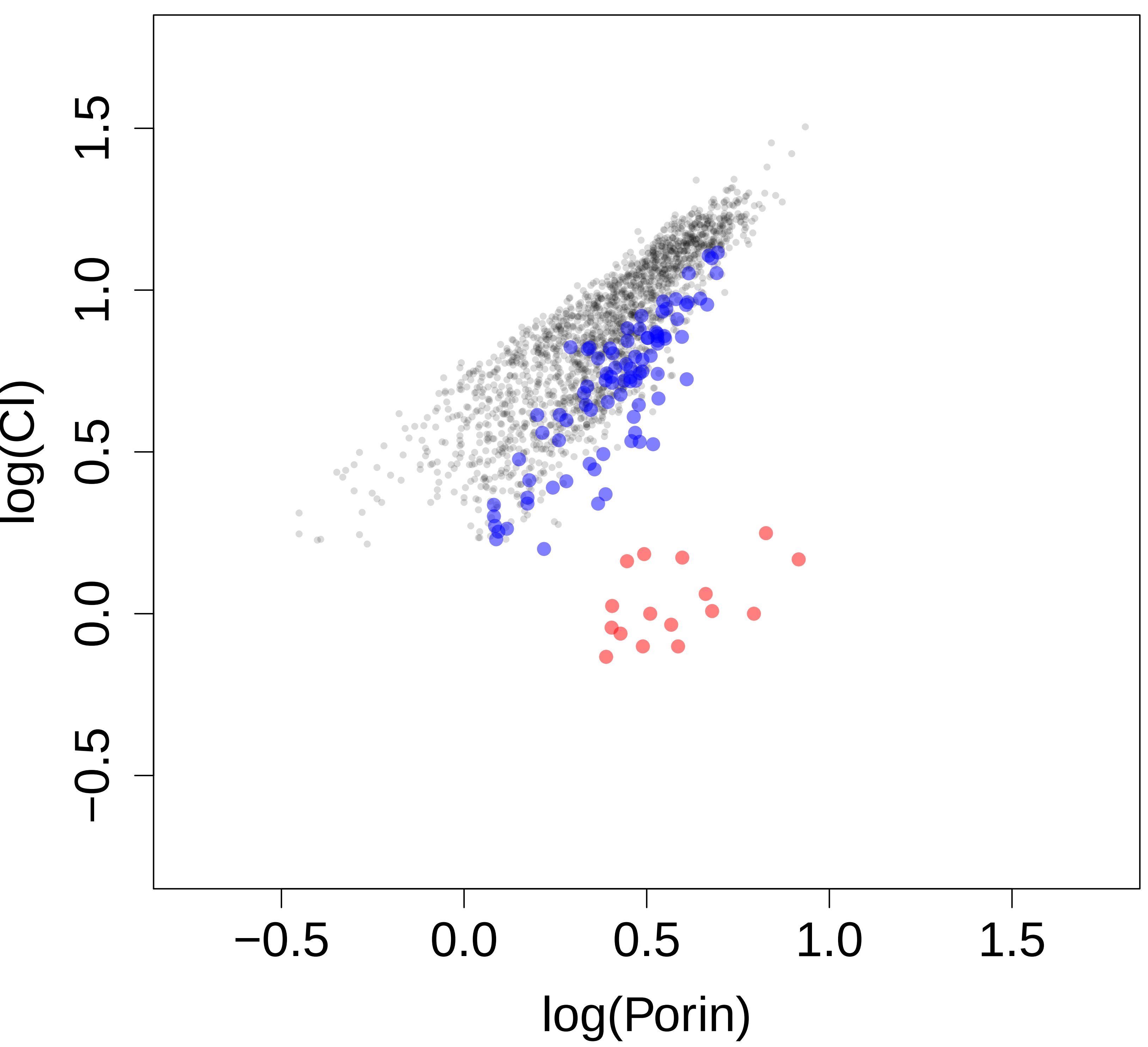


P09 Sampled: 0 Homogenate mutation level: 67 (%)
Total: 104 Normal: 84.6 (%) Deficient: 15.4 (%)

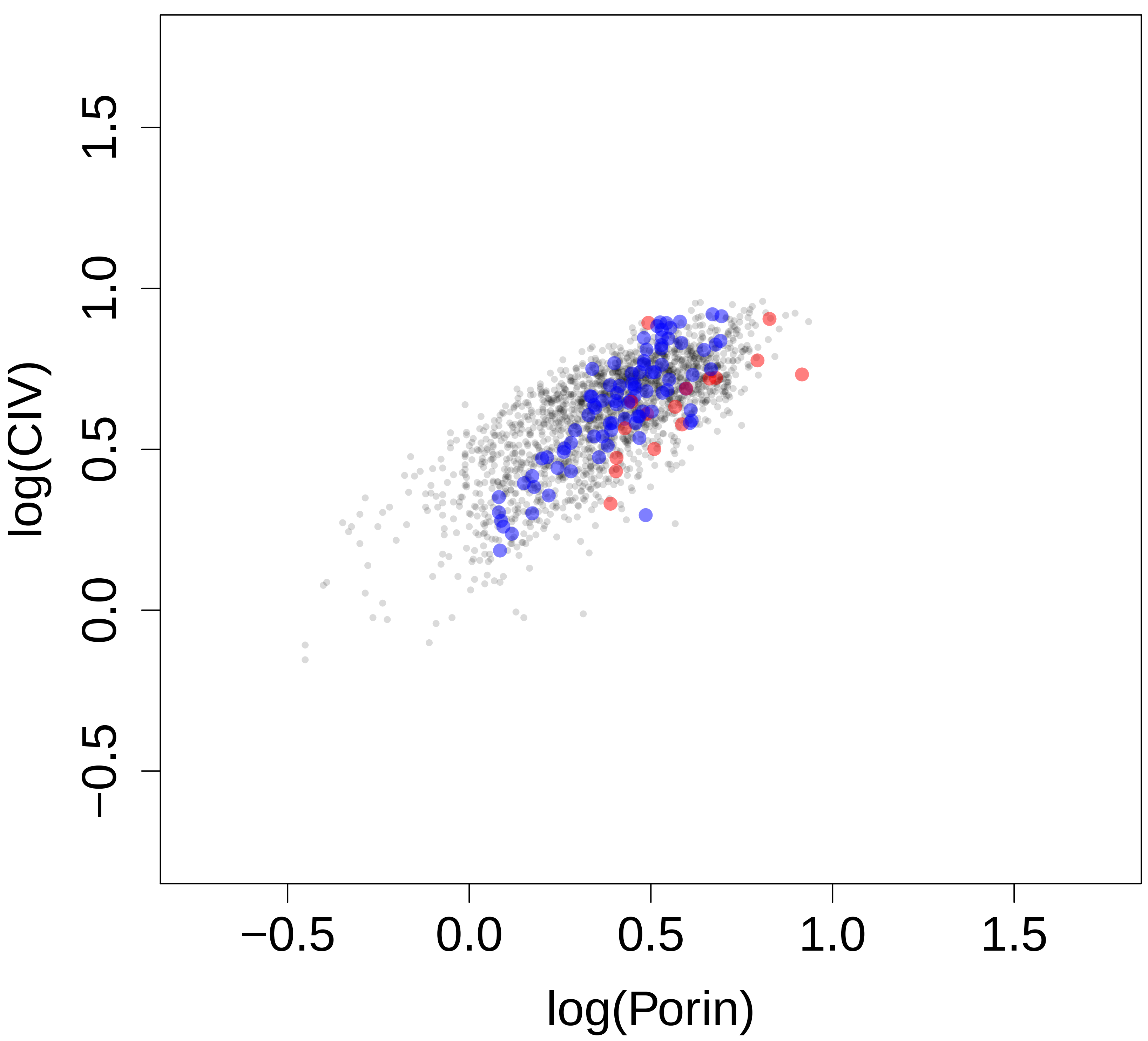


P09 Sampled: 0 Homogenate mutation level: 67 (%)
Total: 104 Normal: 84.6 (%) Deficient: 15.4 (%)

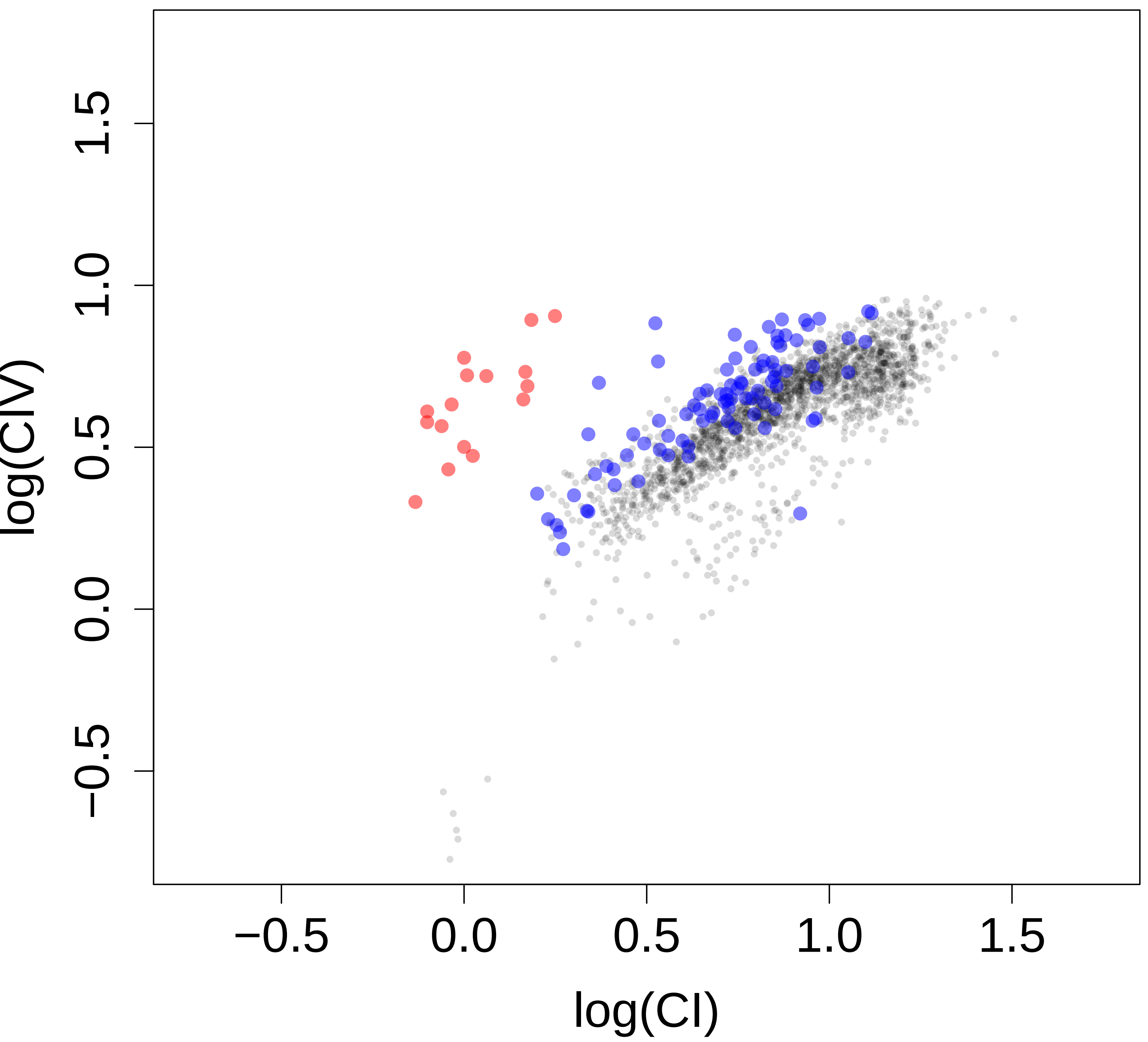

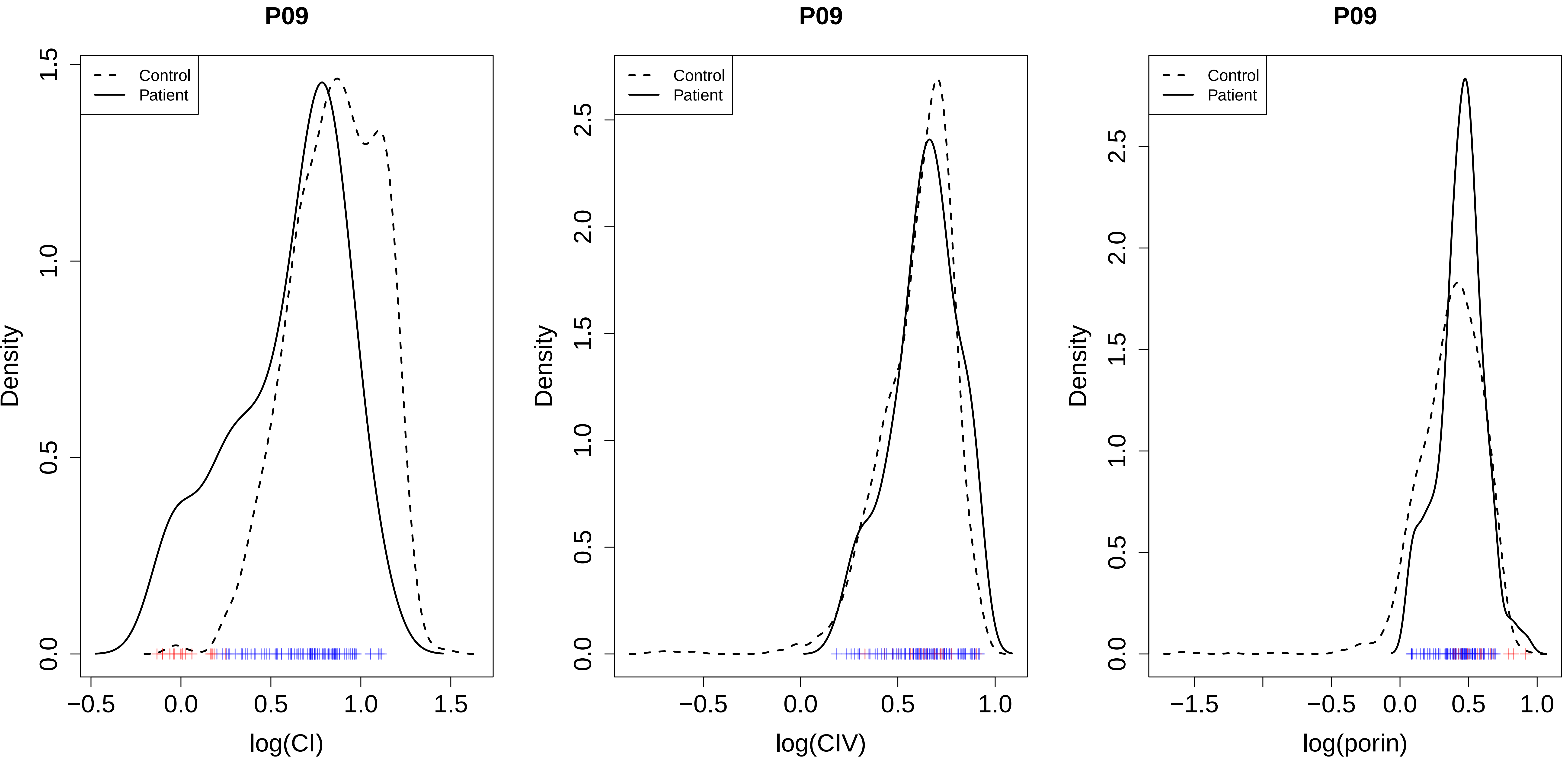

P09
Control
Patient
Density
log(CI)
P09
Control
Patient
Density
log(CIV)
P09
Control
Patient
Density
log(porin)

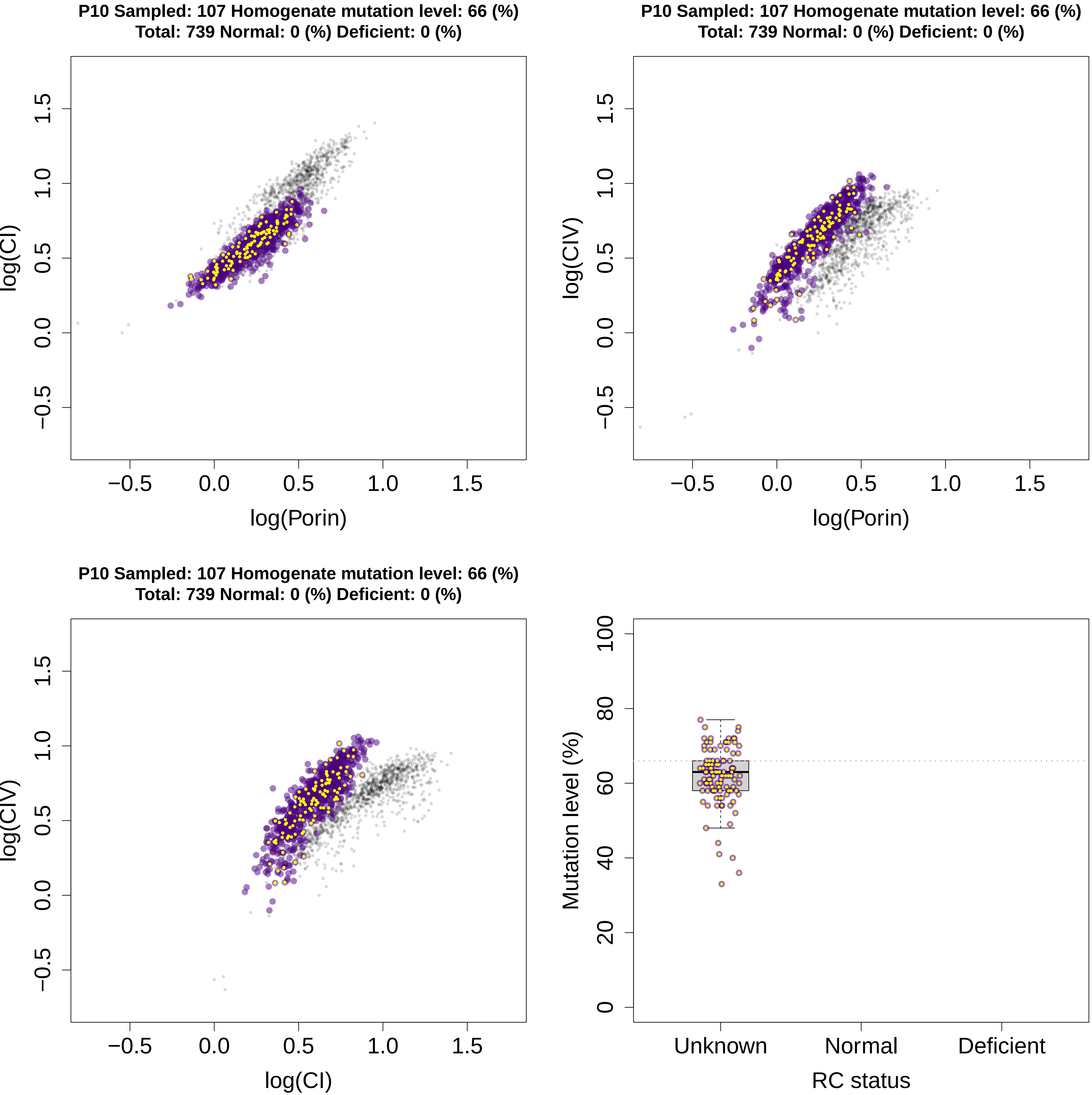
P10 Sampled: 107 Homogenate mutation level: 66 (%)
Total: 739 Normal: 0 (%) Deficient: 0 (%)
log(CI)
log(Porin)
−0.5 0.0 0.5 1.0 1.5
P10 Sampled: 107 Homogenate mutation level: 66 (%)
Total: 739 Normal: 0 (%) Deficient: 0 (%)
log(CIV)
log(Porin)
−0.5 0.0 0.5 1.0 1.5
P10 Sampled: 107 Homogenate mutation level: 66 (%)
Total: 739 Normal: 0 (%) Deficient: 0 (%)
log(CIV)
log(CI)
−0.5 0.0 0.5 1.0 1.5
Mutation level (%)
0 20 40 60 80 100
Unknown
Normal
Deficient
RC status

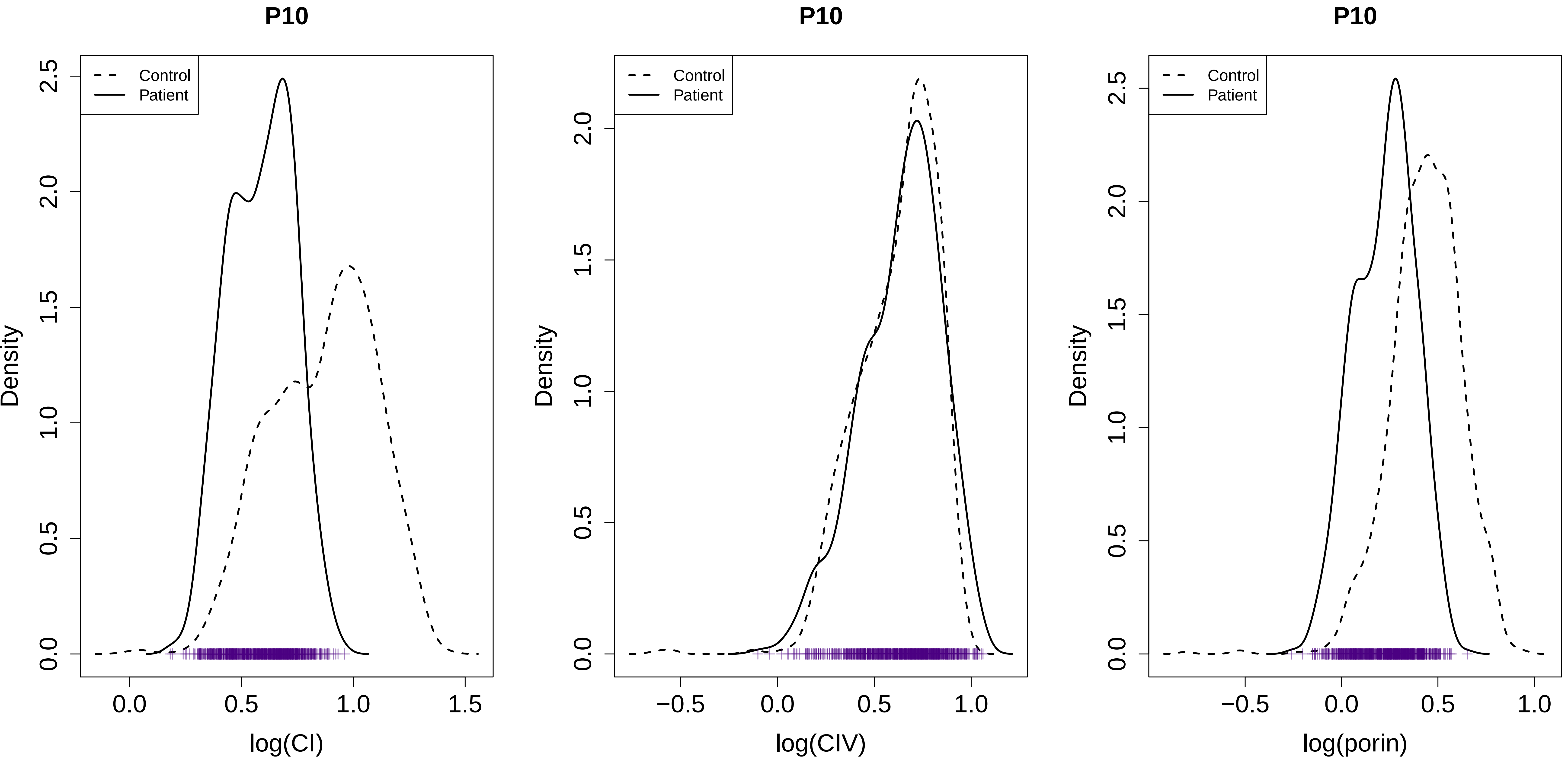
P10
Control
Patient
Density
log(CI)
P10
Control
Patient
Density
log(CIV)
P10
Control
Patient
Density
log(porin)

**P11 Sampled: 0 Homogenate mutation level: 63 (%)**
**Total: 431 Normal: 94.4 (%) Deficient: 5.34 (%)**

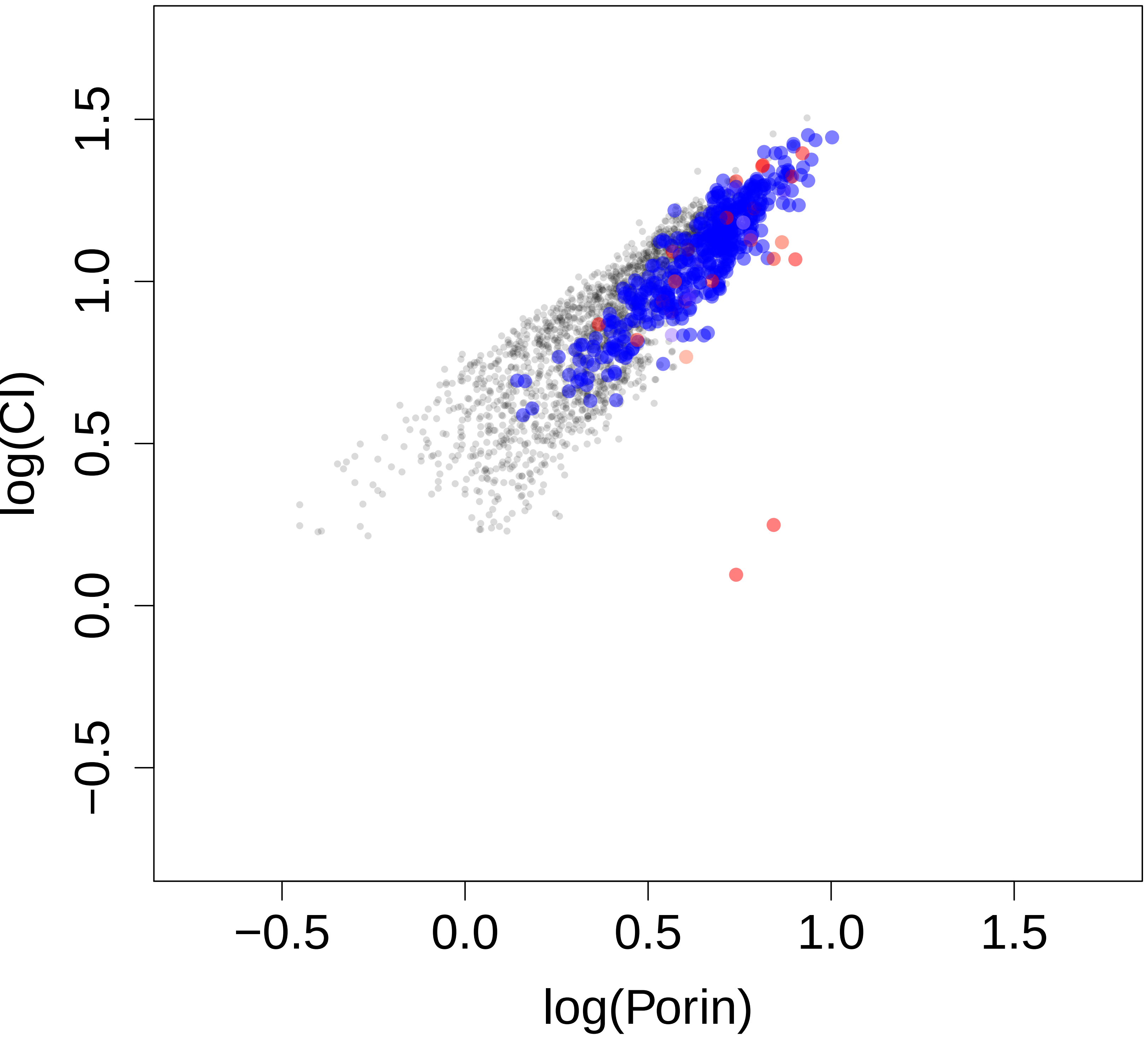


**P11 Sampled: 0 Homogenate mutation level: 63 (%)**
**Total: 431 Normal: 94.4 (%) Deficient: 5.34 (%)**

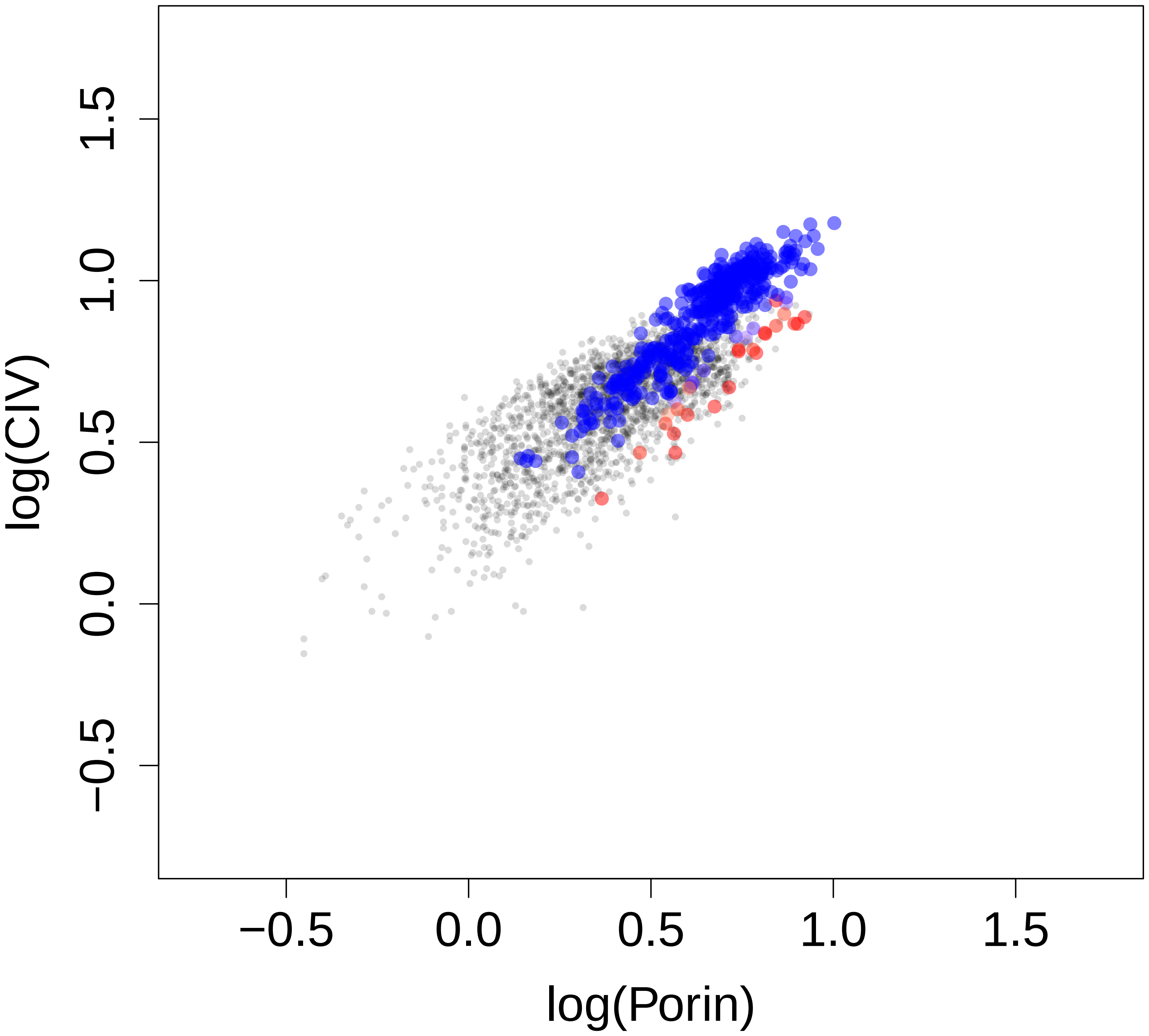


**P11 Sampled: 0 Homogenate mutation level: 63 (%)**
**Total: 431 Normal: 94.4 (%) Deficient: 5.34 (%)**

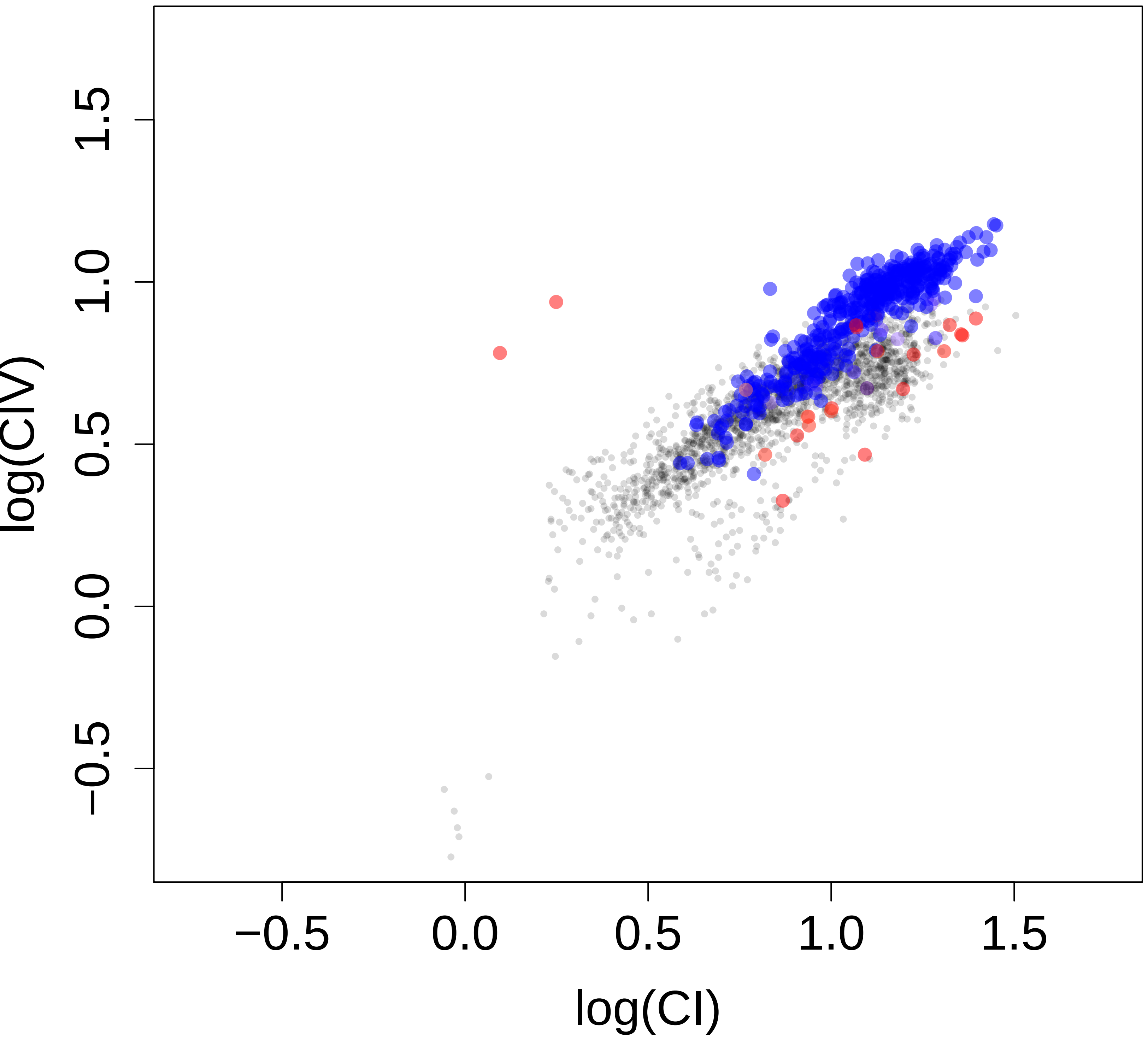

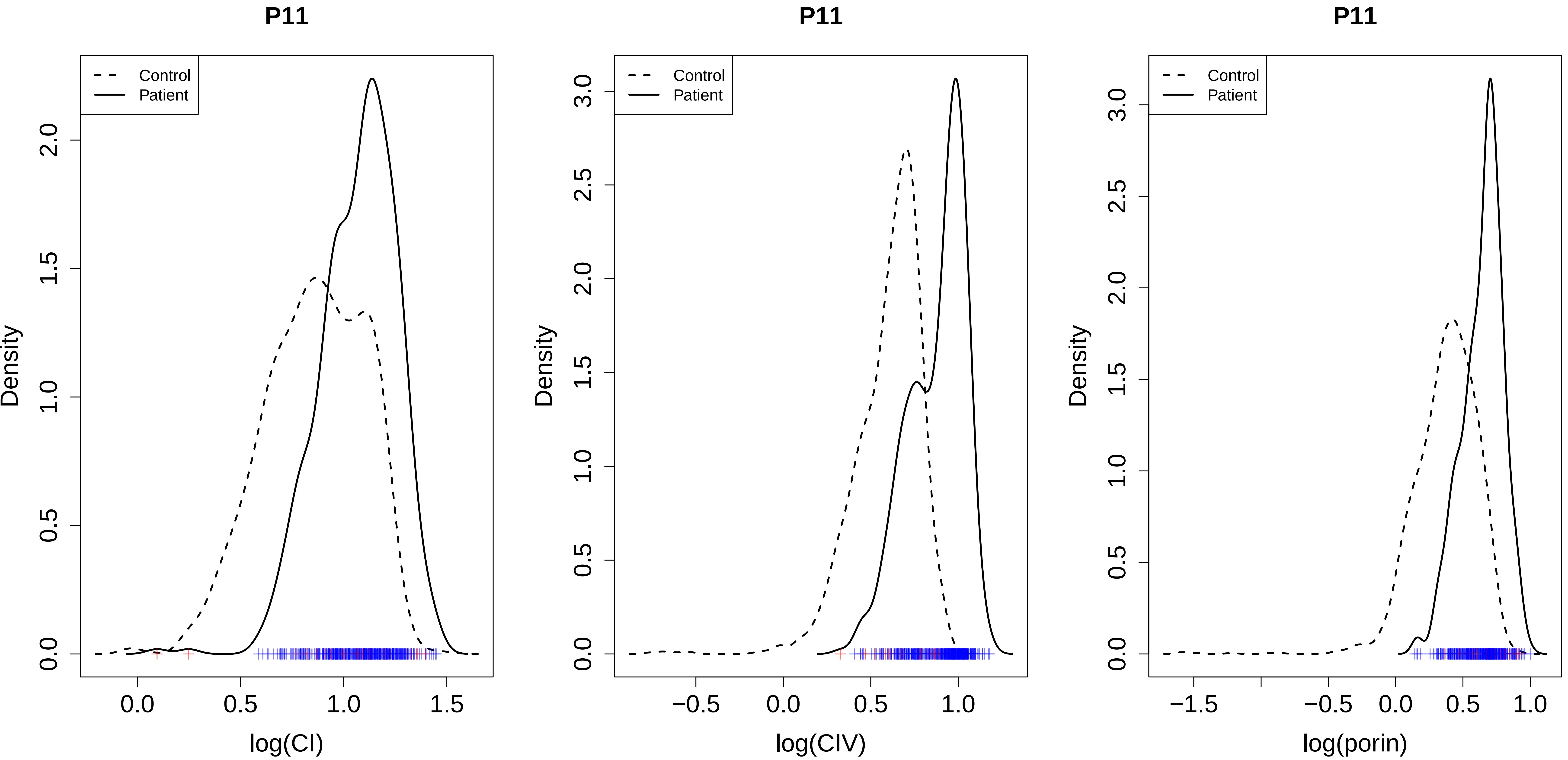
P11
Control
Patient
Density
log(CI)
P11
Control
Patient
Density
log(CIV)
P11
Control
Patient
Density
log(porin)

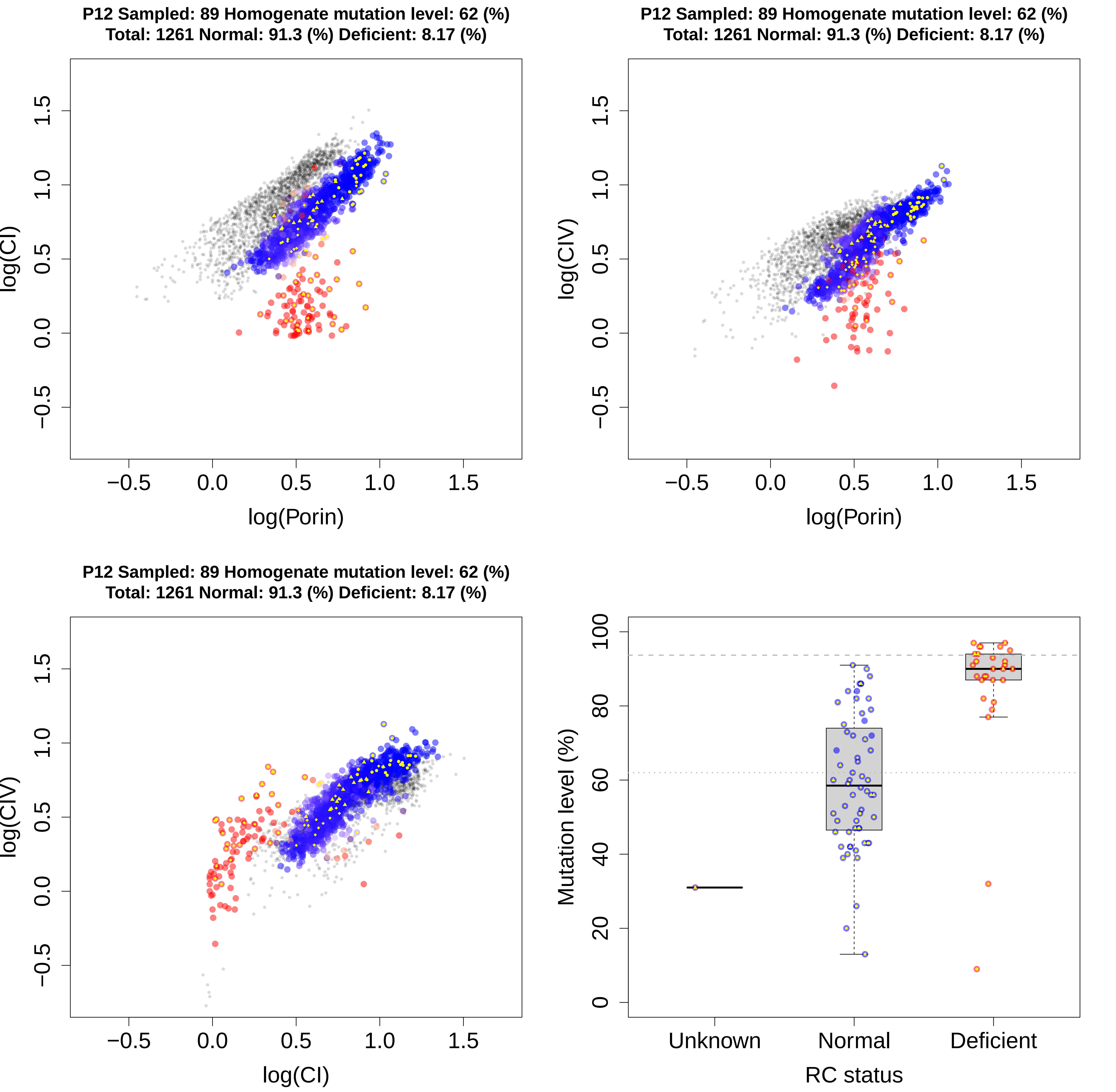
P12 Sampled: 89 Homogenate mutation level: 62 (%)
Total: 1261 Normal: 91.3 (%) Deficient: 8.17 (%)
log(CI)
log(Porin)
P12 Sampled: 89 Homogenate mutation level: 62 (%)
Total: 1261 Normal: 91.3 (%) Deficient: 8.17 (%)
log(CIV)
log(Porin)
P12 Sampled: 89 Homogenate mutation level: 62 (%)
Total: 1261 Normal: 91.3 (%) Deficient: 8.17 (%)
log(CIV)
log(CI)
Mutation level (%)
Unknown
Normal
Deficient
RC status

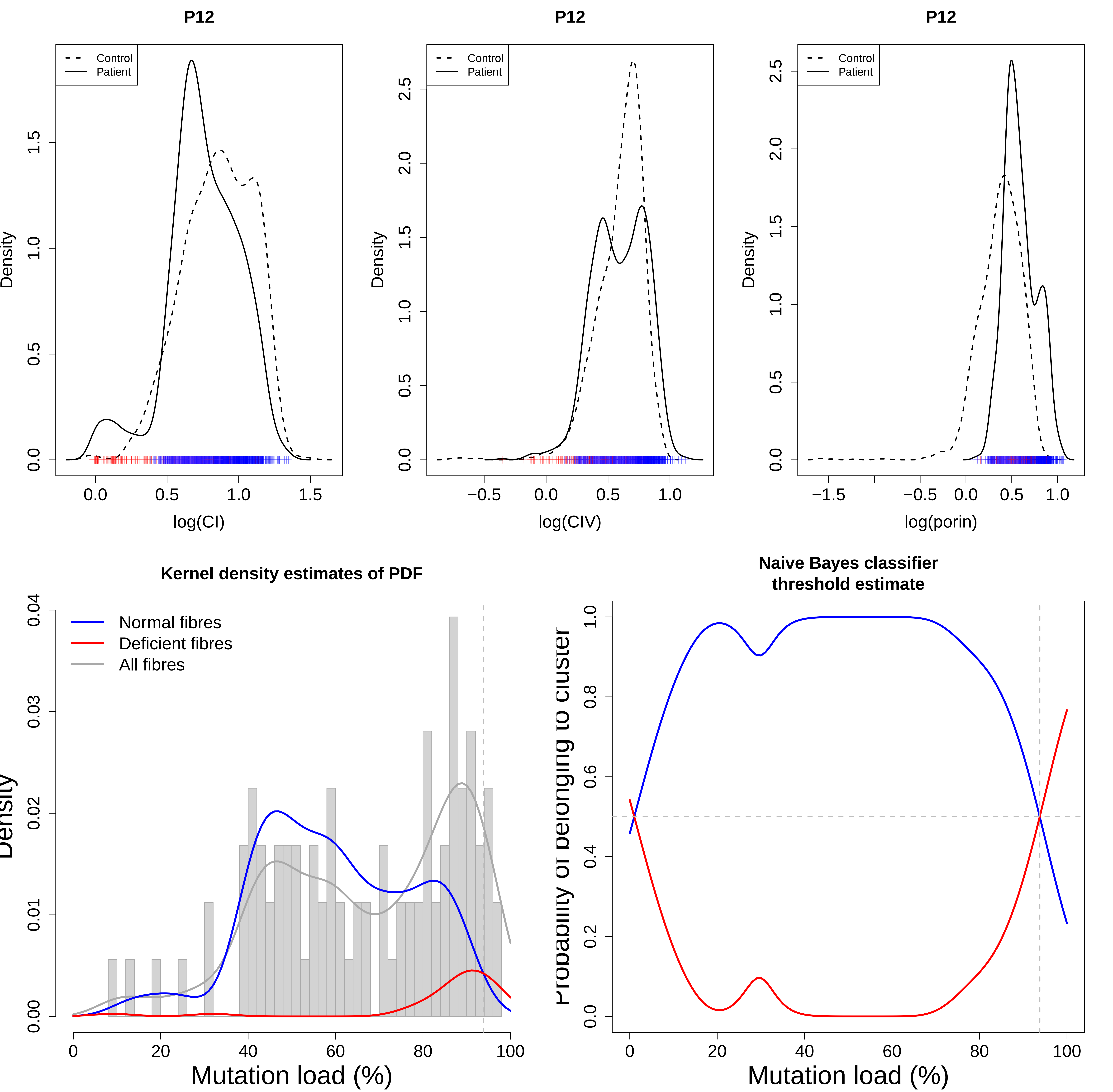
P12
Control
Patient
Density
log(CI)
P12
Control
Patient
Density
log(CIV)
P12
Control
Patient
Density
log(porin)
Kernel density estimates of PDF
Normal fibres
Deficient fibres
All fibres
Density
Mutation load (%)
Naive Bayes classifier
threshold estimate
Probability of belonging to cluster
Mutation load (%)

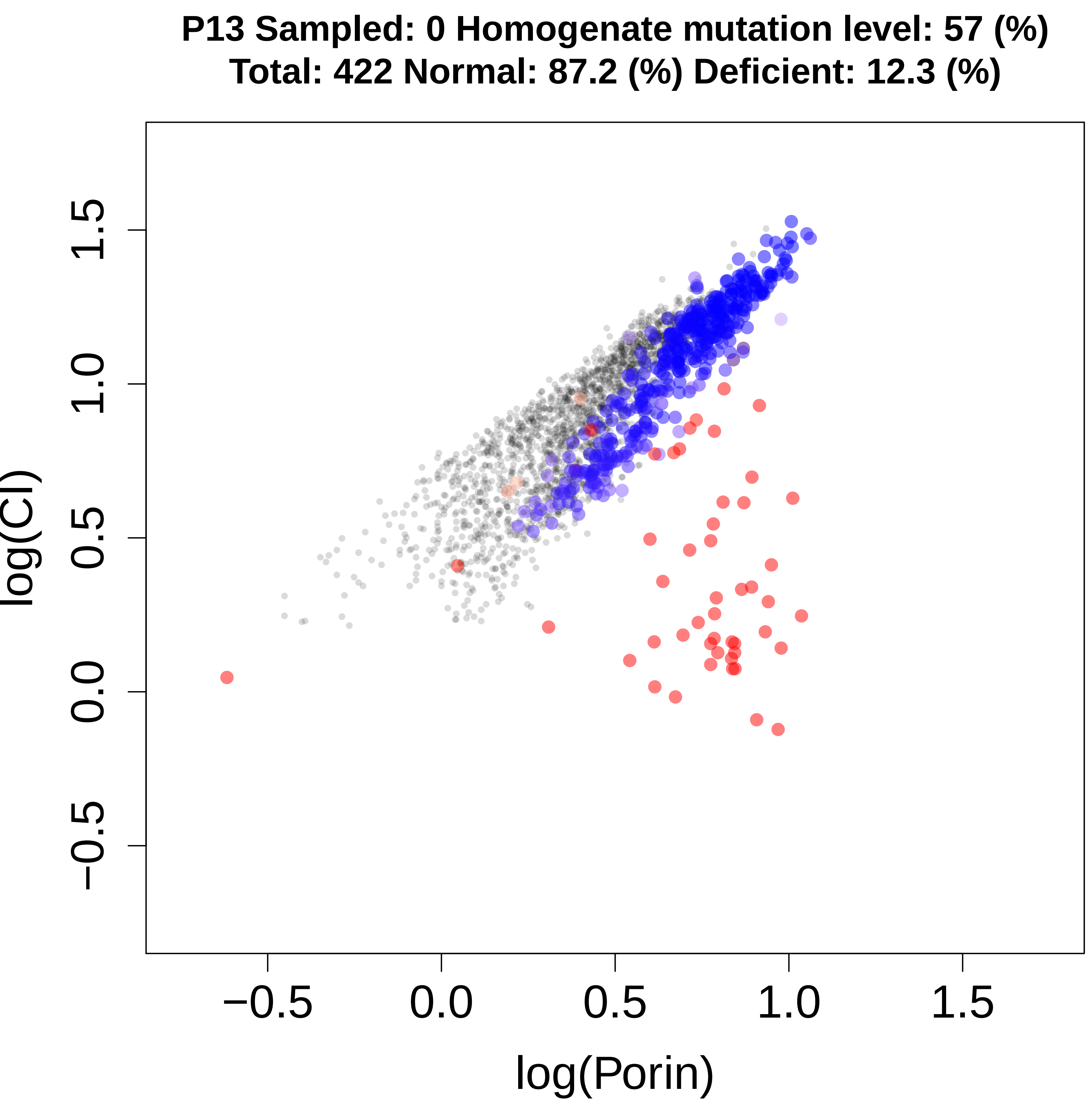
P13 Sampled: 0 Homogenate mutation level: 57 (%)
Total: 422 Normal: 87.2 (%) Deficient: 12.3 (%)
log(CI)
−0.5
0.0
0.5
1.0
1.5
log(Porin)

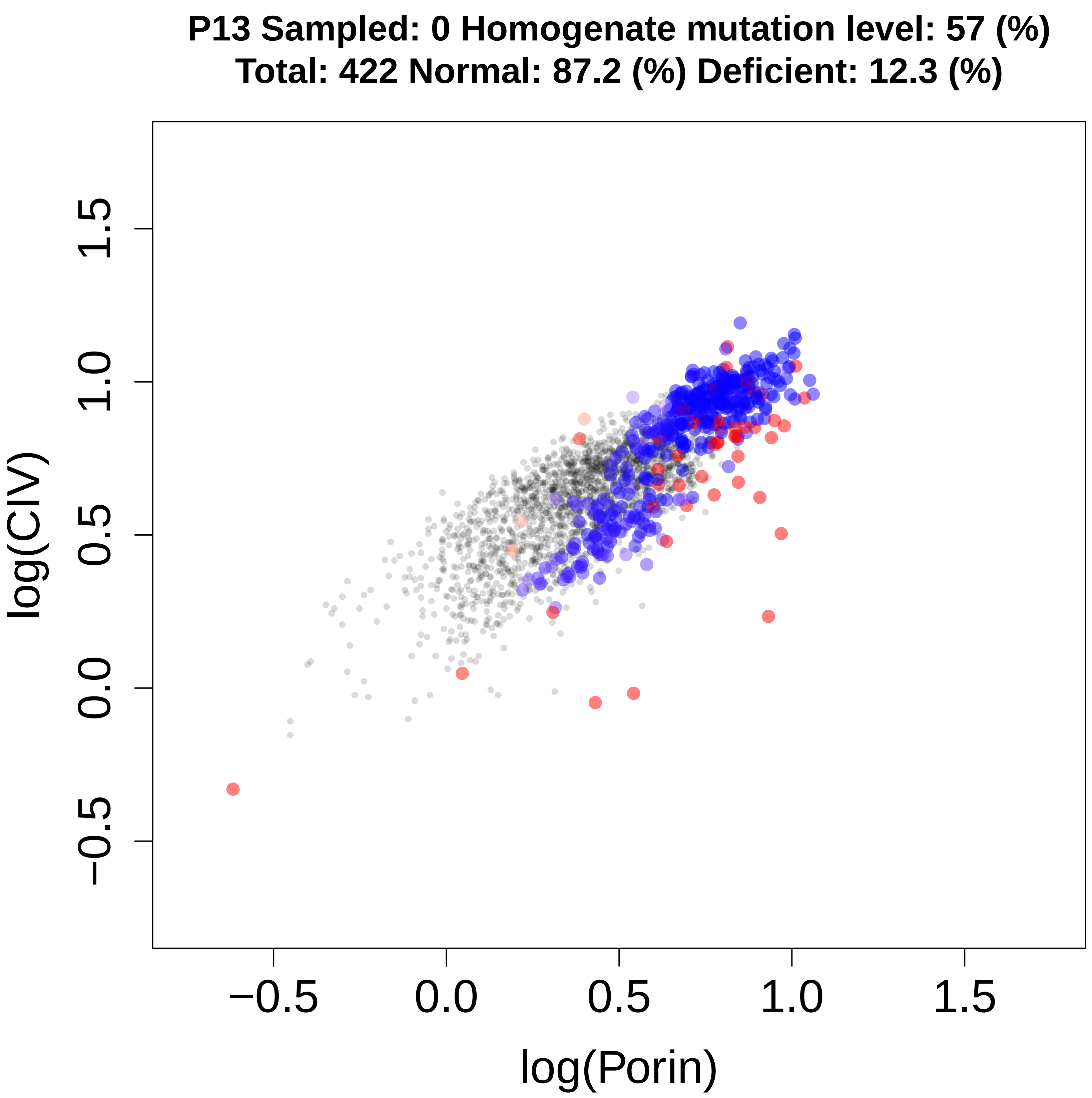
P13 Sampled: 0 Homogenate mutation level: 57 (%)
Total: 422 Normal: 87.2 (%) Deficient: 12.3 (%)
log(CIV)
−0.5
0.0
0.5
1.0
1.5
log(Porin)

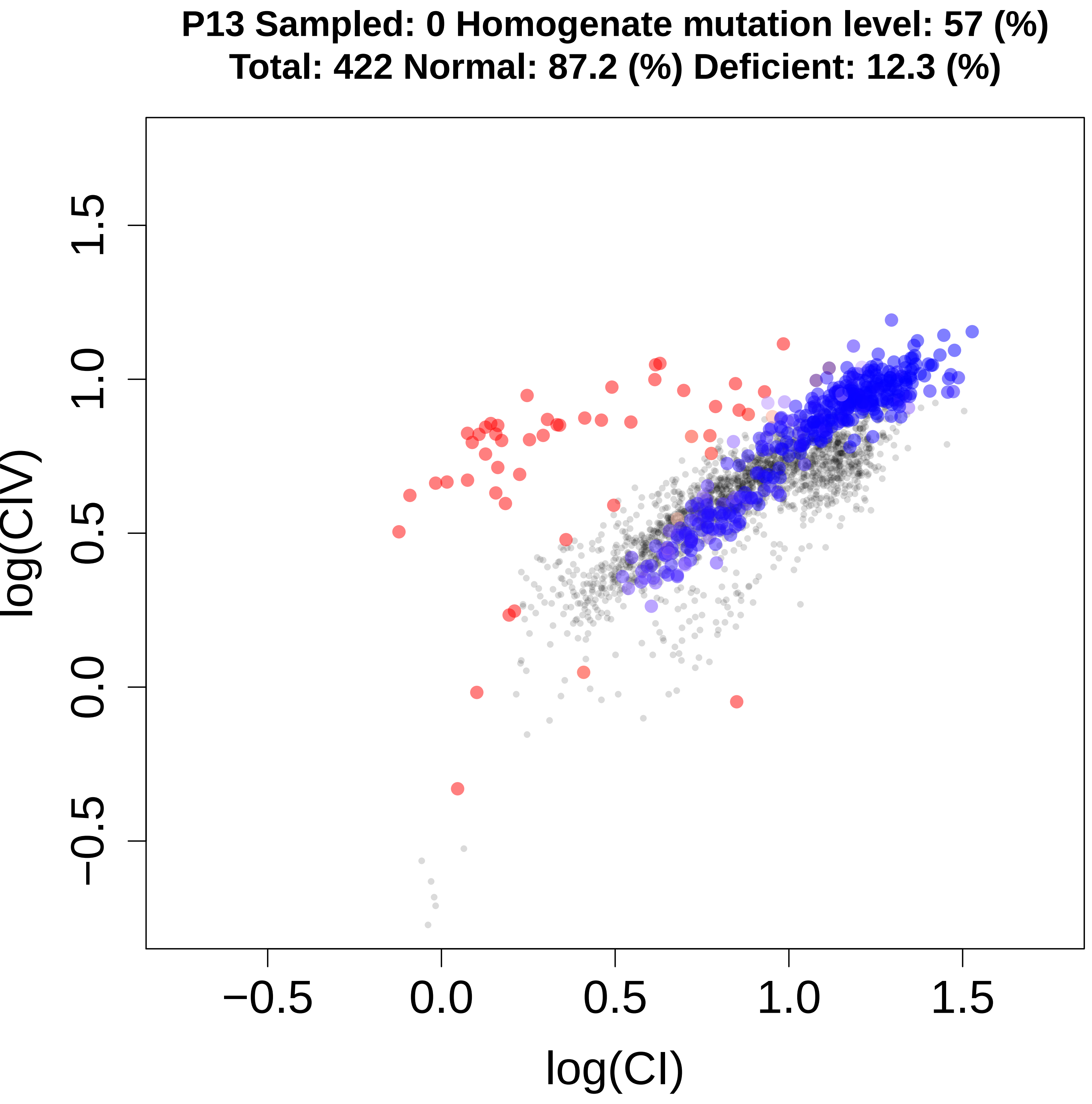
P13 Sampled: 0 Homogenate mutation level: 57 (%)
Total: 422 Normal: 87.2 (%) Deficient: 12.3 (%)
log(CIV)
−0.5
0.0
0.5
1.0
1.5
log(CI)

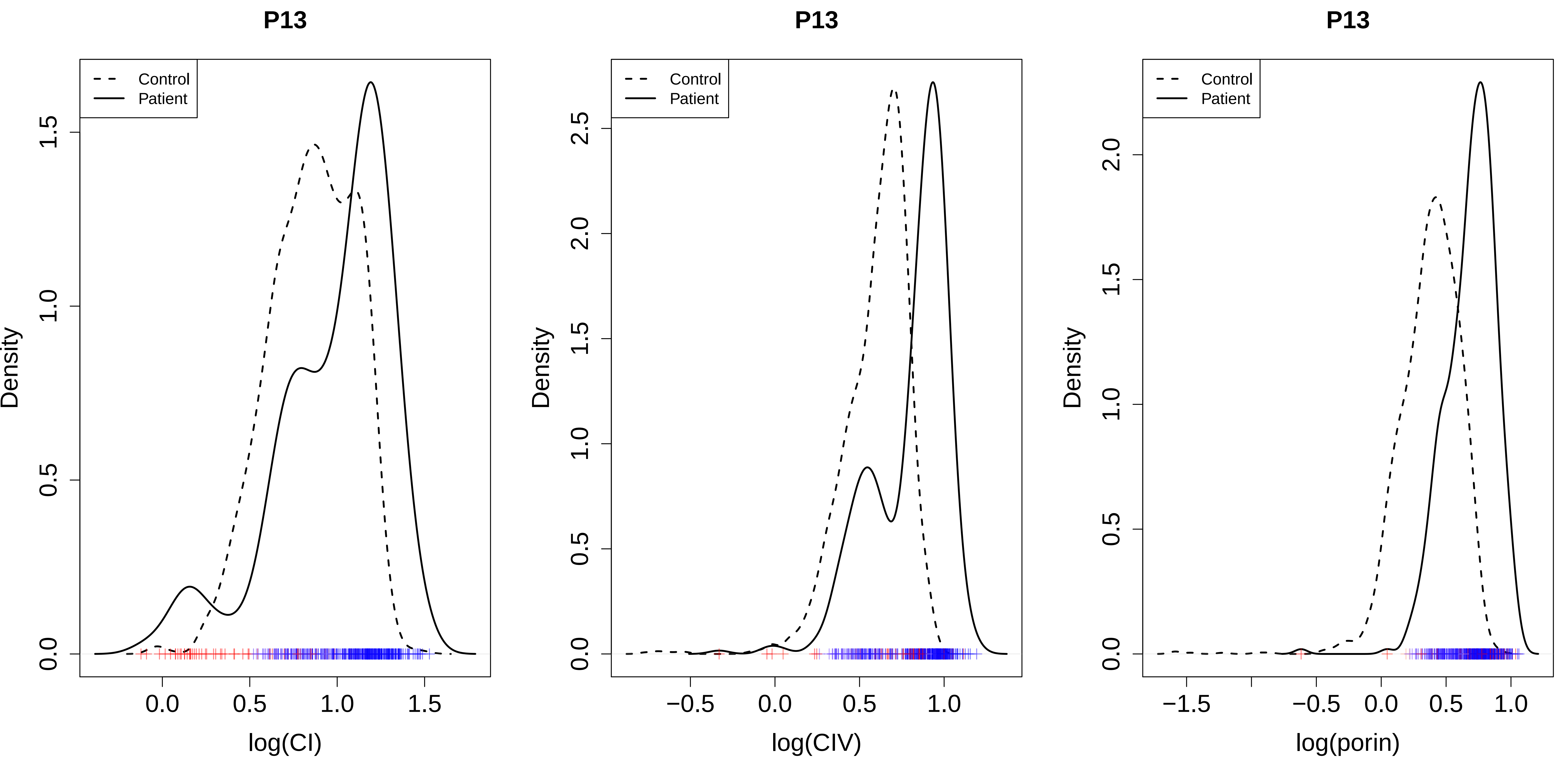

P13
Control
Patient
Density
log(CI)
P13
Control
Patient
Density
log(CIV)
P13
Control
Patient
Density
log(porin)

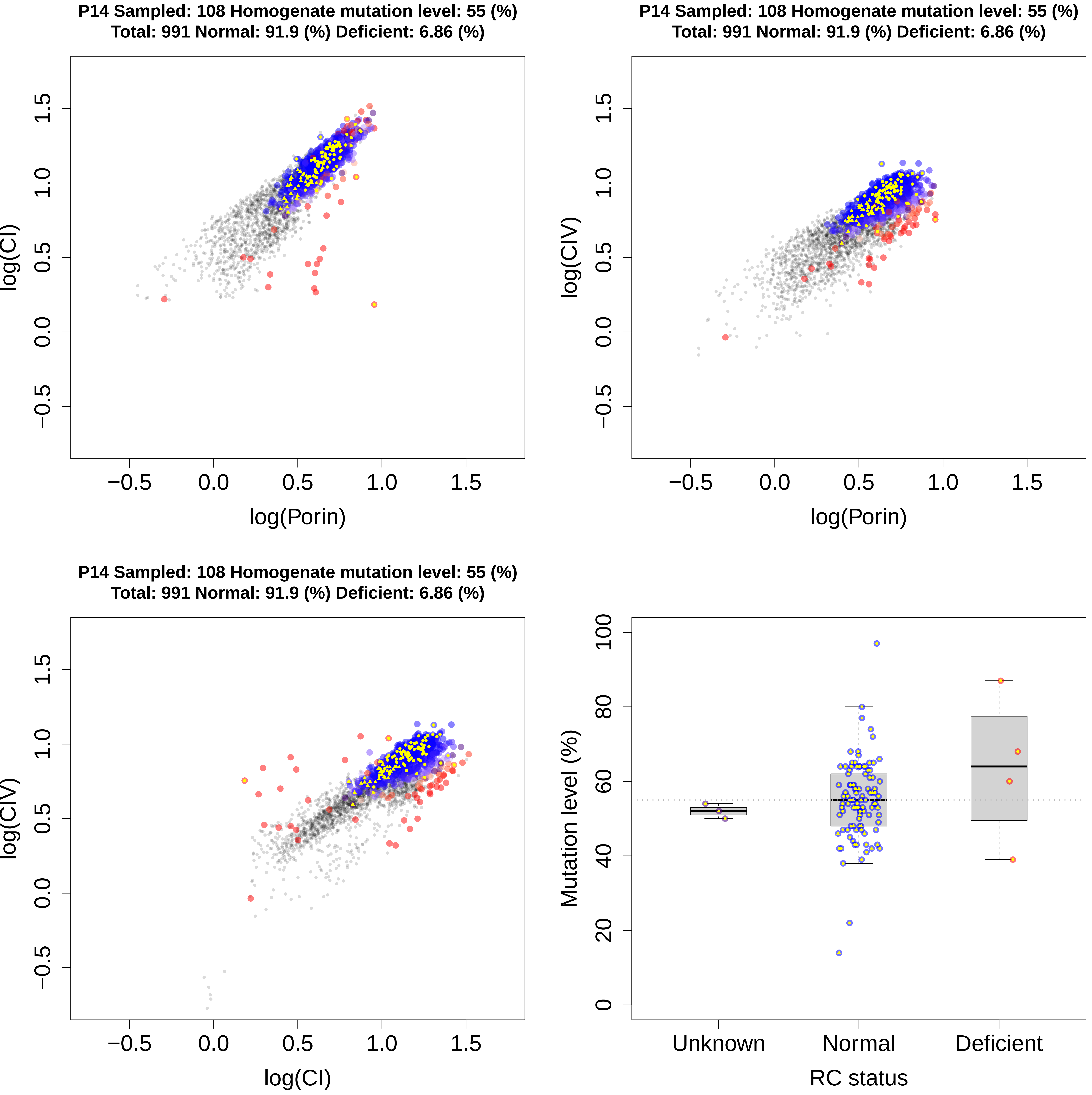
P14 Sampled: 108 Homogenate mutation level: 55 (%)
Total: 991 Normal: 91.9 (%) Deficient: 6.86 (%)
log(CI)
log(Porin)
−0.5
0.0
0.5
1.0
1.5
P14 Sampled: 108 Homogenate mutation level: 55 (%)
Total: 991 Normal: 91.9 (%) Deficient: 6.86 (%)
log(CIV)
log(Porin)
P14 Sampled: 108 Homogenate mutation level: 55 (%)
Total: 991 Normal: 91.9 (%) Deficient: 6.86 (%)
log(CIV)
log(CI)
Mutation level (%)
0
20
40
60
80
100
Unknown
Normal
Deficient
RC status

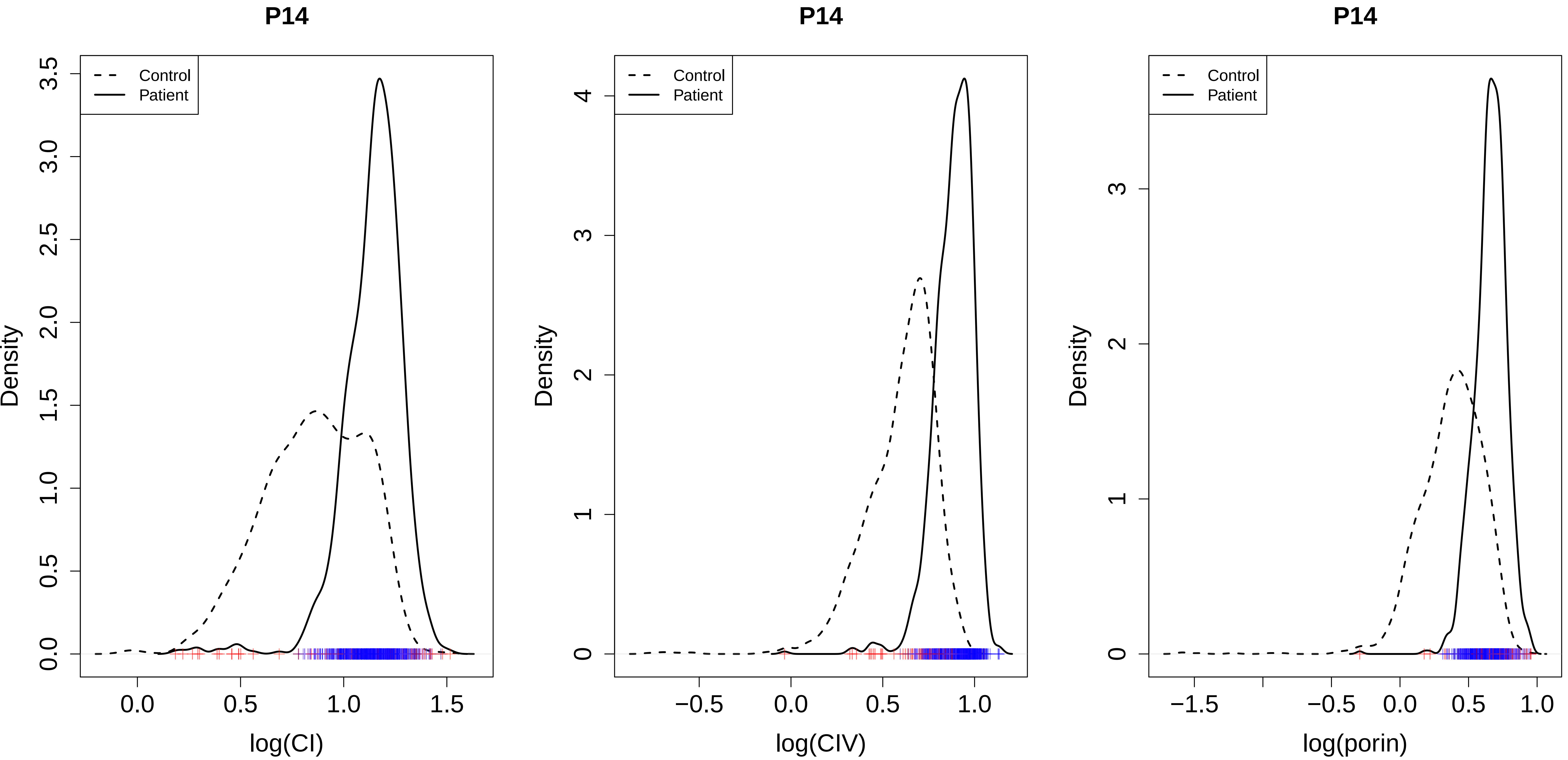
P14
Control
Patient
Density
log(CI)
P14
Control
Patient
Density
log(CIV)
P14
Control
Patient
Density
log(porin)

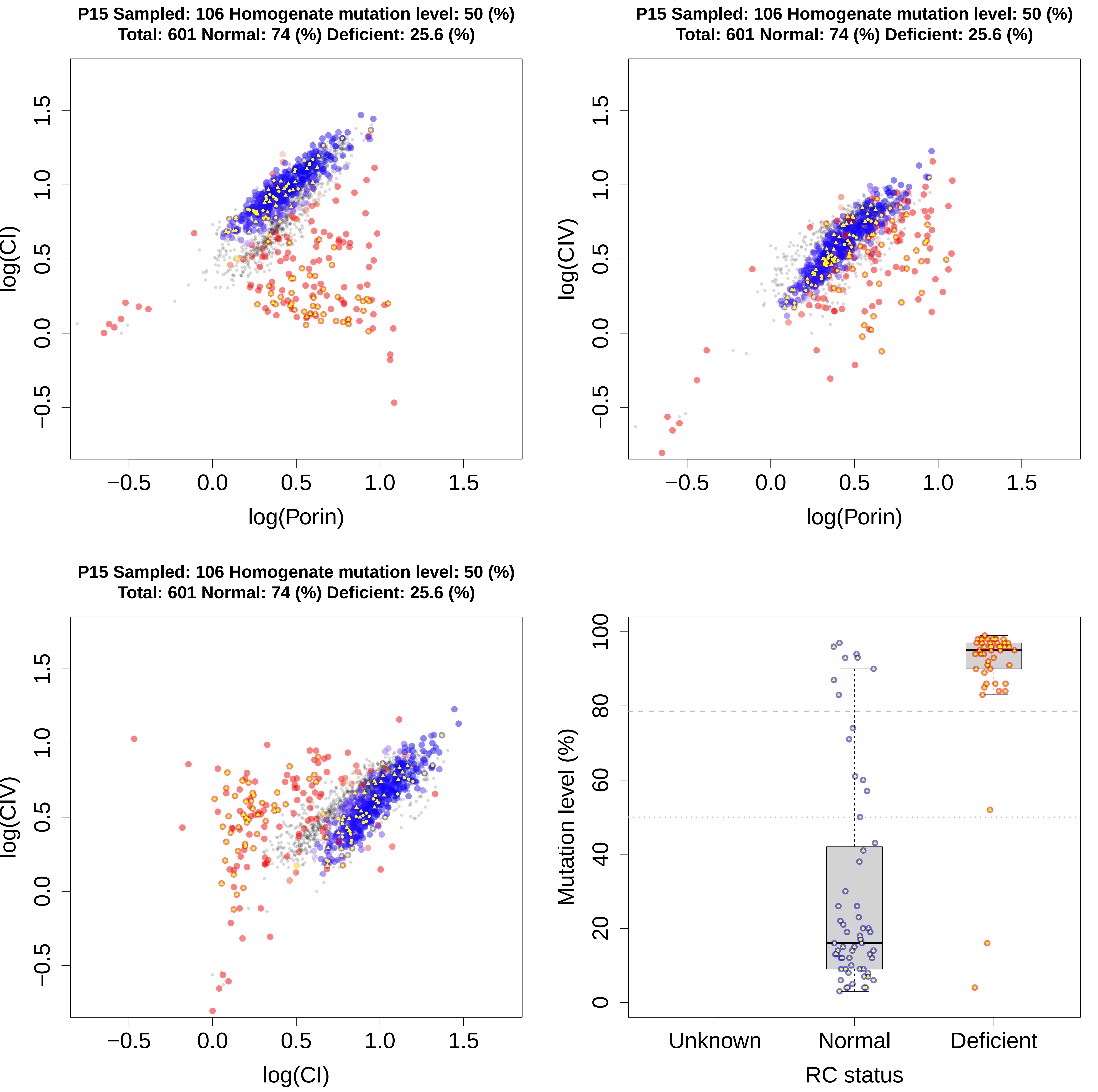
P15 Sampled: 106 Homogenate mutation level: 50 (%)
Total: 601 Normal: 74 (%) Deficient: 25.6 (%)
log(CI)
log(Porin)
−0.5
0.0
0.5
1.0
1.5
P15 Sampled: 106 Homogenate mutation level: 50 (%)
Total: 601 Normal: 74 (%) Deficient: 25.6 (%)
log(CIV)
log(Porin)
P15 Sampled: 106 Homogenate mutation level: 50 (%)
Total: 601 Normal: 74 (%) Deficient: 25.6 (%)
log(CIV)
log(CI)
Mutation level (%)
0
20
40
60
80
100
Unknown
Normal
Deficient
RC status

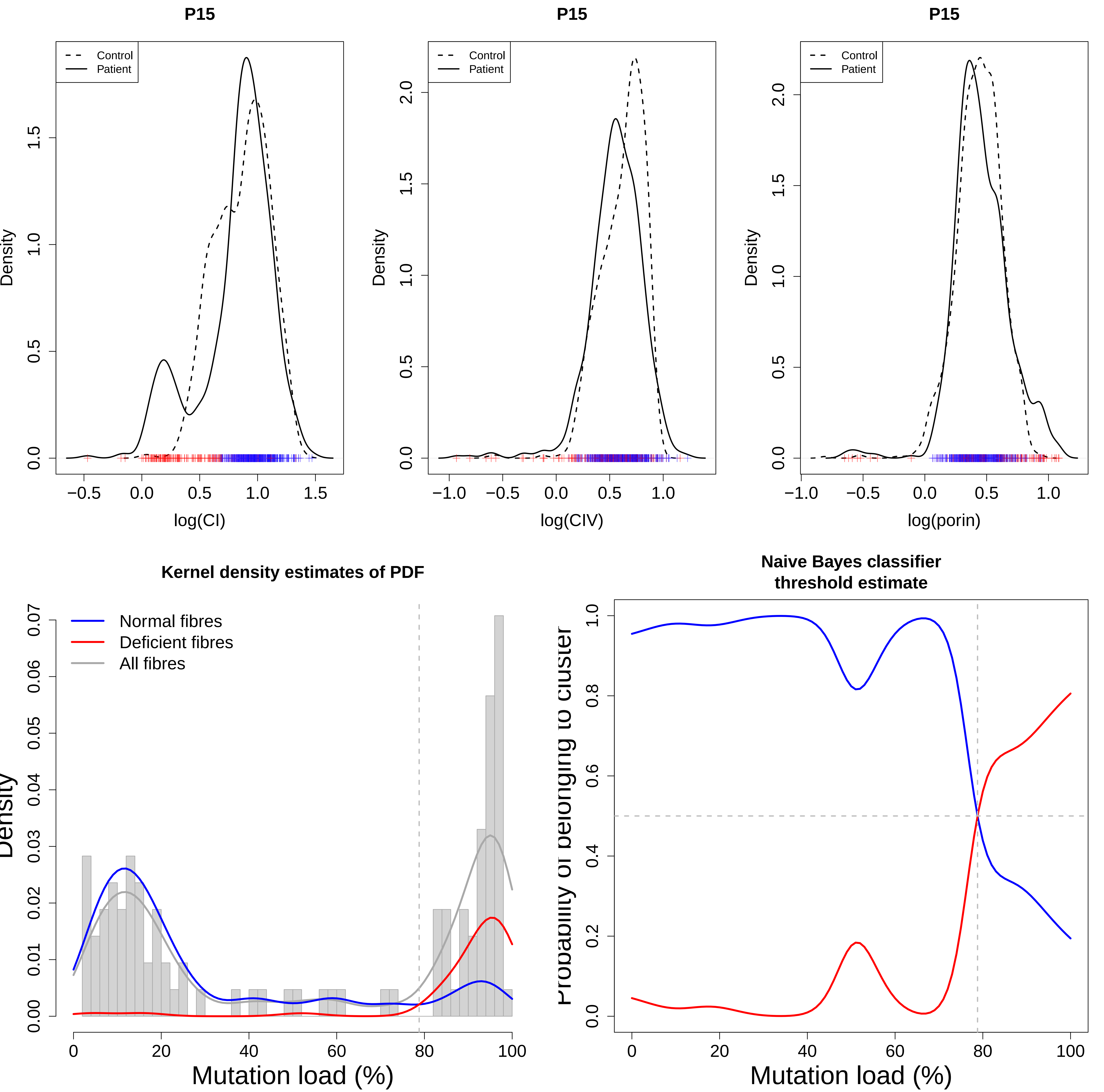
P15
Control
Patient
Density
log(CI)
P15
Control
Patient
Density
log(CIV)
P15
Control
Patient
Density
log(porin)
Kernel density estimates of PDF
Normal fibres
Deficient fibres
All fibres
Density
Mutation load (%)
Naive Bayes classifier
threshold estimate
Probability of belonging to cluster
Mutation load (%)

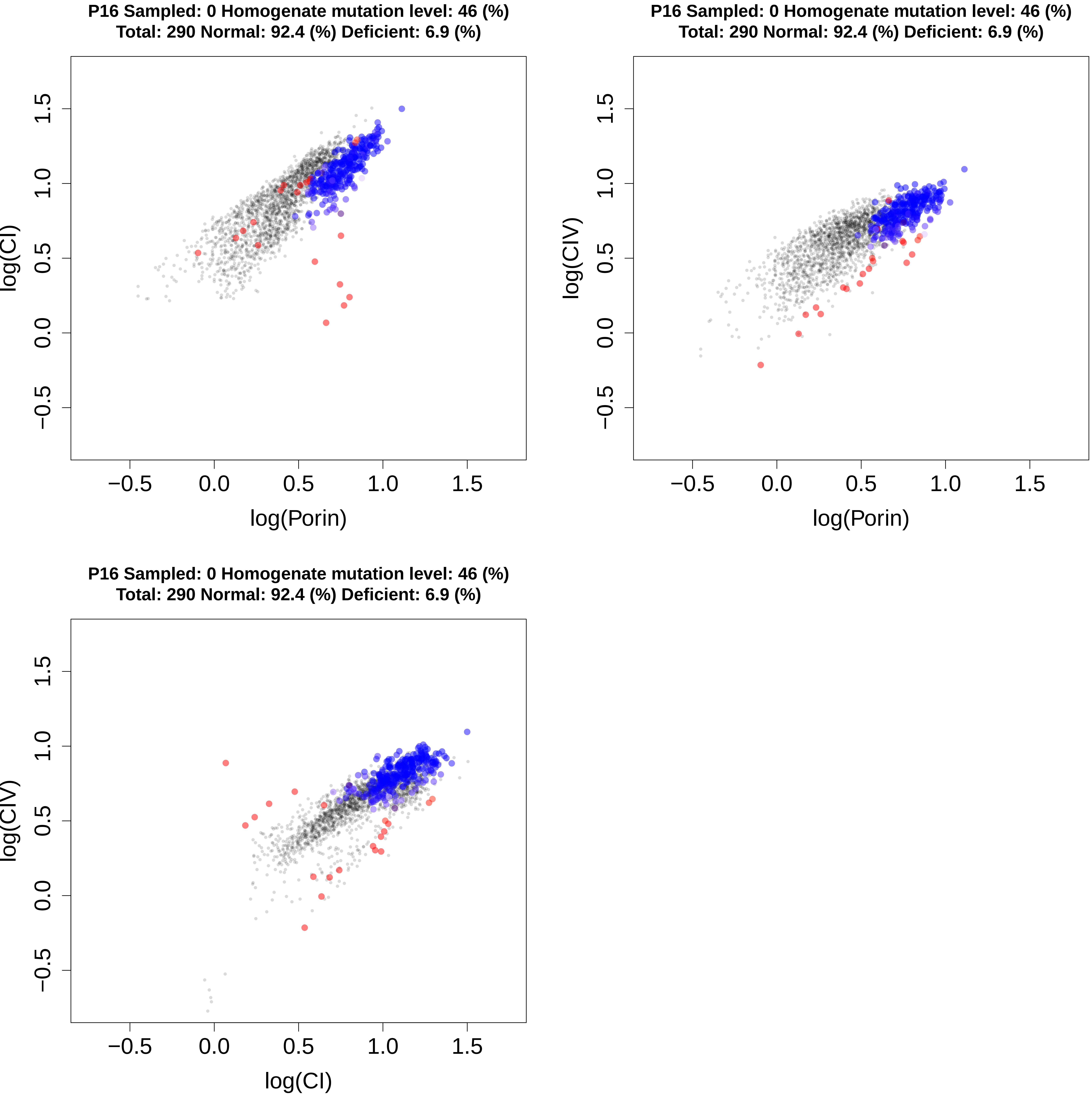
P16 Sampled: 0 Homogenate mutation level: 46 (%)
Total: 290 Normal: 92.4 (%) Deficient: 6.9 (%)
log(CI)
log(Porin)
P16 Sampled: 0 Homogenate mutation level: 46 (%)
Total: 290 Normal: 92.4 (%) Deficient: 6.9 (%)
log(CIV)
log(Porin)
P16 Sampled: 0 Homogenate mutation level: 46 (%)
Total: 290 Normal: 92.4 (%) Deficient: 6.9 (%)
log(CIV)
log(CI)
−0.5
0.0
0.5
1.0
1.5

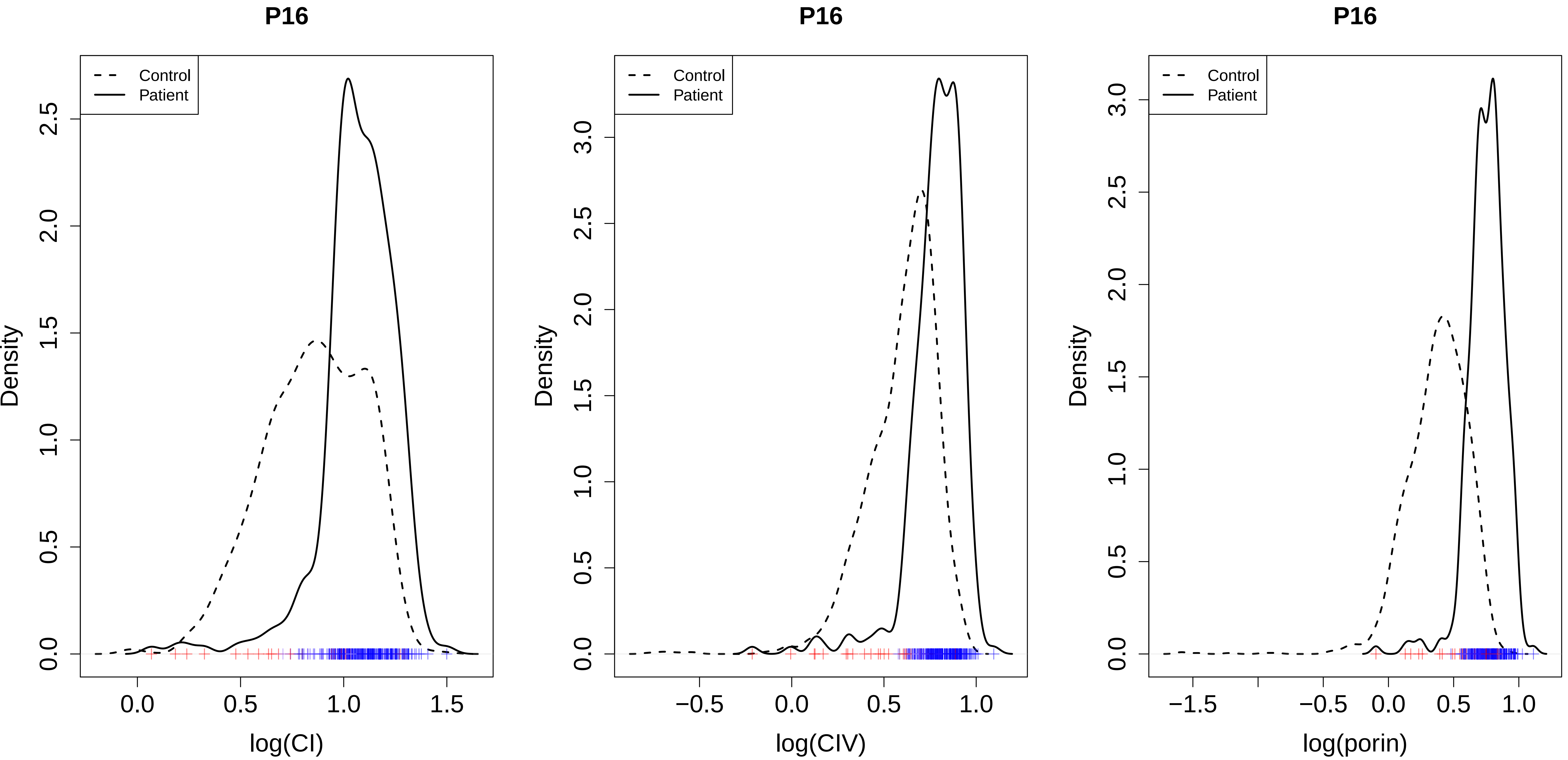
P16
Control
Patient
Density
log(CI)
P16
Control
Patient
Density
log(CIV)
P16
Control
Patient
Density
log(porin)

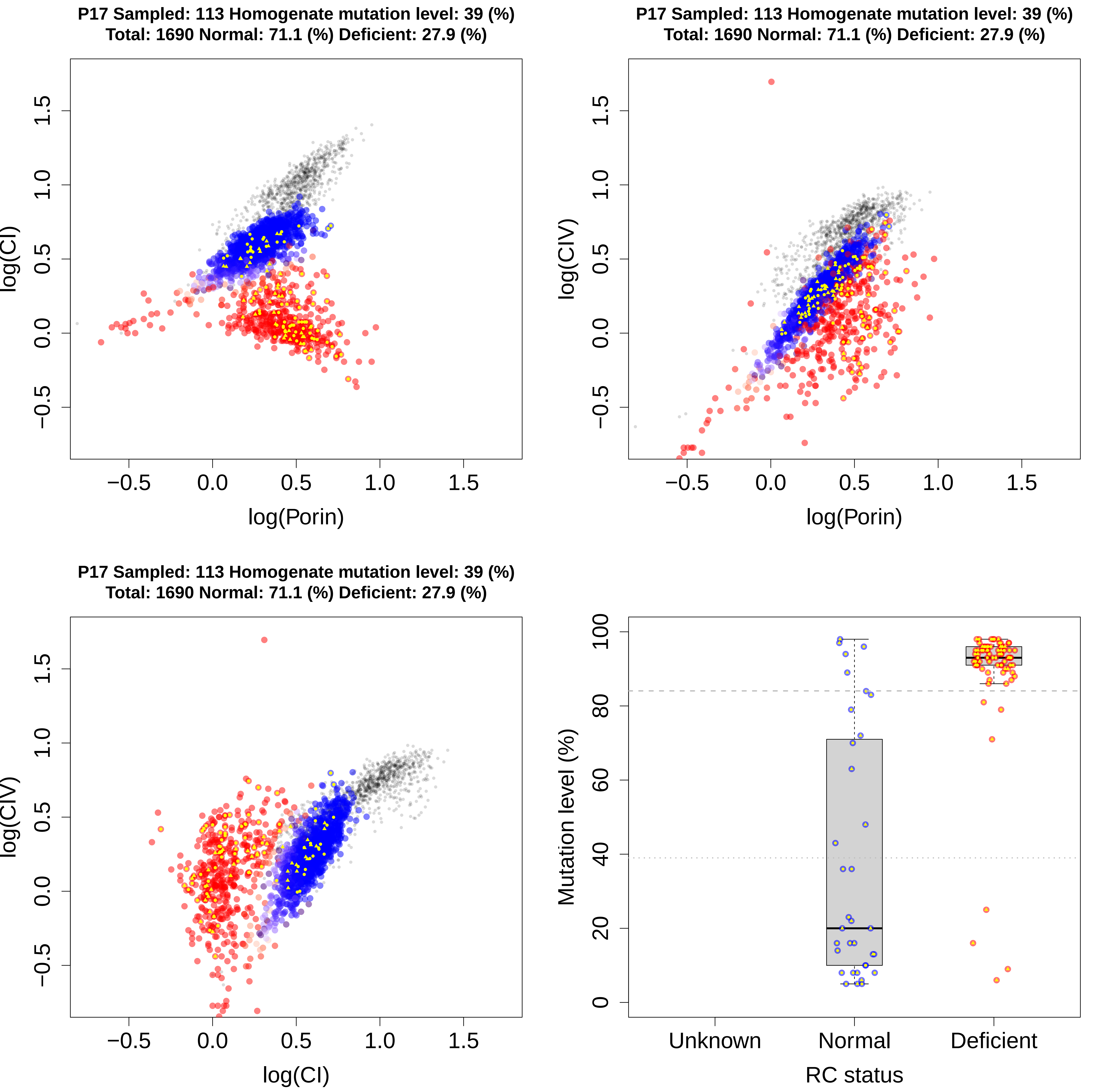

P17 Sampled: 113 Homogenate mutation level: 39 (%)
Total: 1690 Normal: 71.1 (%) Deficient: 27.9 (%)
log(CI)
log(Porin)
−0.5 0.0 0.5 1.0 1.5
P17 Sampled: 113 Homogenate mutation level: 39 (%)
Total: 1690 Normal: 71.1 (%) Deficient: 27.9 (%)
log(CIV)
log(Porin)
−0.5 0.0 0.5 1.0 1.5
P17 Sampled: 113 Homogenate mutation level: 39 (%)
Total: 1690 Normal: 71.1 (%) Deficient: 27.9 (%)
log(CIV)
log(CI)
−0.5 0.0 0.5 1.0 1.5
Mutation level (%)
0 20 40 60 80 100
Unknown
Normal
Deficient
RC status

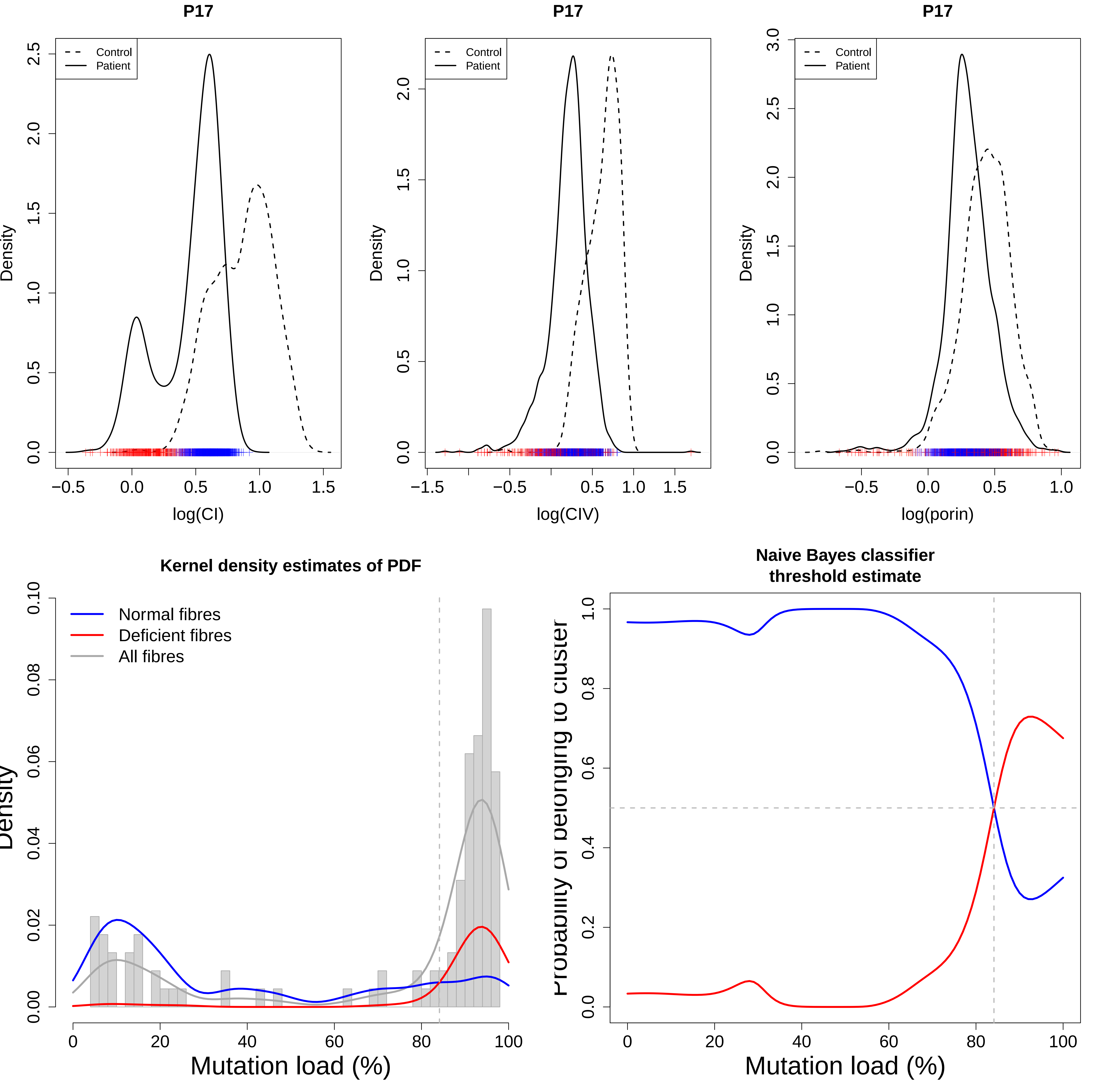

P17
Control
Patient
Density
log(CI)
P17
Control
Patient
Density
log(CIV)
P17
Control
Patient
Density
log(porin)
Kernel density estimates of PDF
Normal fibres
Deficient fibres
All fibres
Mutation load (%)
Naive Bayes classifier
threshold estimate
Mutation load (%)

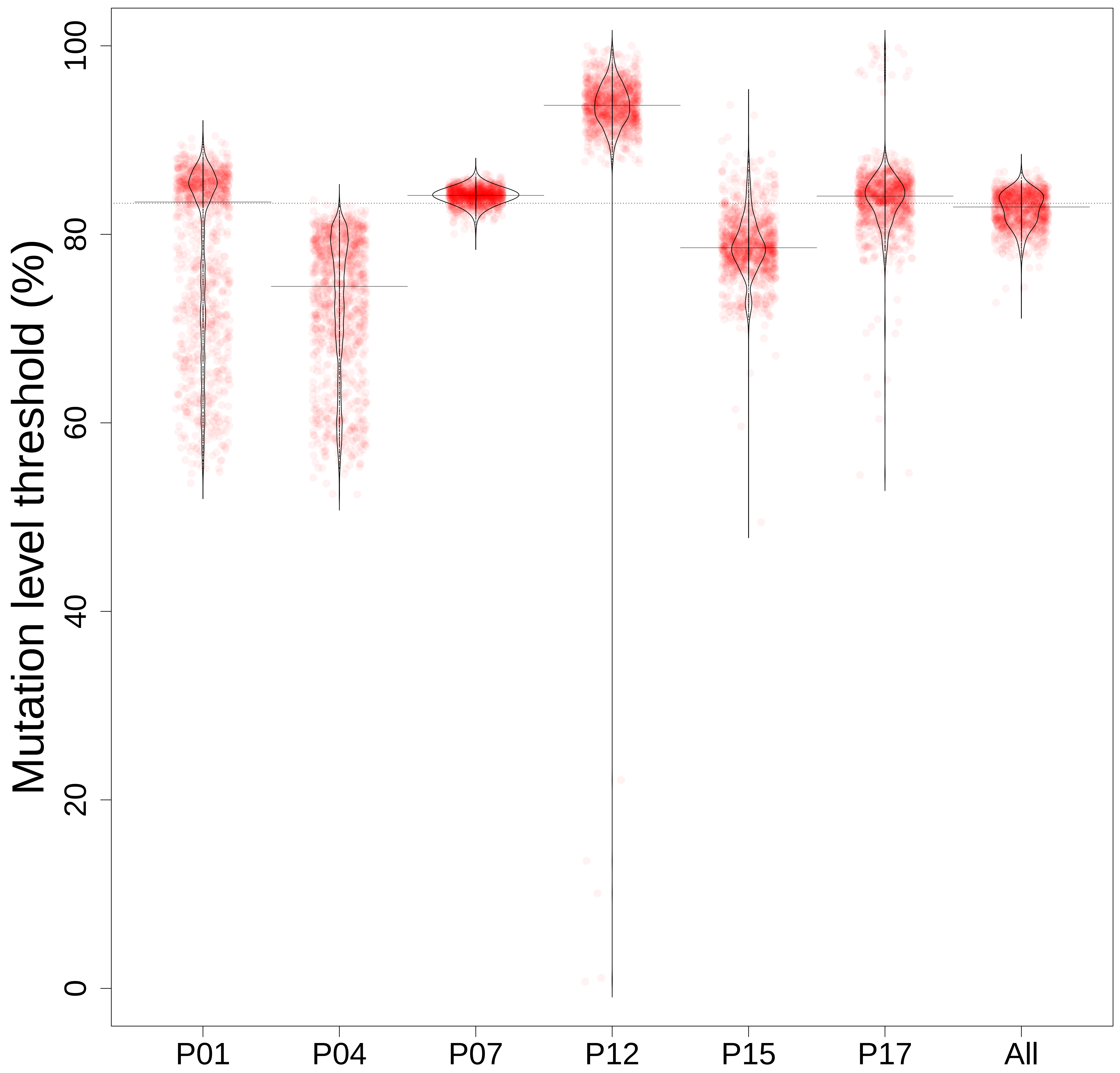
Mutation level threshold (%)
0
20
40
60
80
100
P01
P04
P07
P12
P15
P17
All
